# Temporal Trends in Behavioural Risk Factors for Cancers with Rising Incidence in Younger Adults: An Analysis of Population-Based Data in England

**DOI:** 10.1101/2025.08.21.25333984

**Authors:** Montserrat García-Closas, Zoey Richards, Reuben Frost, Marc J. Gunter, Amy Berrington de Gonzalez

## Abstract

**Objective:** To assess whether changes in behavioural risk factors could explain rising cancer incidence in younger adults in England, and to evaluate the extent to which established and suspected risk factors contribute to these trends

**Methods and Analysis:** Cancer incidence data from national registries (2001-2019) identified cancers with increasing incidence in adults aged 20-49. Trends in smoking, alcohol, diet, body mass index (BMI) and physical inactivity were examined using national health surveys. Annual percentage changes quantified trends by age and sex. Population attributable fractions (PAFs) estimated the proportion of cancers attributable to risk factors and disaggregated attributable from non-attributable incidence rates.

**Results:** Eleven cancers with established behavioural risk factors showed rising incidence in younger adults. Similar trends were observed in older adults, except for colorectal and ovarian cancer, which increased only in younger adults. For some cancers, incidence increased more rapidly in younger than older adults. PAFs for younger adults ranged from 7% to 65% depending on cancer type. All risk factors except obesity showed stable or declining prevalence. For BMI-related cancers, both BMI-attributable and non-attributable incidence increased, though more slowly for the latter. For example, BMI-attributable colorectal cancer in younger women increased from 0.9 to 1.6 per 100,000 (APC 4.3%), while non-attributable rates rose from 6.4 to 9.6 (APC 3.2%).

**Conclusions:** Behavioural risk factors account for a substantial share of cancer burden but, apart from BMI, are unlikely to explain the rising incidence in younger adults. Findings underscore the urgent need to investigate emerging risk factors, while strengthening prevention efforts targeting known factors across all ages.

**Significance of this study:** *What is already known on this topic:* Incidence rates for several cancers are rising among younger adults in England and many other countries. Whether changes in risk factor prevalence explain these trends is unclear and has not been systematically assessed using population-level data.

*What this study adds:* In England (2001-2019), nine of eleven cancers with established behavioural factors that are increasing in younger adults are also rising in older adults where the burden is greatest. Colorectal and ovarian cancers are exceptions. Rates for several cancers in younger adults, rates have stabilised or declined in recent years. Except for BMI, most behavioural risk factors show stable or declining trends. While BMI is a key contributor, it alone is unlikely to explain rising early-onset cancer rates.

*How this study might affect research, practice or policy:* Highlights the need to investigate novel and emerging exposures, while strengthening prevention strategies targeting known risk factors, particularly obesity, across all ages. There is also a need to assess the impact of changing diagnosis and screening practices on incidence trends.

## Introduction

The incidence of cancer in young adults has been increasing for several cancer types in the UK and many other countries, raising questions on potential underlying causes and concerns about the public health implications^1–5^. In the USA, many obesity-related cancers that increased in younger adults between 1995 and 2014 were also rising in older adults, though often at a slower rate^3,6^. A notable exception is colorectal cancer for which incidence has declined in older adults in the USA and has shown stable or decreasing trends in several countries, including the UK^6^. However, how incidence trends for other cancers in the UK compare between younger and older adults is less clear. These comparisons can provide valuable insights into the potential drivers of these increases, e.g. whether younger adults are more susceptible to certain exposures or whether specific risk factors have emerged or intensified at a particular point in time, leading to cohort effects.^1,3^

Multiple established and emerging risk factors have been proposed to explain rising cancer incidence in younger adults ^1,2^. These include established behavioural risk factors such as alcohol use, obesity, physical inactivity, red or processed meat consumption and low fibre intake. Additionally, there are growing concerns about suspected risk factors such as ultra-processed foods, sweetened beverages, environmental exposures (e.g., air pollution, water contaminants, as poly and perfluoroalkyl substances (PFAS), changes in sleep patterns (e.g., light at night), and prenatal and early-life exposures (e.g., antibiotic use during infancy). Despite the extensive list of proposed risk factors for rising cancers in younger adults, there has been no formal evaluation of how temporal trends in these factors align with observed increases in cancer incidence in the UK.

To address these gaps, we analyse cancer incidence trends in England from 2001 to 2019, comparing patterns in younger (age 20-49 years) and older (age ≥50 years) adults. We then focus on cancers with rising incidence in younger adults and established behavioural risk factors to assess whether exposure trends could explain increases in incidence. Finally, we evaluate the evidence for suspected risk factors, by assessing available population data and theoretical calculations. Our aim is to assess the plausibility and potential impact of specific risk factors on rising cancer incidence, and to inform future research directions and prevention strategies.

## Methods

Below is a brief description of the methods used, more details can be found in **Supplementary Methods.**

### Data Sources

#### Cancer Incidence Data

Age-specific cancer incidence data was obtained from the National Disease Registration Service (NDRS) for England from 2001 to 2019 (**Supplementary Table 1**) stratified by age group (20-49, ≥50) and sex (men and women). Cancer incidence rates within the two age groups were age-standardised using the 2013 European Standard Population (ESP)^7^. International Classification of Diseases (ICD version 10) codes were grouped into 25 cancer sites, using groupings similar to Globocan^8^.

#### Behavioural Risk Factor Data

Established behavioural risk factors were defined as those classified as Group 1 carcinogens by the International Agency for Research on Cancer (IARC); or having strong evidence for cancer risk associations by the World Cancer Research Fund (WCRF) (**Supplementary Table 2**). These included cigarette smoking, elevated body mass index (BMI), alcohol consumption, red and processed meat consumption, low fibre intake, and physical inactivity. Data on these risk factors was obtained from population-based surveys in England according to age and sex (**Supplementary Table 1, Supplementary Table 3**).

### Statistical Analyses

#### Cancer Incidence and Risk Factor Trends

Temporal trends in age-standardised cancer incidence rates and risk factor prevalence in England were visualised using time series plots on a log scale to reflect relative changes over time. To quantify changes, we estimated Average Annual Percentage Change (AAPC; 2001-2019) with corresponding 95% confidence intervals (CI) by age and sex groups for 25 cancer sites^9^. On a log scale, parallel lines of cancer incidence rates indicate similar AAPCs across age groups. The most recent APC was calculated for cancer sites with significant and positive AAPCs in younger adults, which also had established associations with behavioural risk factors (as defined above). Risk factor AAPCs were calculated over the timeframe with available data that varied for different risk factors. AAPC were estimated by age, sex, and index of multiple deprivation (IMD) groups^10^. All AAPC and APC calculations were done using the Joinpoint regression software^11^. To assess whether incidence rates in younger adults were increasing faster than in older adults, we conducted a one-sided test comparing the AAPCs between the two age groups.

#### Population Attributable Fractions (PAF)

We estimated the Population Attributable Fraction (PAF), i.e. the proportion of cancer cases attributable to a given risk factor in the population. PAFs were calculated by age group and sex according to the following formula:

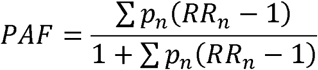

Where:

p_n_ = Population prevalence for the n^th^ level of the risk factor

RR_n_ = Relative Risk for the n^th^ level of the risk factor

PAFs aggregated across risk factors for each cancer site assumed independence between risk factors, and were calculated by combining risk factor specific PAFs, with each additional risk factor applied only to the remaining cancer cases not attributed to a previous exposure. To account for the expected delay between risk factor exposure and cancer diagnosis, a 10-year time lag was applied in estimating attributable cases^4,12,13^. The 95% confidence intervals for PAF estimates were estimated using Monte Carlo simulation methods.

#### Incidence Rates Attributable and Non-Attributable to Risk Factors

From the earliest available data to 2019, yearly PAFs were calculated for each cancer site and risk factor combination if the risk factor showed a statistically significant and increasing AAPC in the 20-49 age group for men or women. Risk factor specific attributable and unattributable incidence rates were calculated across periods with available data, focusing on risk factors whose prevalence significantly increased in younger adults (AAPC>0 and p<0.05). For multi-level risk factors, trends were assessed using the prevalence of the highest exposure categories (i.e. current smoking, moderate/heavy drinking and obesity). AAPCs were estimated for both serial datasets using Joinpoint software to calculate the relative change in attributable and non-attributable cancer incidence across periods with available data^11^.

### Code and data accessibility

All code and publicly available datasets used in this analysis can be found on <u>GitHub</u>. Individual level data on risk factors can be obtained through an End User License Agreement with the UK Data Service (UKDS). Links to all used datasets are available in **Supplementary Table 1**.

### Patient and Public Involvement

Patients and members of the public were not directly involved in the design, conduct, or reporting of this study. However, the national health surveys and cancer registration data utilised are developed and maintained with ongoing patient and public involvement, including advisory boards and consultations that inform data collection and governance. The research questions addressed also reflect priorities informed by public engagement in related studies on cancer risk and prevention. Future work will incorporate input from public and patient representatives to guide dissemination and further research on early-onset cancer.

## Results

### Cancer incidence trends

The incidence for 16 of 22 cancers in younger women and 11 of 21 cancers in younger men increased significantly in England during 2001 to 2019 (AAPC>0, P<0.05) (**Supplementary Figure 1**). Further analyses focused on cancers with significantly increasing trends in younger adults that also had established behavioural risk factors as defined by IARC (group 1) or WCRF (strong evidence). Eleven cancers met these criteria: thyroid, multiple myeloma, liver, kidney, gallbladder, colorectal, pancreatic, endometrial, oral, breast and ovarian cancers (**Supplementary Methods; Supplementary Figure 2**).

**Figure 1** illustrates the temporal trends in age-standardised incidence rates for the eleven selected cancers by age groups and sex. Except for colorectal and ovarian, cancers with significant increases in younger adults also increased significantly in older adults (i.e. AAPC>0, P <0.05; **Supplementary Table 4**). While colorectum cancer rates in older adults were stable (AAPC P≥0.05), ovarian rates were significantly decreasing by 1.31%/year in older women. For all other cancers increasing in both age groups, five out of nine (endometrium, kidney, pancreatic, multiple myeloma and thyroid cancer) increased significantly faster in younger than older women; and one out of seven (multiple myeloma) increased faster in younger than older men (**Supplementary Table 4**). Examination of the most recent APCs revealed that several cancers that increased in younger adults over the 2001 to 2019 period had stable or declining trends in the most recent period, namely breast (APC_2013-2019_ =0.21, P=0.62) and ovarian (APC_2001-2019_ =-0.57, P=0.58) cancer in women, kidney (APC_2016-2019_ =-0.67, P=0.84), liver (APC_2014-2019_ =-3.52, P=0.36), and oral (APC_2014-2019_ =-1.07, P=0.06) cancers in men (**Supplementary Table 4**).

**Figure 1:**
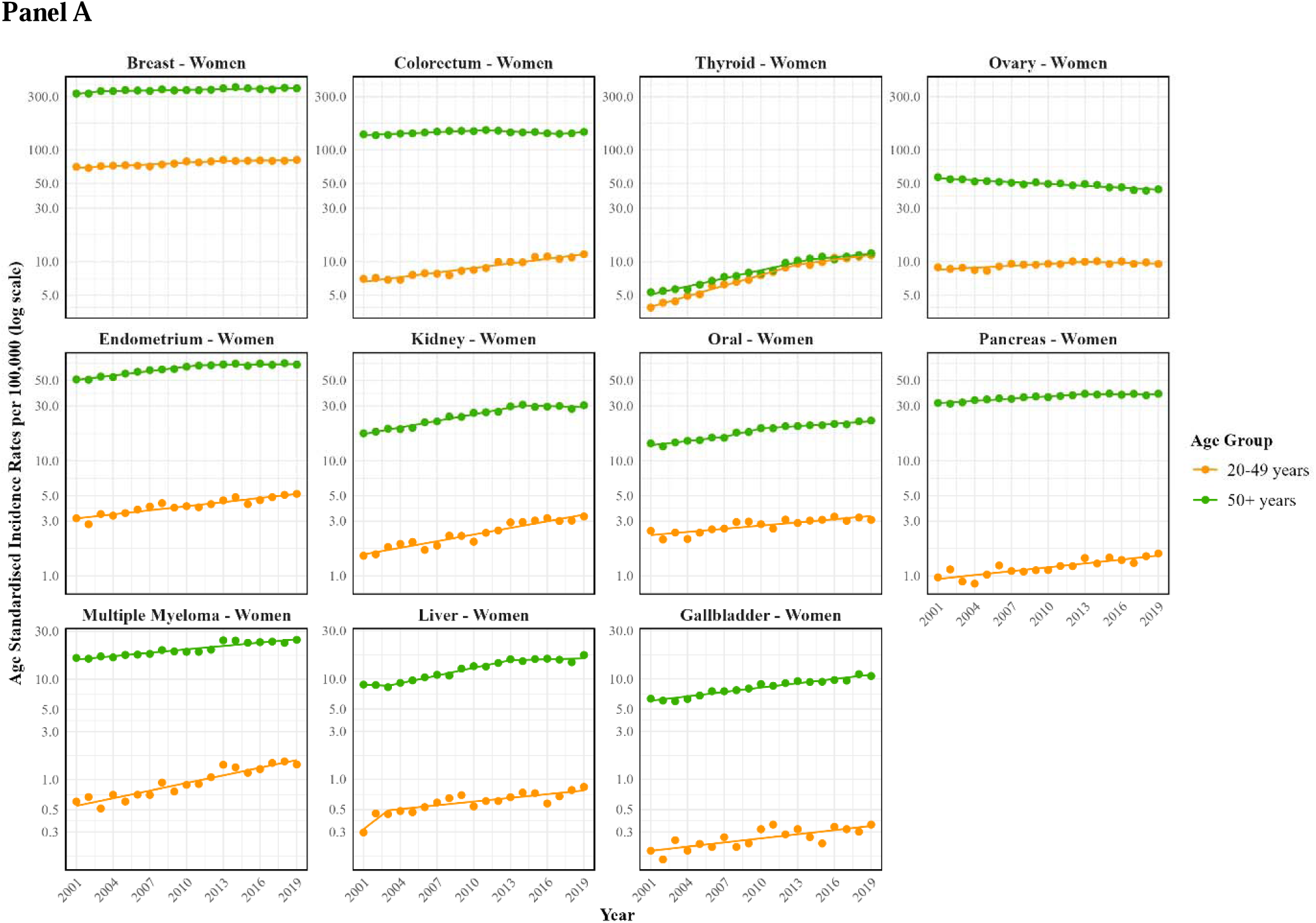
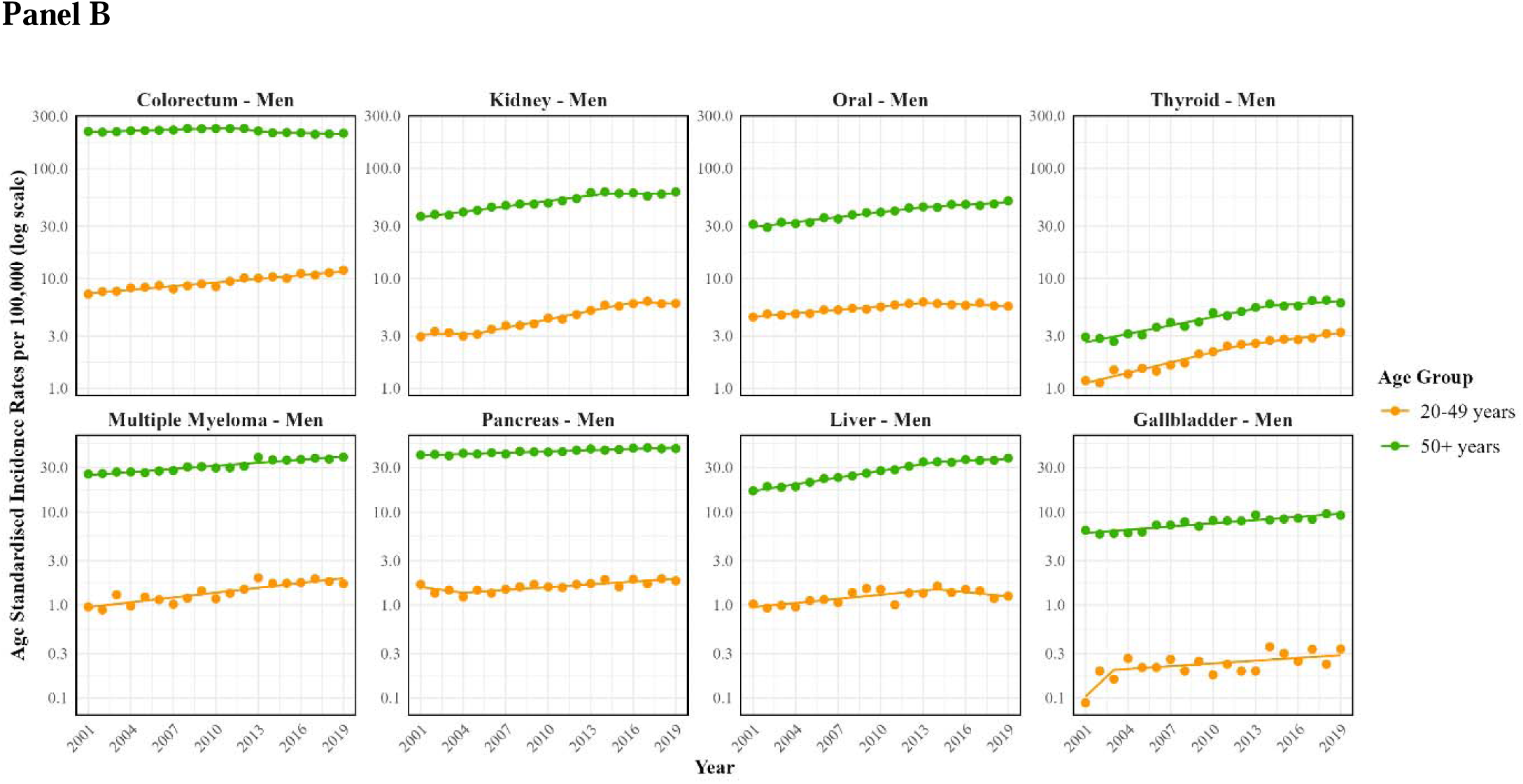
Temporal trends in 10 selected cancer sites with incidence rates increasing in younger adults (20-49 years) in England (2001-2019), and with established behavioural risk factors, by age groups for women (**Panel A**) and men (**Panel B**). Plots have different y-scales and sorted from highest to lowest incidence in younger adults. Straight lines represent the Joinpoint annual percent changes (APC). Estimates of the most recent APC and average APC (AAPC) from 2001 to 2019 are shown in **Supplementary Table 4**.

### Behavioural risk factor trends

We assessed trends for seven established behavioural risk factors associated with the eleven selected cancers increasing in younger adults (Figure 2**, Supplementary Table 5**). All these cancers (except oral) are obesity-related, while six are linked to smoking (liver, colorectal, oral, pancreas, kidney, ovary), four with alcohol (liver, colorectal, oral, breast), three with physical inactivity (colorectal, breast, endometrial), and one with dietary factors (colorectal). Of note, colorectal cancer is linked to all factors examined. Except for BMI, trends in these risk factors over the past one to two decades were stable or improving for younger adults. Cigarette smoking had a ∼ 2%/year relative decrease in prevalence in younger men and women since 1995. In 2019, a lower percentage of younger women than men were current smokers (∼20% vs 25%, respectively). Younger adult drinking trends were decreasing or stable across sex and exposure levels except for light drinking in younger men. Overall, moderate and heavy drinking prevalence remained lower in women than men. Specifically, the prevalence of heavy alcohol drinking in younger men had a relative decline of 3.6%/year between 2011 and 2019 from a prevalence of 7% to 5%. Moderate drinking was more common (∼20% of younger women, 30% of younger men in 2019), with levels also declining since 2011 by 3.0%/year for younger women and 2.2%/year for younger men. Similarly, physical inactivity decreased across sex groups from 2003 to 2012. By 2016, based on updated guidelines, levels of physical inactivity remained lower in younger men (20%) than women (35%). Although risk factor levels varied by age groups, the temporal trends were generally similar.

**Figure 2.**
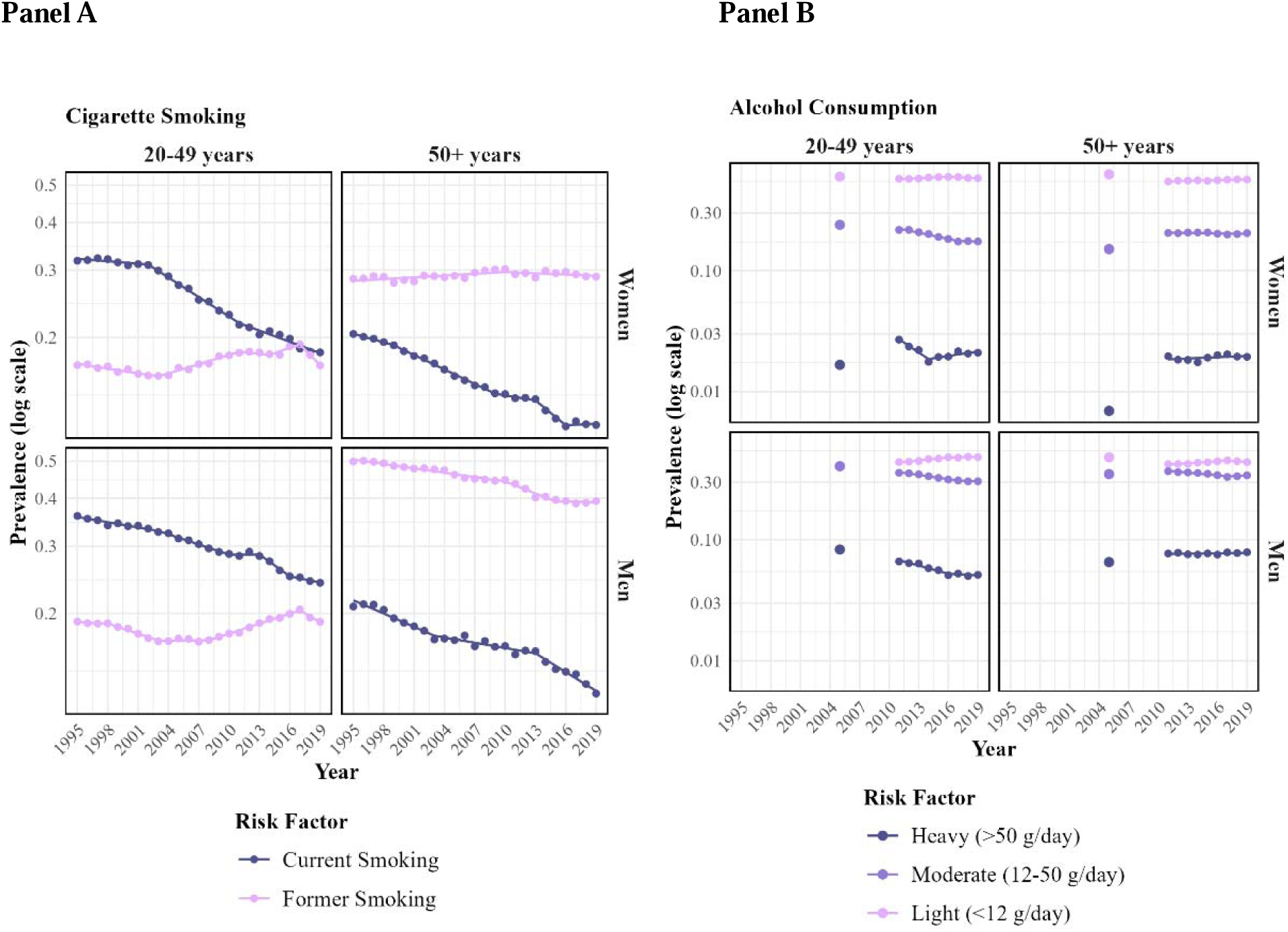
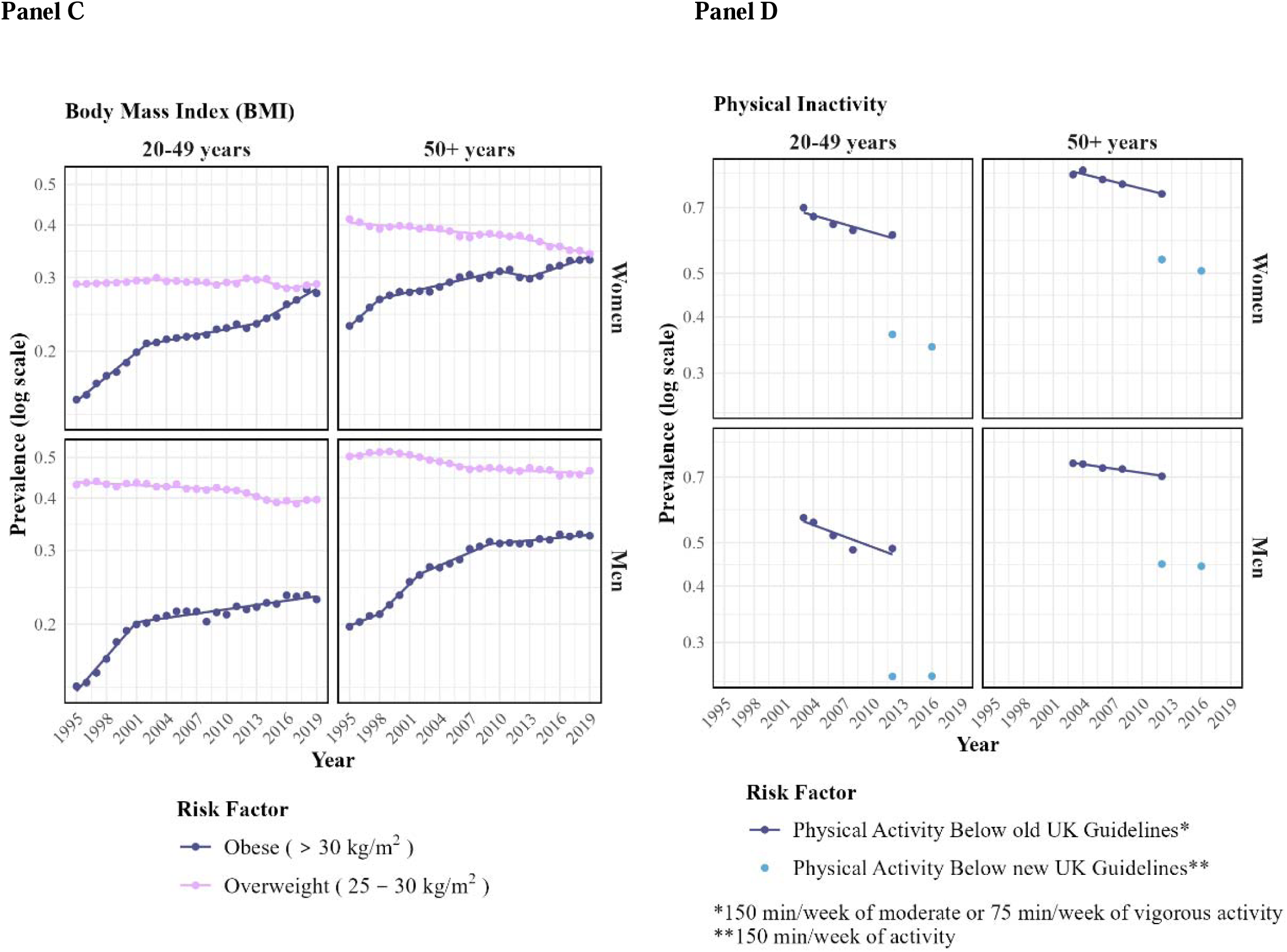
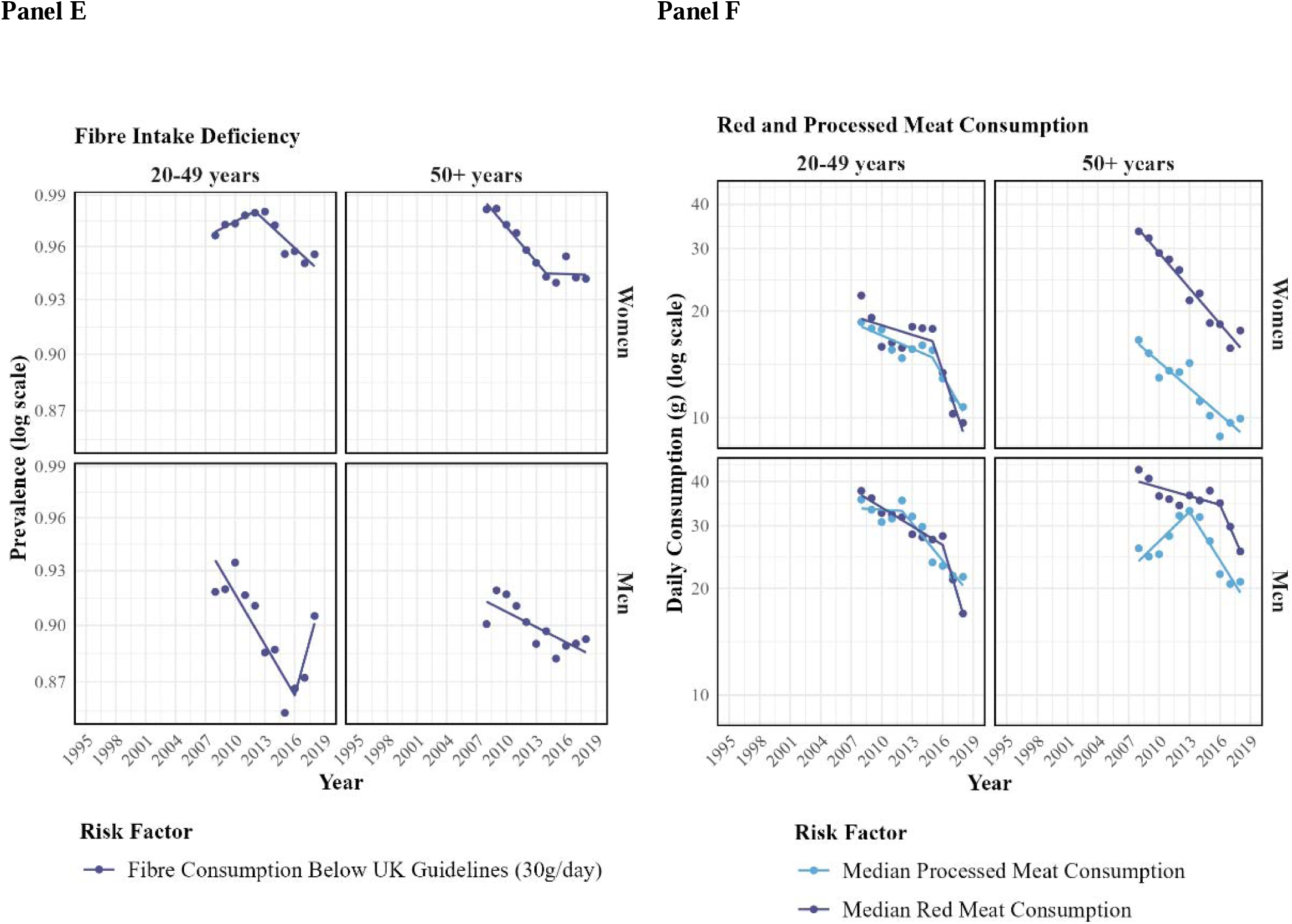
Temporal trends in established behavioural risk factors in England (1995-2019) for 11 selected cancer sites with raising incidence rates in younger adults, by age and sex groups. Straight lines represent the Joinpoint annual percent changes (APC). Estimates of the most recent APC and average APC (AAPC) from earliest available year to 2019 are shown in **Supplementary Table 5**. Risk factors: cigarette smoking (**Panel A**), alcohol consumption (**Panel B**), body mass index (BMI) (**Panel C**), physical inactivity (**Panel D**), fibre intake deficiency (**Panel E**), and red and processed meat consumption (**Panel F**).

Data on dietary consumption trends was available from 2008 to 2018 and showed sharp declines in the consumption of processed and red meat across all age and sex groups. The largest reductions were observed in red meat consumption, with relative annual decreases of 7.4% in younger men and 7.1% in younger women. This resulted in the median amount of red meat consumed per day for younger men decreasing from 38g in 2008 to 17g in 2018, and from 22g to 10g in younger women. Median processed meat consumption levels were consistently lower in younger women (∼10g in 2018) compared to men (∼20g in 2018). The percentage of younger adults with fibre intake deficiency was very high (>90% in 2018), but remained stable or declined across men (-0.4%/year) and women (-0.2%/year) from 2008 to 2018. Trends in younger adults were similar to those in older adults.

In contrast, the prevalence of obesity has increased steadily since 1995 for all adults. The largest increases in obesity are for younger women with about +2.6% per year relative increase since 1995. The lowest increases were for older women whose obesity rates have been only slightly increasing since 2009. By 2019, obesity prevalence was about 30% of older adults and younger women, and 23% of younger men. The prevalence of overweight has been stable or declining across all population groups, but in 2019 were still very high (29-34% in younger and older women, and 40%-46% in younger and older men respectively).

To explore the impact of contextual factors, we evaluated temporal trends of established risk factors in adults by the index of multiple deprivation (IMD) during the periods with data available. All ages were combined to avoid small sample sizes within IMD and age groups. We found decreasing trends for current smoking, physical inactivity, and moderate/heavy alcohol consumption, and increasing trends of obesity across all deprivation groups (**Supplementary Figure 3**). However, there were differences in risk factor prevalences and their rate of decrease. The clearest patterns were for smoking and obesity. Current smoking had a slower decrease in the most deprived compared to the least deprived groups. In contrast, obesity prevalence increased across all deprivation groups with sharper increases and higher levels for the most deprived compared to the least deprived groups.

### Population attributable fractions (PAFs) and exposure attributable incidence rates

Aggregated 2019 PAFs for the seven established behavioural risk factors and BMI-specific PAFs are shown for each cancer by age groups in **Figure 3**. Differences in PAFs by age were small with overlapping 95%CIs. While aggregated PAFs tended to be higher in men than women for most cancers, the BMI-specific PAFs tended to be higher in women than men. Differences in PAFs mainly reflected differences in exposure levels across age and sex groups, since RRs were assumed to be constant by age and sex, except for BMI and breast cancer risk (**Supplementary Table 3).**

**Figure 3:**
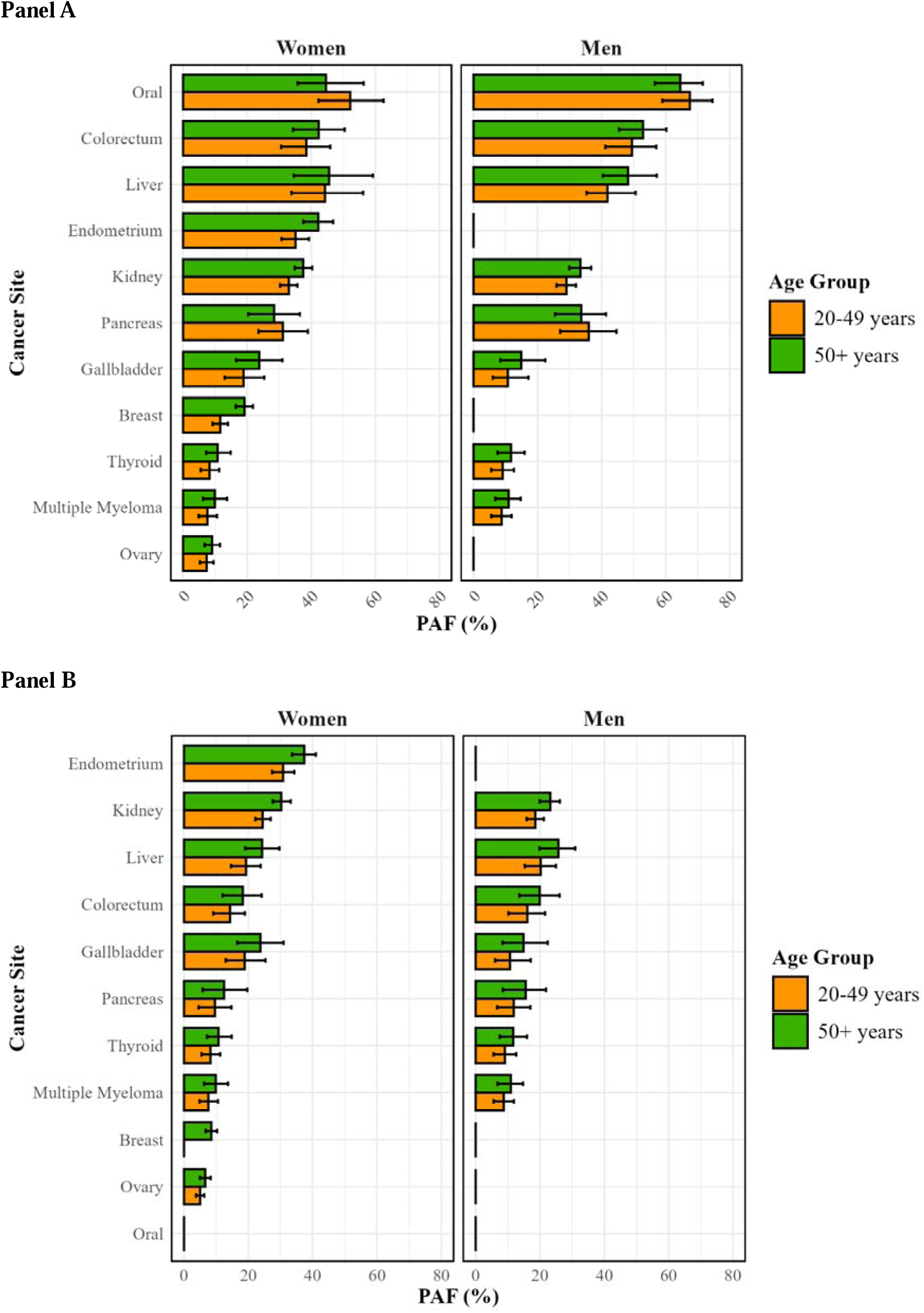
Aggregated (**Panel A**) and BMI (**Panel B**) population attributable fractions (PAF) for behavioural risk factors in 10 cancer sites by age group and sex in England, 2019

In men, the cancers with the highest PAFs (>20%) in 2019 were oral (PAF=68-65% for younger and older adults, respectively), liver (42-48%), colorectal (49-53%), kidney (29-33%), and pancreatic cancers (36-34%). In women, the highest PAFs were for oral (52-45%), endometrial (35-42%), liver (44-46%), colorectal (38-42%), kidney (33-37%), pancreas (31-28%) and gallbladder (19%-24%) cancers. PAF for specific risk factors across different cancer types are shown in **Supplementary Figure 4.** BMI is the risk factor associated with most cancers (all except oral cancer), with 2019 PAFs ranging from 8% for multiple myeloma to 37% for endometrial cancers.

For cancers linked to BMI, the only risk factor showing significantly increasing trends, we evaluated cancer incidence attributable and non-attributable to increases in BMI from 2005 to 2019, by age and sex groups (**Figure 4, Supplementary Figure 5, Supplementary Table 6**). Both, BMI-attributable and non-attributable rates increased in younger adults, although BMI-attributable rates increased faster (**Figure 4; Supplementary Table 6**). For example, BMI-attributable colorectal cancer rates in younger women increased by 4.3%/year from 0.9 to 1.7 per 100,000, while BMI non-attributable rates increased by 3.2%/year from 6.7 to 10.0 per 100,000. For endometrial cancer in younger women, BMI-attributable rates increased by 3.8%/year from 0.9 to 1.6 per 100,000, while BMI non-attributable rates increased by 1.7%/year from 2.6 to 3.6. Similar patterns of BMI-attributable and non-attributable rates were seen in younger and older adults, except for colorectal cancer. BMI-attributable incidence rates for colorectal cancer increased significantly by 0.8%/year in older men and by 0.7%/year in older women, while non-attributable rates were stable with non-significant AAPCs (**Supplementary Figure 5, Supplementary Table 6).**

**Figure 4:**
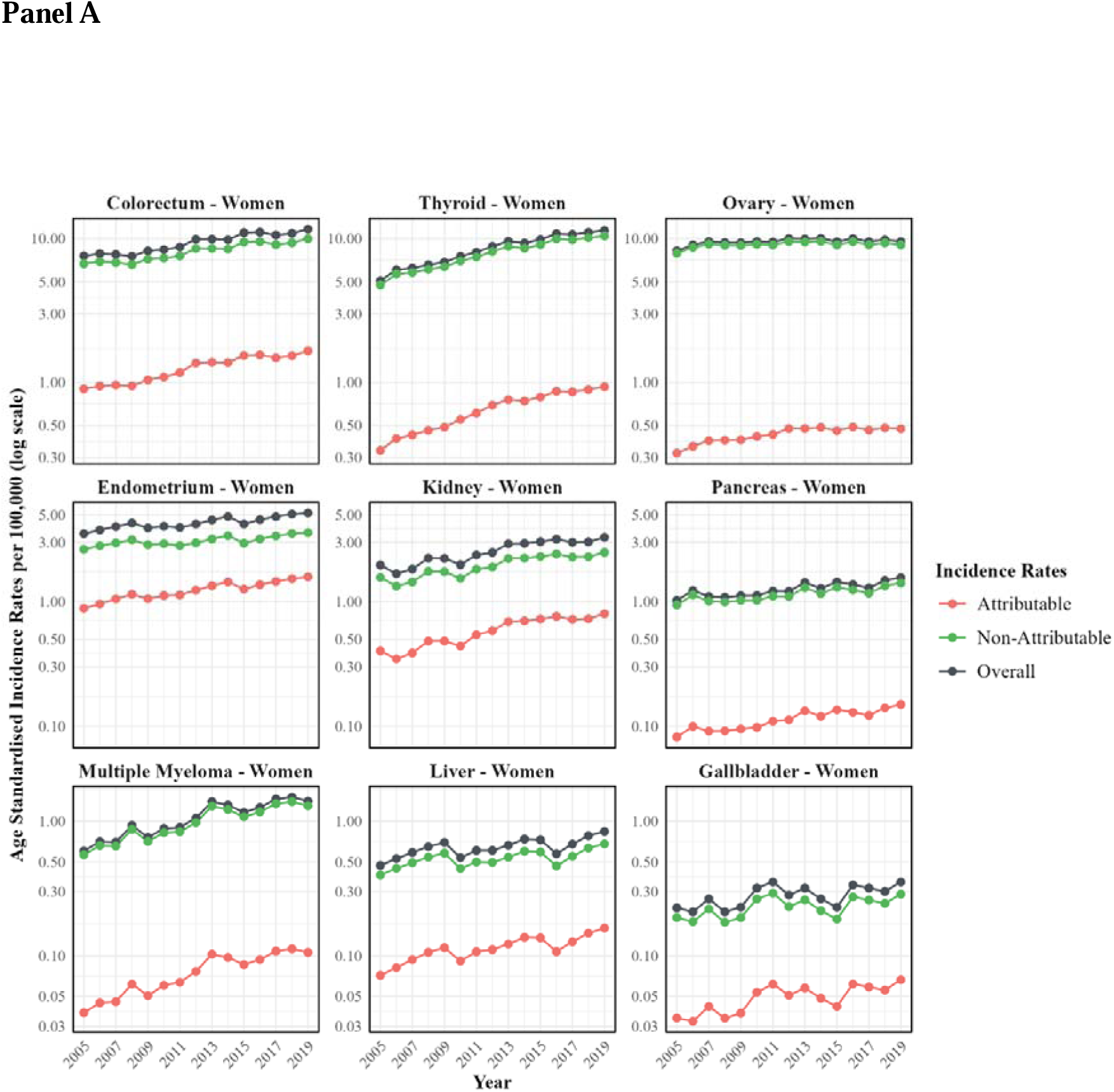
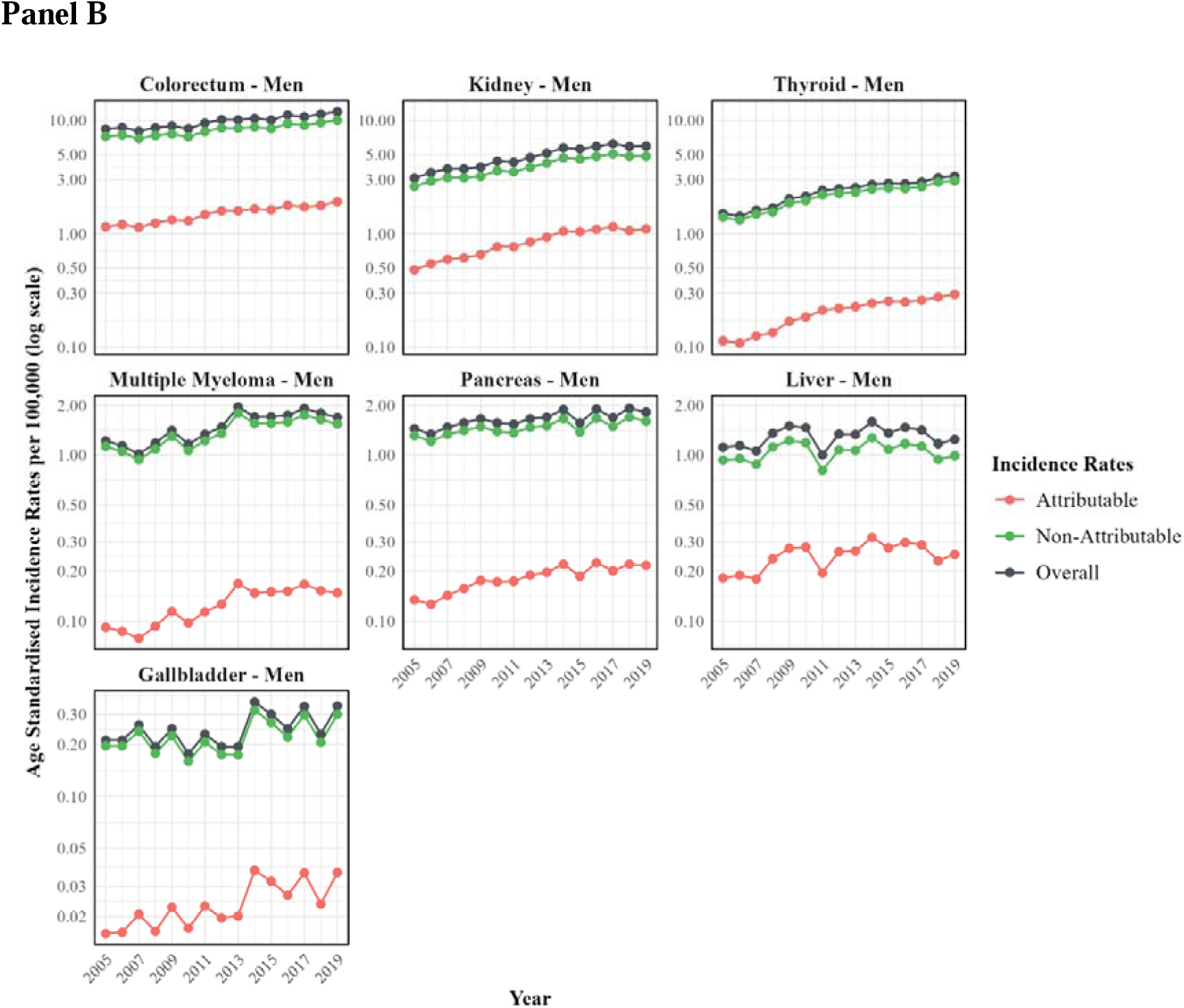
Cancer incidence rates in younger adults (20-49 years) in women (**Panel A**) and men (**Panel B**) for nine BMI-related cancers and partitioned into overall, BMI-attributable, and BMI-non-attributable cancer incidence rates, England 2001-2019. See **Supplementary** Figure 5 for similar figures older adults, and **Supplementary Table 6** for estimates of the most recent Annual Percentage Change (APC) and the Average Annual Percentage Change (AAPC).

## Discussion

Our analysis identified eleven cancers linked to behavioural risk factors with rising incidence in younger adults in England. For most cancers, incidence also increased in older adults at a similar pace, except colorectal and ovarian cancer, which increased only in younger adults, and some cancers that increased faster in younger than older adults. These patterns suggest that while similar risk factors across ages are likely, some cancers may have age-specific exposures, susceptibilities, or differences in screening and detection practices.

Common behavioural risk factors - smoking, alcohol, overweight and obesity, physical inactivity, red and processed meat consumption, and low fibre intake-could explain a substantial fraction of cancers in 2019 in England (e.g.∼ 40-50% of colorectal, endometrium, oral or liver). However, except for obesity, most factors showed stable or declining trends in recent decades. Unless exposure effects are underestimated, have longer lag periods or persistent effects after reductions in exposure (e.g. smoking and lung cancer ^14^), their contribution to increasing trends in cancer incidence is likely limited.

The observed increasing cancer incidence despite declining trends in several behavioural risk factors may reflect the net effect of multiple influences operating in different directions. Other contributing factors not evaluated here, e.g. reproductive history, early-life or prenatal risk factors, and changes in cancer diagnosis and detection practices, may also play a role. Although incidence rates increased overall from 2001-2019, recent stabilising or declining trends for some cancers – e.g. breast and ovarian in women, and kidney, liver and oral cancers in men - may reflect decreasing risk factor trends, for instance declines in smoking and alcohol consumption influencing oral and liver cancer trends. Future cancer site-specific studies are needed to disentangle these complex patterns.

Although overweight and obesity are linked to ten of the eleven cancers and account for a substantial proportion of cancers, both BMI-attributable and non-attributable incidence rates have increased - though the latter more slowly-suggesting other contributors. Given the high prevalence of overweight/obesity, reducing their prevalence could substantially lower incidence (e.g. ∼20-30% for endometrial, kidney, liver and gallbladder cancers in younger women, ∼20% for kidney and liver cancer in younger men, and ∼15% for colorectal cancers in younger men or women). The impact of BMI on incidence trends may be underestimated if the lag times exceed the typically assumed 10-years, or if effect sizes from epidemiological studies are substantially underestimated. Additionally, obesity-related conditions such as diabetes and metabolic syndrome are independently linked to several cancers and may also contribute^15,16^, underscoring the importance of considering metabolic health more broadly.

For a single risk factor to substantially impact changes in cancer incidence at the population level, it must be both common and strongly associated with risk, as illustrated in **Supplementary Figure 6.** In addition, there must also be large shifts in risk factor prevalence over relevant periods of time. Several suspected risk factors - including ultra-processed foods, childhood obesity/physical inactivity, sedentary behaviour, antibiotic use, sweetened beverages, and air pollution - have been proposed as contributors to rising cancer incidence in younger adults. While these factors are common in England, most have shown stable or declining trends in the last decade (**Table 1**). Other emerging factors such as the gut microbiome and potential interactions with diet, obesity, and lifestyle factors in colorectal cancer risk^17^ warrant further study. For example, a somatic sequencing study identified a mutational signature in colorectal tumours linked to colibactin, a mutagen produced by some gut bacteria, which was common and enriched in early-onset cancers and may act early in life ^18^. Evaluating the role of these and other emerging factors in rising incidence trends will require sufficiently powered study designs and accurate exposure measurements over different time periods (e.g. early-life).

**Table 1:**
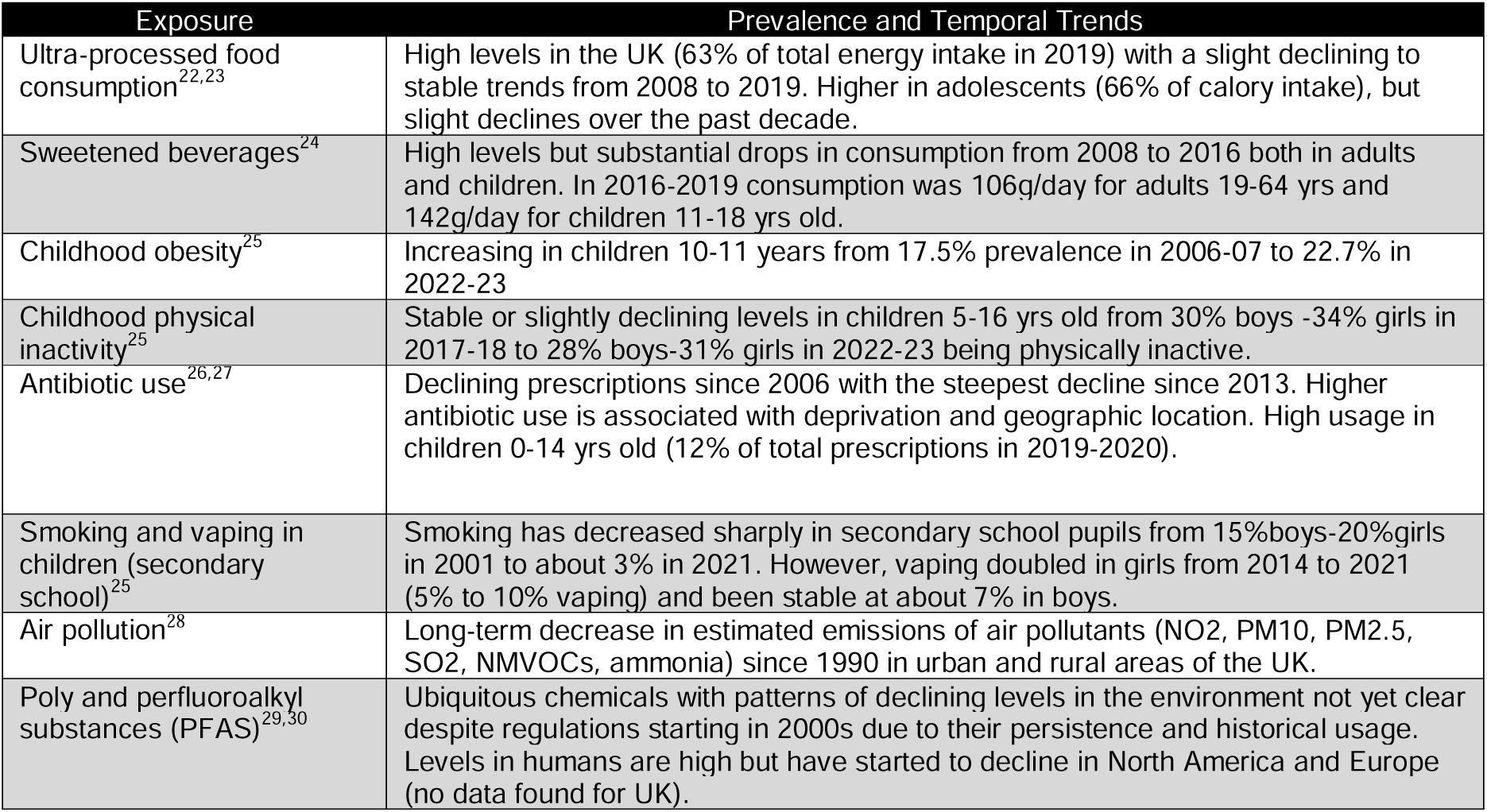
Description of prevalence and temporal trends of exposures suggested to be implicated in rising cancer trends in younger adults

| Exposure | Prevalence and Temporal Trends |
| --- | --- |
| Ultra-processed food consumption<sup>22,23</sup> | High levels in the UK (63% of total energy intake in 2019) with a slight declining to stable trends from 2008 to 2019. Higher in adolescents (66% of calory intake), but slight declines over the past decade. |
| Sweetened beverages<sup>24</sup> | High levels but substantial drops in consumption from 2008 to 2016 both in adults and children. In 2016-2019 consumption was 106g/day for adults 19-64 yrs and 142g/day for children 11-18 yrs old. |
| Childhood obesity<sup>25</sup> | Increasing in children 10-11 years from 17.5% prevalence in 2006-07 to 22.7% in 2022-23 |
| Childhood physical inactivity<sup>25</sup> | Stable or slightly declining levels in children 5-16 yrs old from 30% boys -34% girls in 2017-18 to 28% boys-31% girls in 2022-23 being physically inactive. |
| Antibiotic use<sup>26,27</sup> | Declining prescriptions since 2006 with the steepest decline since 2013. Higher antibiotic use is associated with deprivation and geographic location. High usage in children 0-14 yrs old (12% of total prescriptions in 2019-2020). |
| Smoking and vaping in children (secondary school)<sup>25</sup> | Smoking has decreased sharply in secondary school pupils from 15%boys-20%girls in 2001 to about 3% in 2021. However, vaping doubled in girls from 2014 to 2021 (5% to 10% vaping) and been stable at about 7% in boys. |
| Air pollution<sup>28</sup> | Long-term decrease in estimated emissions of air pollutants (NO2, PM10, PM2.5, SO2, NMVOCs, ammonia) since 1990 in urban and rural areas of the UK. |
| Poly and perfluoroalkyl substances (PFAS)<sup>29,30</sup> | Ubiquitous chemicals with patterns of declining levels in the environment not yet clear despite regulations starting in 2000s due to their persistence and historical usage. Levels in humans are high but have started to decline in North America and Europe (no data found for UK). |

Although cancer incidence is rising in younger adults, rates remain much higher in older adults for most cancers, with thyroid being the main exception (**Figure 1** and **Supplementary Figure 7**). For most cancers, incidence is increasing across both age groups and PAFs are broadly similar, suggesting that common causes likely act across ages thus making studies of populations with wide age ranges most informative. Colorectal cancer is a notable exception, with persistent increases limited to younger adults, pointing to causes unique to younger generations^6^ and the need for studies focused on early-life exposures. The NHS Bowel Cancer Screening Programme, introduced for older adults in the mid-2000s, likely contributed to declines in that group^18^. However, it is unlikely to fully explain the differences by age groups given that rates have been different since early 2000’s. Other preventable cancers rising faster in younger adults—including endometrium, kidney, pancreatic, multiple myeloma and thyroid cancer —also suggest possible sex- or age-specific risk factors or differences in susceptibility that warrant investigation. Thus, while similar patterns in trends across age groups mean that studying older adults can provide insights into causes of cancer in younger populations, focused research on younger adults remains critical, particularly when incidence patterns point to unique generational or life course influences.

Sex differences in incidence rates and PAFs likely reflect differing exposure prevalences in men and women that require monitoring and targeted action. Risk factors also differ by socioeconomic groups: although overall risk factor trends were similar across deprivation groups, the most deprived—defined by the IMD, which reflects income, employment, education, housing, health, crime, and environment—experienced slower declines in smoking, physical inactivity, and alcohol use, alongside faster rises in obesity, and consistently higher exposure prevalence than the least deprived. Together, these sex and socioeconomic disparities contribute to unequal cancer burdens and underscore the need to account for both demographic and contextual factors when developing strategies to reduce health inequities.

A limitation of our analyses is the lack of consistent, long-term national data for several risk factors. Physical inactivity data, for example, was available only for five timepoints between 2003 and 2012, when the UK guidelines’ definition of inactivity changed substantially. This restricted our ability to evaluate long-term exposure trends beyond one to two decades, depending on the risk factor. Our analysis focused on England because of its size and readiness of survey data-future analyses are needed for other geographical regions in the UK. Another limitation is the assumption of a 10-year lag period between exposure and cancer incidence. While this might be reasonable for many cancers, we were unable to evaluate longer lag times due to limited exposure data. This limitation also applies to other studies that have suggested that these risk factors could explain increases in cancer incidence without formal evaluation. Our analysis makes the issue more explicit. **Supplementary Figure 8** illustrates this using an example of early-onset colorectal cancer showing available data on incidence trends and risk factor prevalence with a 10-year time lag.

Self-reported surveys are prone to measurement error in exposures, such as underreporting alcohol or overreporting physical activity with rising health awareness. However, the use of standardized, validated questionnaires in national surveys minimizes differential bias across years. Using exposure surrogates, e.g. BMI for adiposity, or broad categories like ultra-processed foods could underestimate the impact of underlying exposures. More precise measures, such as organ-specific fat deposition or finer food classifications, could improve risk estimation and assessment of the impact of risk factors on incidence trends. Another limitation was the lack of RR estimates consistently reported by age and sex groups. Therefore, we only consider age-specific RR for the well-established example of obesity and breast cancer risk, which has an inverse risk association in younger women and a direct association in older women^19^. Additionally, while we evaluated the independent impact of each risk factor on cancer incidence thus avoiding double counting in aggregated PAF estimates, multiple weakly associated exposures acting together could create cumulative effects that are difficult to quantify.

Changes in detection and diagnostic practices such as population-wide cancer screening programs, increased used of diagnostic tests or advanced imaging technologies and broadening of disease definitions could also have played a role in the observed cancer incidence trends^20^. For instance, in the UK there is evidence for changes in practice and possible overdiagnosis of cancers of oral, endometrial, thyroid, kidney and breast cancers which were found to be increasing^21^. Future analyses of incidence trends by stage at diagnosis or accounting for overdiagnosis could help clarify the impact of surveillance and screening.

A strength of this report is the systematic evaluation of both cancer incidence and risk factor trends in younger and older adults, using a formal approach to estimate the contribution of exposures changes to cancer incidence. While analyses are descriptive in nature and preclude causal inference, these ecological comparisons provide valuable context for assessing whether previously proposed explanations for rising incidence trends are consistent with population-level patterns. Although not exhaustive, our examples provide useful benchmarks for assessing whether a given risk factor could plausibly explain a substantial proportion of cancer cases.

In conclusion, the incidence of several cancers is increasing in both younger and older adults in England. Apart from BMI, established behavioural risk factors do not seem likely contributors for the increase in early-onset cancers, with incidence rates increasing despite favourable trends in several known risk factors. This underscores the influence of obesity and other risk factors for the increases in younger and older adults. Further research into emerging and interacting exposures, improved measurement, and continued surveillance is needed, alongside evaluation of the impact of screening and shifts in stage at diagnosis. Finally, although increases in cancer in younger adults are concerning, the absolute burden remains far higher in older adults, underscoring the public health and clinical importance of studying risk factors across all ages.

### Ethics approval

This study used de-identified, routinely collected cancer registry and national health survey data. Ethical approval was not required for this analysis as per guidance from the Health Research Authority (HRA) and the UK Policy Framework for Health and Social Care Research. All data were accessed and analysed in accordance with the data providers’ governance frameworks and approvals.

### Transparency declaration

The lead author affirms that this manuscript is an honest, accurate, and transparent account of the study being reported; that no important aspects of the study have been omitted; and that any discrepancies from the study as planned (and, if relevant, registered) have been explained.

## Acknowledgements

The authors would like to thank the UKDS and the NDRS for collating, maintaining, and quality ensuring data from nationally representative surveys and the NHS, respectively. They are both important data resources of large accessible datasets which are vital to this analysis.

## Authors’ Contributions

MGC, MJG, ABdeG conceived the study. MGC and ABdeG provided overall direction for the work. MGC drafted the manuscript. ZR and RF obtained the data sources, conducted statistical analyses and contributed to drafting the Methods section. All authors contributed to the interpretation of results and critically revised the manuscript.

## Ethics approval and consent to participate

This study used de-identified, routinely collected cancer registry and national health survey data. Ethical approval was not required for this analysis as per guidance from the Health Research Authority (HRA) and the UK Policy Framework for Health and Social Care Research. All data were accessed and analysed in accordance with the data providers’ governance frameworks and approvals. The study was conducted in accordance with the principles of the Declaration of Helsinki.

## Data availability

All publicly available datasets used in this analysis can be found on <u>GitHub</u>. Individual level data on risk factors can be obtained through an End User License Agreement with the UK Data Service (UKDS). Links to all used datasets are available in **Supplementary Table 1**.

