## Supplementary Materials for "Temporal Trends in Behavioural Risk Factors for Cancers with Rising Incidence in Younger Adults: An Analysis of Population-Based Data in England"

### Supplementary Methods

#### Data Sources

*Cancer Incidence Data*

Age-specific cancer incidence data was obtained from the National Disease Registration Service (NDRS) for England from 2001 to 2019 (**Supplementary Table 1**), stratified by 5-year age groups and sex. The 2019 cutoff was selected to avoid distortions in incidence trends caused by diagnostic and reporting changes during the COVID-19 pandemic. The starting year 2001 was the earliest with data available for download from the NDRS. Age-standardised cancer incidence rates for the age groups 20-49 and 50+ were then derived using the weights corresponding to the 2013 European standardised population[1]. For cancer site selection, we used the ICD groupings utilised by Globocan in their "Cancer Over Time" project including 28 cancer sites[2] . From this list, we combined colon and rectal cancers of their similar aetiologies and rarity of rectal cancer and separated cervix uteri and cervix corpus into cervix and endometrium, respectively, because of their distinct aetiologies. This resulted in a total of 25 cancer sites to be evaluated.

*Risk Factor Data*

We evaluated temporal trends for the following behavioural risk factors: cigarette smoking, elevated body mass index (BMI), alcohol consumption, red and processed meat consumption, low fibre intake, and physical inactivity. These factors were selected because they are established risk factors for the subset of cancer sites with statistically significant increases in incidence in younger adults between 2001 and 2019 in our analyses (see statistical analyses below). Established risk factors were defined as those classified as Group 1 carcinogens by the International Agency for Research on Cancer (IARC); or having strong evidence for cancer risk associations by the World Cancer Research Fund (WCRF) (**Supplementary Table 2**). Data on these risk factors was obtained from population-based surveys in England, covering the longest available timeframe from 1995 until 2019 (**Supplementary Table 1**). Data availability varied by risk factor due to changes in definitions and measurement over time: the widest range of exposure was for smoking (1995-2019) and the shortest for physical activity (2003-2012). Risk factor exposure levels were defined using pre-existing cutoffs (**Supplementary Table 3**). To smooth trends in risk factors, we applied a moving average with a period length of 3 years to the collected survey data. All prevalences quoted in the manuscript or used in analyses are running averages.

#### Statistical Analyses

*Cancer Incidence and Risk Factor Trends*

Temporal trends in cancer incidence rates and risk factor prevalence were visualised using time series plots on a log scale to reflect proportional differences over time. To quantify changes, we estimated Average Annual Percentage Change (AAPC) from 2001 to 2019, and the most recent Annual Percentage Change (APC) with corresponding 95% confidence intervals (CI) using Joinpoint regression[3] AAPC and APC were derived from a log-linear model to reflect the annual relative change in incidence or risk factor prevalence (i.e. percent change relative to the previous year). AAPC was estimated for 25 cancer sites by age group (ages 20-49 and ≥50) and sex (men and women). Depending on the number of joins in the final selected model, a z-test or t-test was conducted to test if AAPC was different from zero using an alpha-level of 0.05. The APC of the most recent trend is presented as an estimated of the most recent relative changes in incidence of cancers with significant and positive AAPCs

(2001-2019) in younger adults, which also met the inclusion criteria for having associations with behavioural risk factors (as defined above). Joinpoint regression was applied to serial data on risk factors over the available timeframe to estimate AAPCs for relative changes in risk factor prevalence by age group, sex, and index of multiple deprivation (IMD) category[4]. All AAPC and APC calculations were done in JoinPoint software[5].

*Population Attributable Fractions (PAF)*
We estimated the Population Attributable Fraction (PAF), i.e. the proportion of cancer cases attributable to a given risk factor in the population.
PAFs were calculated by age and sex groups according to the following formula:

$$PAF = \frac{\sum p_n(RR_n - 1)}{1 + \sum p_n(RR_n - 1)}$$

Where:
$p_n$ = Population prevalence for the $n^{th}$ level of the risk factor
$RR_n$ = Relative Risk for the $n^{th}$ level of the risk factor
Risk factor RR for selected cancers were identified using a literature review, following the same methodology as in Brown et al[6]. Briefly, cancer site and risk factor combinations were searched in PubMed using AND functions between each combination (**Supplementary Table 7**). PubMed searches were supplemented by reference lists in relevant papers. Preference for RR extraction was given to papers that conducted prospective pooled analysis. For colorectal cancer, RR for colon cancer was extracted for physical activity as it is the more prevalent cancer subtype, and the literature was limited due to changes in physical activity measurement over time.
For risk factors with no pre-established exposure categories (e.g. red meat, processed meat and fiber intake), continuous measures of consumption in grams were grouped into exposure categories, and corresponding category RRs were calculated for the consumption midpoint. For factors with reported RR <1.0 (i.e. fiber intake, physical activity) we calculated the RR for the risk category (i.e. absence of the protective factor) as (1 + ln(1/RR))[6].
PAFs aggregated across all risk factors for a given cancer site assume independence between risk factors, and were calculated by combining factor specific PAFs, applying each additional risk factor to only the remaining cancer cases not attributed to a previous exposure. To account for the delay between risk factor exposure and cancer diagnosis, a 10-year time lag was applied in estimating attributable cases[6–8]. When risk factor data was not available for 2009, the closest year with data available was chosen.
Monte Carlo simulation methods were used to estimate 95% CIs for the PAF estimates. For each sample, RR values were drawn from a log normal distribution with variance inferred from reported CIs, and risk factor prevalences were sampled from a multinomial distribution *Multinom(n, p)*, where n is the number of survey participants, and p is the survey prevalences. To account for the smoothing applied to the survey data, the prevalences were sampled for the survey years directly before and after the survey year of interest, and the prevalences were averaged to recreate the moving average. We simulated individual PAFs 1000 times and used the resulting 2.5% and 97.5% quantiles of PAFs to define 95% CIs.

*Attributable and Non-Attributable Cancer Incidence Rates*
Disaggregated cancer incidence rates attributable and non-attributable to a risk factor were calculated for risk factors with increasing temporal trends in younger adults (defined as AAPC were statistically significantly different from 0 and increasing in the 20-49 age group in men or women). For categorical risk factors (i.e. smoking, alcohol, BMI), only the highest exposure level AAPC were considered to determine if exposure was increasing (i.e. current

smokers, heavy drinkers, and obese). Disaggregated incidence rates were calculated across the available periods, i.e. from the first year with available risk factor data is to 2019, by calculating PAFs for each year and multiplying it by the cancer incidence rate for the corresponding year. AAPC were calculated using Joinpoint regression software to estimate the relative change in attributable and non-attributable cancer incidence across the available period[5].

**Code and data accessibility**

All code and publicly available datasets used in this analysis can be found on GitHub. Individual level data on risk factors can be obtained through an End User License Agreement with the UK Data Service (UKDS). Links to all used datasets are available in **Supplementary Table 1.**

### Supplementary Tables

**Supplementary Table 1**: Cancer incidence and risk factor data availability and sourcing information

| Dataset | Year(s) | Date Last Accessed | Source | Publicly Available | Total Number of Participants | Cancer Incidence | Smoking Status | Alcohol Usage | BMI | Physical Activity | Daily Diet |
|---|---|---|---|---|---|---|---|---|---|---|---|
| National Disease Registration Service (Age Groups: 20-49, 50+) | 2001-2019 | 07/04/2025 | NDRS | Yes | - | ✓ | - | - | - | - | - |
| Health Survey for England | 1995 | 30/10/2024 | UKDS | No | 19788 | - | ✓ | - | ✓ | - | - |
| Health Survey for England | 1996 | 30/10/2024 | UKDS | No | 20328 | - | ✓ | - | ✓ | - | - |
| Health Survey for England | 1997 | 30/10/2024 | UKDS | No | 15546 | - | ✓ | - | ✓ | - | - |
| Health Survey for England | 1998 | 30/10/2024 | UKDS | No | 19654 | - | ✓ | - | ✓ | - | - |
| Health Survey for England | 1999 | 30/10/2024 | UKDS | No | 9640 | - | ✓ | - | ✓ | - | - |
| Health Survey for England | 2000 | 30/10/2024 | UKDS | No | 12413 | - | ✓ | - | ✓ | - | - |
| Health Survey for England | 2001 | 30/10/2024 | UKDS | No | 19640 | - | ✓ | - | ✓ | - | - |
| Health Survey for England | 2002 | 30/10/2024 | UKDS | No | 18396 | - | ✓ | - | ✓ | - | - |
| Health Survey for England | 2003 | 30/10/2024 | UKDS | No | 18553 | - | ✓ | - | ✓ | ✓ | - |
| Health Survey for England | 2004 | 31/10/2024 | UKDS | No | 8354 | - | ✓ | - | ✓ | ✓ | - |
| Health Survey for England | 2005 | 30/10/2024 | UKDS | No | 13297 | - | ✓ | - | ✓ | - | - |
| Health Survey for England | 2006 | 30/10/2024 | UKDS | No | 21399 | - | ✓ | - | ✓ | ✓ | - |
| Health Survey for England | 2007 | 30/10/2024 | UKDS | No | 14386 | - | ✓ | - | ✓ | - | - |
| Health Survey for England | 2008 | 30/10/2024 | UKDS | No | 22619 | - | ✓ | - | ✓ | ✓ | - |
| Health Survey for England | 2009 | 30/10/2024 | UKDS | No | 8602 | - | ✓ | - | ✓ | - | - |
| Health Survey for England | 2010 | 30/10/2024 | UKDS | No | 14112 | - | ✓ | - | ✓ | - | - |
| Health Survey for England | 2011 | 30/10/2024 | UKDS | No | 10617 | - | ✓ | ✓ | ✓ | - | - |
| Health Survey for England | 2012 | 30/10/2024 | UKDS | No | 10333 | - | ✓ | ✓ | ✓ | ✓ | - |
| Health Survey for England | 2013 | 30/10/2024 | UKDS | No | 10980 | - | ✓ | ✓ | ✓ | - | - |
| Health Survey for England | 2014 | 30/10/2024 | UKDS | No | 10080 | - | ✓ | ✓ | ✓ | - | - |

| | | | | | | | | | | | |
|---|---|---|---|---|---|---|---|---|---|---|---|
| Health Survey for England | 2015 | 30/10/2024 | UKDS | No | 13748 | - | ✓ | ✓ | ✓ | - | - |
| Health Survey for England | 2016 | 30/10/2024 | UKDS | No | 10067 | - | ✓ | ✓ | ✓ | ✓ | - |
| Health Survey for England | 2017 | 30/10/2024 | UKDS | No | 9982 | - | ✓ | ✓ | ✓ | - | - |
| Health Survey for England | 2018 | 30/10/2024 | UKDS | No | 10250 | - | ✓ | ✓ | ✓ | - | - |
| Health Survey for England | 2019 | 30/10/2024 | UKDS | No | 10299 | - | ✓ | ✓ | ✓ | - | - |
| General Household Survey | 2005 | 04/11/2024 | UKDS | No | 30069 | - | - | ✓ | - | - | - |
| National Diet and Nutrition Survey | 2008 | 06/11/2024 | UKDS | No | 1646 | - | - | - | - | - | ✓ |
| National Diet and Nutrition Survey | 2009 | 06/11/2024 | UKDS | No | 1669 | - | - | - | - | - | ✓ |
| National Diet and Nutrition Survey | 2010 | 06/11/2024 | UKDS | No | 1565 | - | - | - | - | - | ✓ |
| National Diet and Nutrition Survey | 2011 | 06/11/2024 | UKDS | No | 1948 | - | - | - | - | - | ✓ |
| National Diet and Nutrition Survey | 2012 | 06/11/2024 | UKDS | No | 1197 | - | - | - | - | - | ✓ |
| National Diet and Nutrition Survey | 2013 | 06/11/2024 | UKDS | No | 1349 | - | - | - | - | - | ✓ |
| National Diet and Nutrition Survey | 2014 | 06/11/2024 | UKDS | No | 1353 | - | - | - | - | - | ✓ |
| National Diet and Nutrition Survey | 2015 | 06/11/2024 | UKDS | No | 1370 | - | - | - | - | - | ✓ |
| National Diet and Nutrition Survey | 2016 | 06/11/2024 | UKDS | No | 1253 | - | - | - | - | - | ✓ |
| National Diet and Nutrition Survey | 2017 | 06/11/2024 | UKDS | No | 1211 | - | - | - | - | - | ✓ |
| National Diet and Nutrition Survey | 2018 | 06/11/2024 | UKDS | No | 1094 | - | - | - | - | - | ✓ |

**Supplementary Table 2**: Source of evidence to identify behavioural risk factors causally associated with 11 selected cancer sites (oral, endometrial, pancreatic, gallbladder, colorectal, liver, kidney, thyroid, multiple myeloma, and breast cancers).

| Exposure | Exposure Categories | Decision Source | Date Last Accessed |
|---|---|---|---|
| Smoking | Current Smoking | IARC[9] | 11/08/2025 |
| Smoking | Former Smoking | IARC[9] | 11/08/2025 |
| Alcohol | Light (<12 g/day) Drinking | IARC, WCRF[9,10] | 21/02/2025 |
| Alcohol | Medium (12-50 g/day) Drinking | IARC, WCRF[9,10] | 21/02/2025 |
| Alcohol | Heavy (>50 g/day) Drinking | IARC, WCRF[9,10] | 21/02/2025 |
| BMI | Overweight (25-30 kg/m$^2$) | IARC, WCRF[9,11] | 11/08/2025 |
| BMI | Obese (>30 kg/m$^2$ ) | IARC, WCRF[9,11] | 11/08/2025 |
| Physical Activity | Being active less than the 150 minutes per week | WCRF[12] | 21/02/2025 |
| Processed Meat | Any consumption increases risk | IARC, WCRF[9,13] | 21/02/2025 |
| Red Meat | Any consumption increases risk | WCRF[13] | 21/02/2025 |
| Fibre | Consuming less than the UK guideline of 30g/day | WCRF[14] | 21/02/2025 |

**Supplementary Table 3**: Relative risks for the association between established behavioural risk factors and 11 selected cancer sites, used in population attributable fraction (PAF) calculations

| Risk Factors | Oral | Endometrium | Pancreas | Gallbladder | Colorectum | Liver | Kidney | Thyroid | Multiple Myeloma | Breast | Ovary |
|---|---|---|---|---|---|---|---|---|---|---|---|
| Smoking[15–20] | | | | | | | | | | | |
| Current Smoking | | | | | | | | | | | |
| Men | 3.43 (2.37, 4.94) [B] | - | 2.2 (1.71, 2.83) | - | 1.19 (1.11,1.21) [B] | 1.61 (1.38, 1.89) | 1.39 (1.28, 1.51) [B] | - | - | - | - |
| Women | 3.43 (2.37, 4.94) [B] | - | 2.2 (1.71, 2.83) | - | 1.17 (1.09, 1.25) [B] | 1.86 (1.33, 2.60) | 1.39 (1.28, 1.51) [B] | - | - | - | 1.06 (1.00, 1.13)[B] |
| Former Smoking | | | | | | | | | | | |
| Men | 1.4 (0.99, 2.00) [B] | - | 1.17 (1.02, 1.34) | - | 1.22 (1.18, 1.26) [B] | 1.47 (1.19, 1.82) | 1.2 (1.14, 1.27) [B] | - | - | - | - |
| Women | 1.4 (0.99, 2.00) [B] | - | 1.17 (1.02, 1.34) | - | 1.16 (1.13, 1.19) [B] | 1.45 (0.80, 2.65) [B] | 1.2 (1.14, 1.27) [B] | - | - | - | 1.06 1.00, 1.12)[B] |
| Alcohol[21] | | | | | | | | | | | |
| Light (<12 g/day) Drinking | | | | | | | | | | | |
| Men | 1.2 (1.06, 1.35) [B] | - | - | - | 1.05 (0.95, 1.16) [B] | 1.05 (0.84, 1.32) [B] | - | - | - | - | - |
| Women | 1 (0.78, 1.27) [B] | - | - | - | 0.95 (0.89, 1.01) [B] | 0.81 (0.59, 1.12) [B] | - | - | - | 1.06 (1.03, 1.10) [B] | - |
| Medium (12-50 g/day) Drinking | | | | | | | | | | | |
| Men | 2.01 (1.69, 2.40) [B] | - | - | - | 1.21 (1.11, 1.32) [B] | 1.08 (0.88, 1.32) [B] | - | - | - | - | - |
| Women | 1.67 (1.25, 2.22) [B] | - | - | - | 1.07 (0.99, 1.16) [B] | 1.24 (0.88, 1.75) [B] | - | - | - | 1.22 (1.17, 1.27) [B] | - |
| Heavy (>50 g/day) Drinking | | | | | | | | | | | |
| Men | 5.33 | - | - | - | 1.53 | 1.59 | - | - | - | - | - |

| | | | | | | | | | | | |
|---|---|---|---|---|---|---|---|---|---|---|---|
| | (4.28, 6.63)[B] | | | | (1.30, 1.80)[B] | (1.21, 2.09)[B] | | | | | |
| Women | 5.7 (3.75, 8.66)[B] | - | - | - | 1.24 (0.68, 2.25)[B] | 3.89 (1.60, 9.48)[B] | - | - | - | 1.5 (1.19, 1.89 )[B] | - |
| BMI[22–30] | | | | | | | | | | | |
| Overweight (25-30 kg/m$^2$) | | | | | | | | | | | |
| Men | - | - | 1.09 (1.02, 1.16)[B] | 1.01 (0.92, 1.12) | 1.22 (1.10, 1.36)[B] | 1.18 (1.06, 1.31) | 1.22 (1.17, 1.28) | 1.1 (1.03, 1.18) | 1.12 (1.07, 1.18)[B] | - | - |
| Women | - | 1.34 (1.20, 1.48) | 1.15 (1.03, 1.29)[B] | 1.22 (1.09, 1.37) | 1.22 (1.10, 1.36)[B] | 1.18 (1.06, 1.31) | 1.38 (1.29, 1.47) | 1.1 (1.03, 1.18) | 1.12 (1.07, 1.18)[B] | 1.10 (1.06, 1.13)[B, C] | 1.05 (1.00, 1.09)[B] |
| Obese (>30 kg/m$^2$) | | | | | | | | | | | |
| Men | - | - | 1.45 (1.21, 1.75)[B] | 1.54 (1.25, 1.89) | 1.46 (1.24, 1.73)[B] | 1.83 (1.59, 2.11) | 1.63 (1.50, 1.77) | 1.27 (1.15, 1.40) | 1.21 (1.08, 1.35)[B] | - | - |
| Women | - | 2.54 (2.27, 2.81) | 1.28 (1.07, 1.54)[B] | 1.75 (1.44, 2.14) | 1.46 (1.24, 1.73)[B] | 1.83 (1.59, 2.11) | 1.95 (1.81, 2.10) | 1.27 (1.15, 1.40) | 1.21 (1.08, 1.35)[B] | 1.18 (1.12, 1.25)[B,C] | 1.17 (1.14, 1.21)[B] |
| Physical Activity[31] | | | | | | | | | | | |
| Meeting physical activity guidelines (150 min/week) | | | | | | | | | | | |
| Men | - | - | - | - | 0.95 (0.87, 0.99)[B, D] | - | - | - | - | - | - |
| Women | - | 0.9 (0.84, 0.97) | - | - | 0.95 (0.87, 0.99)[B, D] | - | - | - | - | 0.95 (0.92, 0.97) | - |
| Red Meat[32] | | | | | | | | | | | |
| Consumption per 100g/day | | | | | | | | | | | |
| Men | - | - | - | - | 1.12 (1.00, 1.25)[B] | - | - | - | - | - | - |
| Women | - | - | - | - | 1.12 (1.00, 1.25)[B] | - | - | - | - | - | - |
| Processed Meat[32] | | | | | | | | | | | |
| Consumption per 50 g/day | | | | | | | | | | | |
| Men | - | - | - | - | 1.16 | - | - | - | - | - | - |

| | | | | | | | | | | | |
|---|---|---|---|---|---|---|---|---|---|---|---|
| | | | | | (1.10, 1.28) [B] | | | | | | |
| Women | - | - | - | - | 1.16<br>(1.10, 1.28) B | - | - | - | - | - | - |
| Fibre[14] | | | | | | | | | | | |
| Consumption per 10g/day | | | | | | | | | | | |
| Men | - | - | - | - | 0.93<br>(0.87, 1.00) [B] | - | - | - | - | - | - |
| Women | - | - | - | - | 0.93<br>(0.87, 1.00) [B] | - | - | - | - | - | - |

[A] Relative risk factors were selected if they are behavioural risk factors that are causally associated with the cancer sites of interest as defined by IARC (group 1) or WCRF (strong evidence).

[B] Relative risks updated from Brown et al, 2018 paper using its own literature search methodology.

[C] This is for the women ages 50+ only as it only applies to postmenopausal breast cancer. All other relative risks are assumed to be constant across age groups.

[D] Relative risk for colon as it is the most common subtype of colorectal cancer.

**Supplementary Table 4**: Joinpoint average annual percentage change (AAPC) and most recent annual percentage change (APC) for cancer incidence trends in 11 selected cancer sites by age group for women (**Panel A**) and men (**Panel B**)

**Panel A**

| Age Group | AAPC | | | | | Most Recent APC | | | |
|---|---|---|---|---|---|---|---|---|---|
| | Time Period | AAPC[1] | CI 95% | P Value | P Value Difference | Time Period | APC[2] | CI 95% | P Value |
| **Breast** | | | | | | | | | |
| 20-49 | (2001, 2019) | 0.88 | (0.56, 1.21) | <0.0001 | - | (2013, 2019) | 0.21 | (-0.67, 1.09) | 0.62 |
| 50+ | (2001, 2019) | 0.70 | (0.17, 1.23) | 0.01 | 0.28 | (2003, 2019) | 0.42 | (0.22, 0.61) | 0.00037 |
| **Colorectum** | | | | | | | | | |
| 20-49 | (2001, 2019) | 3.20 | (2.78, 3.62) | <0.0001 | - | (2001, 2019) | 3.20 | (2.78, 3.62) | <0.0001 |
| 50+ | (2001, 2019) | 0.36 | (-0.13, 0.85) | 0.15 | <0.0001 | (2017, 2019) | 2.01 | (-1.88, 6.06) | 0.28 |
| **Endometrium** | | | | | | | | | |
| 20-49 | (2001, 2019) | 2.77 | (2.22, 3.32) | <0.0001 | - | (2001, 2019) | 2.77 | (2.22, 3.32) | <0.0001 |
| 50+ | (2001, 2019) | 1.80 | (1.46, 2.14) | <0.0001 | 0.0027 | (2011, 2019) | 0.29 | (-0.33, 0.91) | 0.34 |
| **Gallbladder** | | | | | | | | | |
| 20-49 | (2001, 2019) | 3.21 | (1.86, 4.58) | <0.0001 | - | (2001, 2019) | 3.21 | (1.86, 4.58) | <0.0001 |
| 50+ | (2001, 2019) | 3.38 | (2.95, 3.81) | <0.0001 | 0.59 | (2001, 2019) | 3.38 | (2.95, 3.81) | <0.0001 |
| **Kidney** | | | | | | | | | |
| 20-49 | (2001, 2019) | 4.53 | (3.82, 5.25) | <0.0001 | - | (2001, 2019) | 4.53 | (3.82, 5.25) | <0.0001 |
| 50+ | (2001, 2019) | 3.04 | (2.42, 3.66) | <0.0001 | 0.0012 | (2014, 2019) | -0.65 | (-2.64, 1.38) | 0.5 |
| **Liver** | | | | | | | | | |
| 20-49 | (2001, 2019) | 5.08 | (1.61, 8.66) | 0.0038 | - | (2003, 2019) | 2.89 | (1.66, 4.15) | 0.00018 |
| 50+ | (2001, 2019) | 3.39 | (1.72, 5.10) | <0.0001 | 0.2 | (2013, 2019) | 0.70 | (-1.59, 3.05) | 0.52 |
| **Multiple Myeloma** | | | | | | | | | |

| Age Group | | AAPC | | | | | Most Recent APC | | |
|---|---|---|---|---|---|---|---|---|---|
| | Time Period | AAPC[1] | CI 95% | P Value | P Value Difference | Time Period | APC[2] | CI 95% | P Value |
| 20-49 | (2001, 2019) | 6.02 | (4.93, 7.13) | <0.0001 | - | (2001, 2019) | 6.02 | (4.93, 7.13) | <0.0001 |
| 50+ | (2001, 2019) | 2.62 | (2.13, 3.11) | <0.0001 | <0.0001 | (2001, 2019) | 2.62 | (2.13, 3.11) | <0.0001 |
| **Oral** | | | | | | | | | |
| 20-49 | (2001, 2019) | 2.15 | (1.50, 2.80) | <0.0001 | - | (2001, 2019) | 2.15 | (1.50, 2.80) | <0.0001 |
| 50+ | (2001, 2019) | 2.70 | (1.48, 3.93) | <0.0001 | 0.79 | (2010, 2019) | 1.61 | (0.93, 2.30) | 0.00028 |
| **Ovary** | | | | | | | | | |
| 20-49 | (2001, 2019) | 0.72 | (0.05, 1.39) | 0.036 | - | (2013, 2019) | -0.57 | (-2.33, 1.23) | 0.51 |
| 50+ | (2001, 2019) | -1.31 | (-1.51, -1.10) | <0.0001 | <0.0001 | (2001, 2019) | -1.31 | (-1.51, -1.10) | <0.0001 |
| **Pancreas** | | | | | | | | | |
| 20-49 | (2001, 2019) | 2.63 | (1.86, 3.41) | <0.0001 | - | (2001, 2019) | 2.63 | (1.86, 3.41) | <0.0001 |
| 50+ | (2001, 2019) | 0.98 | (0.67, 1.28) | <0.0001 | <0.0001 | (2013, 2019) | -0.09 | (-0.89, 0.72) | 0.81 |
| **Thyroid** | | | | | | | | | |
| 20-49 | (2001, 2019) | 6.12 | (5.51, 6.73) | <0.0001 | - | (2013, 2019) | 3.27 | (1.69, 4.88) | 0.00053 |
| 50+ | (2001, 2019) | 4.77 | (3.94, 5.60) | <0.0001 | 0.0051 | (2014, 2019) | 2.36 | (-0.32, 5.12) | 0.081 |

[1]Average Annual Percentage Change

[2]Annual Percentage Change

**Panel B**

| Age Group | AAPC | | | | | Most Recent APC | | | |
|---|---|---|---|---|---|---|---|---|---|
| | Time Period | AAPC[1] | CI 95% | P Value | P Value Difference | Time Period | APC[2] | CI 95% | P Value |
| **Colorectum** | | | | | | | | | |
| 20-49 | (2001, 2019) | 2.60 | (2.26, 2.93) | <0.0001 | - | (2001, 2019) | 2.60 | (2.26, 2.93) | <0.0001 |
| 50+ | (2001, 2019) | -0.21 | (-0.79, 0.37) | 0.48 | <0.0001 | (2014, 2019) | -0.58 | (-1.37, 0.21) | 0.13 |
| **Gallbladder** | | | | | | | | | |
| 20-49 | (2001, 2019) | 5.87 | (-0.68, 12.86) | 0.08 | - | (2003, 2019) | 2.29 | (-0.05, 4.68) | 0.054 |
| 50+ | (2001, 2019) | 2.68 | (2.08, 3.29) | <0.0001 | 0.17 | (2001, 2019) | 2.68 | (2.08, 3.29) | <0.0001 |
| **Kidney** | | | | | | | | | |
| 20-49 | (2001, 2019) | 3.69 | (2.18, 5.22) | <0.0001 | - | (2016, 2019) | -0.67 | (-7.32, 6.46) | 0.84 |
| 50+ | (2001, 2019) | 2.79 | (2.10, 3.49) | <0.0001 | 0.15 | (2014, 2019) | 0.05 | (-2.19, 2.34) | 0.96 |
| **Liver** | | | | | | | | | |
| 20-49 | (2001, 2019) | 1.47 | (-0.93, 3.93) | 0.23 | - | (2014, 2019) | -3.52 | (-10.98, 4.56) | 0.36 |
| 50+ | (2001, 2019) | 4.52 | (3.86, 5.20) | <0.0001 | >0.99 | (2014, 2019) | 1.49 | (-0.68, 3.70) | 0.16 |
| **Multiple Myeloma** | | | | | | | | | |
| 20-49 | (2001, 2019) | 4.07 | (2.99, 5.16) | <0.0001 | - | (2001, 2019) | 4.07 | (2.99, 5.16) | <0.0001 |
| 50+ | (2001, 2019) | 2.58 | (2.15, 3.02) | <0.0001 | 0.0077 | (2001, 2019) | 2.58 | (2.15, 3.02) | <0.0001 |
| **Oral** | | | | | | | | | |
| 20-49 | (2001, 2019) | 1.27 | (0.85, 1.70) | <0.0001 | - | (2013, 2019) | -1.07 | (-2.17, 0.05) | 0.059 |
| 50+ | (2001, 2019) | 2.92 | (2.27, 3.58) | <0.0001 | >0.99 | (2013, 2019) | 1.75 | (0.03, 3.50) | 0.047 |
| **Pancreas** | | | | | | | | | |
| 20-49 | (2001, 2019) | 1.13 | (-0.55, 2.84) | 0.19 | - | (2004, 2019) | 2.34 | (1.47, 3.22) | <0.0001 |
| 50+ | (2001, 2019) | 1.06 | (0.88, 1.24) | <0.0001 | 0.47 | (2001, 2019) | 1.06 | (0.88, 1.24) | <0.0001 |
| **Thyroid** | | | | | | | | | |
| 20-49 | (2001, 2019) | 6.04 | (4.81, 7.27) | <0.0001 | - | (2012, 2019) | 3.79 | (1.17, 6.48) | 0.0075 |
| 50+ | (2001, 2019) | 4.91 | (3.52, 6.33) | <0.0001 | 0.12 | (2014, 2019) | 1.98 | (-2.51, 6.67) | 0.37 |

| Age Group | AAPC | | | | | Most Recent APC | | | |
|---|---|---|---|---|---|---|---|---|---|
| | Time Period | AAPC[1] | CI 95% | P Value | P Value Difference | Time Period | APC[2] | CI 95% | P Value |

[1]Average Annual Percentage Change

[2]Annual Percentage Change

**Supplementary Table 5**: Joinpoint average annual percentage change (AAPC) and most recent annual percentage change (APC) for established behavioural risk factor trends by age group for women (**Panel A**) and men (**Panel B**)

**Panel A**

| | AAPC | | | | Most Recent APC | | | |
|---|---|---|---|---|---|---|---|---|
| **Age Group** | **Time Period** | **AAPC[1]** | **CI 95%** | **P Value** | **Time Period** | **APC[2]** | **CI 95%** | **P Value** |
| **Cigarette Smoking - Former** | | | | | | | | |
| 20-49 | (1995, 2019) | -0.04 | (-0.69, 0.61) | 0.9 | (2017, 2019) | -6.12 | (-9.43, -2.69) | 0.0026 |
| 50+ | (1995, 2019) | 0.13 | (-0.07, 0.32) | 0.2 | (2010, 2019) | -0.25 | (-0.68, 0.18) | 0.24 |
| **Cigarette Smoking - Current** | | | | | | | | |
| 20-49 | (1995, 2019) | -2.33 | (-2.60, -2.07) | <0.0001 | (2011, 2019) | -2.23 | (-2.70, -1.76) | <0.0001 |
| 50+ | (1995, 2019) | -2.23 | (-2.61, -1.86) | <0.0001 | (2016, 2019) | 0.34 | (-0.88, 1.58) | 0.55 |
| **Alcohol Consumption - Light Drinker[3]** | | | | | | | | |
| 20-49 | (2011, 2019) | 0.19 | (-0.25, 0.64) | 0.39 | (2016, 2019) | -1.03 | (-2.35, 0.30) | 0.097 |
| 50+ | (2011, 2019) | 0.44 | (0.24, 0.64) | 0.0013 | (2011, 2019) | 0.44 | (0.24, 0.64) | 0.0013 |
| **Alcohol Consumption - Moderate Drinker[3]** | | | | | | | | |
| 20-49 | (2011, 2019) | -3.02 | (-4.25, -1.77) | <0.0001 | (2017, 2019) | -0.46 | (-6.69, 6.17) | 0.85 |
| 50+ | (2011, 2019) | -0.27 | (-0.65, 0.11) | 0.14 | (2011, 2019) | -0.27 | (-0.65, 0.11) | 0.14 |
| **Alcohol Consumption - Heavy Drinker[3]** | | | | | | | | |
| 20-49 | (2011, 2019) | -2.85 | (-5.00, -0.65) | 0.011 | (2014, 2019) | 2.97 | (-0.10, 6.14) | 0.055 |
| 50+ | (2011, 2019) | 0.81 | (-0.61, 2.26) | 0.22 | (2011, 2019) | 0.81 | (-0.61, 2.26) | 0.22 |
| **Body Mass Index (BMI) - Overweight[4]** | | | | | | | | |
| 20-49 | (1995, 2019) | 0.01 | (-0.23, 0.26) | 0.93 | (2016, 2019) | 0.82 | (0.03, 1.61) | 0.043 |
| 50+ | (1995, 2019) | -0.72 | (-0.87, -0.56) | <0.0001 | (2012, 2019) | -1.29 | (-1.75, -0.82) | <0.0001 |
| **Body Mass Index (BMI) - Obese[4]** | | | | | | | | |
| 20-49 | (1995, 2019) | 2.61 | (2.33, 2.88) | <0.0001 | (2013, 2019) | 3.23 | (2.48, 3.99) | <0.0001 |

| Age Group | AAPC Time Period | AAPC[1] | CI 95% | P Value | Most Recent APC Time Period | APC[2] | CI 95% | P Value |
|---|---|---|---|---|---|---|---|---|
| 50+ | (1995, 2019) | 1.61 | (1.00, 2.22) | <0.0001 | (2013, 2019) | 1.90 | (1.15, 2.66) | <0.0001 |
| **Physical Inactivity - Below UK Recommendations[5]** | | | | | | | | |
| 20-49 | (2003, 2012) | -1.44 | (-2.43, -0.44) | 0.02 | (2003, 2012) | -1.44 | (-2.43, -0.44) | 0.02 |
| 50+ | (2003, 2012) | -1.25 | (-1.88, -0.61) | 0.0084 | (2003, 2012) | -1.25 | (-1.88, -0.61) | 0.0084 |
| **Fibre Intake Deficiency - Below UK Recommendations[6]** | | | | | | | | |
| 20-49 | (2008, 2018) | -0.20 | (-0.45, 0.06) | 0.13 | (2012, 2018) | -0.55 | (-0.88, -0.22) | 0.0066 |
| 50+ | (2008, 2018) | -0.43 | (-0.68, -0.18) | 0.00071 | (2014, 2018) | -0.02 | (-0.63, 0.59) | 0.94 |
| **Red Meat Consumption - Median** | | | | | | | | |
| 20-49 | (2008, 2018) | -7.06 | (-12.04, -1.81) | 0.009 | (2015, 2018) | -17.80 | (-32.31, -0.19) | 0.048 |
| 50+ | (2008, 2018) | -7.39 | (-8.64, -6.13) | <0.0001 | (2008, 2018) | -7.39 | (-8.64, -6.13) | <0.0001 |
| **Processed Meat Consumption - Median** | | | | | | | | |
| 20-49 | (2008, 2018) | -5.36 | (-7.88, -2.77) | <0.0001 | (2015, 2018) | -10.99 | (-19.09, -2.09) | 0.024 |
| 50+ | (2008, 2018) | -5.55 | (-7.37, -3.70) | <0.0001 | (2008, 2018) | -5.55 | (-7.37, -3.70) | <0.0001 |

[1]Average Annual Percentage Change

[2]Annual Percentage Change

[3]Heavy (>50g/day), moderate (12-50 g/day), light (<12 g/day)

[4]Overweight (>30 kg/m²), obese (25-30 kg/m²)

[5]Recommendations are 150 min/week of moderate or 75 min/week or vigorous activity

[6]Guidelines are 30g of fibre per day

**Panel B**

| | AAPC | | | | Most Recent APC | | | |
|---|---|---|---|---|---|---|---|---|
| Age Group | Time Period | AAPC[1] | CI 95% | P Value | Time Period | APC[2] | CI 95% | P Value |
| **Cigarette Smoking - Former** | | | | | | | | |
| 20-49 | (1995, 2019) | -0.01 | (-0.33, 0.32) | 0.96 | (2017, 2019) | -3.54 | (-5.93, -1.09) | 0.009 |
| 50+ | (1995, 2019) | -1.07 | (-1.31, -0.82) | <0.0001 | (2015, 2019) | -0.11 | (-1.07, 0.85) | 0.81 |
| **Cigarette Smoking - Current** | | | | | | | | |
| 20-49 | (1995, 2019) | -1.63 | (-2.04, -1.22) | <0.0001 | (2016, 2019) | -1.26 | (-2.43, -0.09) | 0.037 |
| 50+ | (1995, 2019) | -2.25 | (-2.62, -1.88) | <0.0001 | (2013, 2019) | -3.71 | (-4.68, -2.74) | <0.0001 |
| **Alcohol Consumption - Light Drinker[3]** | | | | | | | | |
| 20-49 | (2011, 2019) | 1.30 | (0.70, 1.91) | <0.0001 | (2016, 2019) | 0.35 | (-1.45, 2.18) | 0.62 |
| 50+ | (2011, 2019) | 0.61 | (0.06, 1.16) | 0.029 | (2017, 2019) | -1.31 | (-3.99, 1.44) | 0.25 |
| **Alcohol Consumption - Moderate Drinker[3]** | | | | | | | | |
| 20-49 | (2011, 2019) | -2.17 | (-2.90, -1.44) | <0.0001 | (2017, 2019) | -0.75 | (-4.44, 3.08) | 0.61 |
| 50+ | (2011, 2019) | -1.10 | (-1.56, -0.64) | <0.0001 | (2017, 2019) | 0.69 | (-1.66, 3.09) | 0.47 |
| **Alcohol Consumption - Heavy Drinker[3]** | | | | | | | | |
| 20-49 | (2011, 2019) | -3.56 | (-5.03, -2.06) | <0.0001 | (2016, 2019) | -1.10 | (-5.61, 3.63) | 0.55 |
| 50+ | (2011, 2019) | 0.25 | (-0.27, 0.77) | 0.29 | (2011, 2019) | 0.25 | (-0.27, 0.77) | 0.29 |
| **Body Mass Index (BMI) - Overweight[4]** | | | | | | | | |
| 20-49 | (1995, 2019) | -0.41 | (-0.67, -0.14) | 0.0026 | (2015, 2019) | 0.39 | (-0.49, 1.28) | 0.37 |
| 50+ | (1995, 2019) | -0.39 | (-0.57, -0.20) | <0.0001 | (2007, 2019) | -0.24 | (-0.40, -0.09) | 0.0046 |
| **Body Mass Index (BMI) - Obese[4]** | | | | | | | | |
| 20-49 | (1995, 2019) | 2.19 | (1.89, 2.50) | <0.0001 | (2001, 2019) | 0.79 | (0.58, 1.01) | <0.0001 |
| 50+ | (1995, 2019) | 2.11 | (1.69, 2.53) | <0.0001 | (2009, 2019) | 0.50 | (0.21, 0.80) | 0.0024 |
| **Physical Inactivity - Below UK Recommendations[5]** | | | | | | | | |
| 20-49 | (2003, 2012) | -1.86 | (-3.51, -0.18) | 0.039 | (2003, 2012) | -1.86 | (-3.51, -0.18) | 0.039 |
| 50+ | (2003, 2012) | -0.73 | (-0.93, -0.53) | 0.0014 | (2003, 2012) | -0.73 | (-0.93, -0.53) | 0.0014 |

| | AAPC | | | | Most Recent APC | | | |
|---|---|---|---|---|---|---|---|---|
| **Age Group** | **Time Period** | **AAPC[1]** | **CI 95%** | **P Value** | **Time Period** | **APC[2]** | **CI 95%** | **P Value** |
| **Fibre Intake Deficiency - Below UK Recommendations[6]** | | | | | | | | |
| 20-49 | (2008, 2018) | -0.38 | (-1.34, 0.58) | 0.43 | (2016, 2018) | 2.19 | (-3.31, 8.00) | 0.37 |
| 50+ | (2008, 2018) | -0.30 | (-0.50, -0.10) | 0.0072 | (2008, 2018) | -0.30 | (-0.50, -0.10) | 0.0072 |
| **Red Meat Consumption - Median** | | | | | | | | |
| 20-49 | (2008, 2018) | -7.41 | (-8.92, -5.87) | <0.0001 | (2016, 2018) | -19.99 | (-27.17, -12.10) | 0.0012 |
| 50+ | (2008, 2018) | -4.40 | (-8.10, -0.54) | 0.026 | (2016, 2018) | -13.92 | (-31.34, 7.91) | 0.16 |
| **Processed Meat Consumption - Median** | | | | | | | | |
| 20-49 | (2008, 2018) | -4.88 | (-7.38, -2.32) | 0.00023 | (2012, 2018) | -7.77 | (-10.91, -4.52) | 0.0012 |
| 50+ | (2008, 2018) | -2.03 | (-5.33, 1.38) | 0.24 | (2013, 2018) | -9.97 | (-15.25, -4.36) | 0.0054 |

[1]Average Annual Percentage Change

[2]Annual Percentage Change

[3]Heavy (>50g/day), moderate (12-50 g/day), light (<12 g/day)

[4]Overweight (>30 kg/m²), obese (25-30 kg/m²)

[5]Recommendations are 150 min/week of moderate or 75 min/week or vigorous activity

[6]Guidelines are 30g of fibre per day

**Supplementary Table 6**: Joinpoint average annual percentage change (AAPC) and most recent annual percentage change (APC) for Body Mass Index (BMI) attributable and non-attributable cancer incidence trends for the 11 selected cancer sites for young women (**Panel A**), young men (**Panel B**), older women (**Panel C**), and older men (**Panel D**)

**Panel A**

| | AAPC | | | | Most Recent APC | | | |
|---|---|---|---|---|---|---|---|---|
| **Rate** | **Time Period** | **AAPC[1]** | **CI 95%** | **P Value** | **Time Period** | **APC[2]** | **CI 95%** | **P Value** |
| **Colorectum** | | | | | | | | |
| Attributable | (2005, 2019) | 4.28 | (1.79, 6.83) | 0.00067 | (2012, 2019) | 2.86 | (1.22, 4.53) | 0.0043 |
| Non-Attributable | (2005, 2019) | 3.18 | (2.61, 3.76) | <0.0001 | (2005, 2019) | 3.18 | (2.61, 3.76) | <0.0001 |
| Overall | (2005, 2019) | 3.39 | (2.80, 3.98) | <0.0001 | (2005, 2019) | 3.39 | (2.80, 3.98) | <0.0001 |
| **Endometrium** | | | | | | | | |
| Attributable | (2005, 2019) | 3.80 | (3.09, 4.50) | <0.0001 | (2005, 2019) | 3.80 | (3.09, 4.50) | <0.0001 |
| Non-Attributable | (2005, 2019) | 1.71 | (1.03, 2.40) | 0.00012 | (2005, 2019) | 1.71 | (1.03, 2.40) | 0.00012 |
| Overall | (2005, 2019) | 2.29 | (1.62, 2.96) | <0.0001 | (2005, 2019) | 2.29 | (1.62, 2.96) | <0.0001 |
| **Gallbladder** | | | | | | | | |
| Attributable | (2005, 2019) | 4.31 | (2.28, 6.39) | 0.00048 | (2005, 2019) | 4.31 | (2.28, 6.39) | 0.00048 |
| Non-Attributable | (2005, 2019) | 2.37 | (0.47, 4.31) | 0.018 | (2005, 2019) | 2.37 | (0.47, 4.31) | 0.018 |
| Overall | (2005, 2019) | 2.70 | (0.78, 4.66) | 0.0092 | (2005, 2019) | 2.70 | (0.78, 4.66) | 0.0092 |
| **Kidney** | | | | | | | | |
| Attributable | (2005, 2019) | 5.64 | (3.04, 8.30) | <0.0001 | (2015, 2019) | 1.18 | (-7.01, 10.08) | 0.76 |
| Non-Attributable | (2005, 2019) | 4.31 | (3.24, 5.40) | <0.0001 | (2005, 2019) | 4.31 | (3.24, 5.40) | <0.0001 |
| Overall | (2005, 2019) | 4.71 | (3.60, 5.83) | <0.0001 | (2005, 2019) | 4.71 | (3.60, 5.83) | <0.0001 |
| **Liver** | | | | | | | | |
| Attributable | (2005, 2019) | 4.46 | (3.00, 5.94) | <0.0001 | (2005, 2019) | 4.46 | (3.00, 5.94) | <0.0001 |
| Non-Attributable | (2005, 2019) | 2.35 | (0.94, 3.77) | 0.0031 | (2005, 2019) | 2.35 | (0.94, 3.77) | 0.0031 |

| | AAPC | | | | Most Recent APC | | | |
| --- | --- | --- | --- | --- | --- | --- | --- | --- |
| **Rate** | **Time Period** | **AAPC**[1] | **CI 95%** | **P Value** | **Time Period** | **APC**[2] | **CI 95%** | **P Value** |
| Overall | (2005, 2019) | 2.71 | (1.30, 4.14) | 0.0011 | (2005, 2019) | 2.71 | (1.30, 4.14) | 0.0011 |
| **Multiple Myeloma** | | | | | | | | |
| Attributable | (2005, 2019) | 7.89 | (5.41, 10.42) | <0.0001 | (2013, 2019) | 3.94 | (-0.81, 8.91) | 0.095 |
| Non-Attributable | (2005, 2019) | 6.52 | (5.04, 8.03) | <0.0001 | (2005, 2019) | 6.52 | (5.04, 8.03) | <0.0001 |
| Overall | (2005, 2019) | 6.63 | (5.14, 8.15) | <0.0001 | (2005, 2019) | 6.63 | (5.14, 8.15) | <0.0001 |
| **Ovary** | | | | | | | | |
| Attributable | (2005, 2019) | 2.53 | (1.68, 3.38) | <0.0001 | (2012, 2019) | 0.21 | (-1.12, 1.56) | 0.73 |
| Non-Attributable | (2005, 2019) | 1.10 | (-0.05, 2.26) | 0.06 | (2007, 2019) | 0.14 | (-0.36, 0.65) | 0.55 |
| Overall | (2005, 2019) | 1.19 | (0.01, 2.39) | 0.048 | (2007, 2019) | 0.22 | (-0.30, 0.74) | 0.37 |
| **Pancreas** | | | | | | | | |
| Attributable | (2005, 2019) | 3.94 | (3.04, 4.85) | <0.0001 | (2005, 2019) | 3.94 | (3.04, 4.85) | <0.0001 |
| Non-Attributable | (2005, 2019) | 2.39 | (1.55, 3.25) | <0.0001 | (2005, 2019) | 2.39 | (1.55, 3.25) | <0.0001 |
| Overall | (2005, 2019) | 2.53 | (1.68, 3.38) | <0.0001 | (2005, 2019) | 2.53 | (1.68, 3.38) | <0.0001 |
| **Thyroid** | | | | | | | | |
| Attributable | (2005, 2019) | 7.27 | (6.32, 8.23) | <0.0001 | (2013, 2019) | 3.89 | (2.05, 5.77) | 0.00078 |
| Non-Attributable | (2005, 2019) | 5.49 | (4.58, 6.40) | <0.0001 | (2013, 2019) | 3.38 | (1.60, 5.20) | 0.0017 |
| Overall | (2005, 2019) | 5.62 | (4.71, 6.54) | <0.0001 | (2013, 2019) | 3.43 | (1.64, 5.24) | 0.0015 |

[1]Average Annual Percentage Change

[2]Annual Percentage Change

**Panel B**

| Rate | AAPC | | | | Most Recent APC | | | |
|---|---|---|---|---|---|---|---|---|
| | Time Period | AAPC[1] | CI 95% | P Value | Time Period | APC[2] | CI 95% | P Value |
| **Colorectum** | | | | | | | | |
| Attributable | (2005, 2019) | 3.87 | (3.29, 4.44) | <0.0001 | (2005, 2019) | 3.87 | (3.29, 4.44) | <0.0001 |
| Non-Attributable | (2005, 2019) | 2.48 | (1.94, 3.02) | <0.0001 | (2005, 2019) | 2.48 | (1.94, 3.02) | <0.0001 |
| Overall | (2005, 2019) | 2.69 | (2.16, 3.21) | <0.0001 | (2005, 2019) | 2.69 | (2.16, 3.21) | <0.0001 |
| **Gallbladder** | | | | | | | | |
| Attributable | (2005, 2019) | 5.57 | (3.05, 8.15) | 0.00032 | (2005, 2019) | 5.57 | (3.05, 8.15) | 0.00032 |
| Non-Attributable | (2005, 2019) | 2.51 | (-0.01, 5.09) | 0.051 | (2005, 2019) | 2.51 | (-0.01, 5.09) | 0.051 |
| Overall | (2005, 2019) | 2.79 | (0.28, 5.36) | 0.032 | (2005, 2019) | 2.79 | (0.28, 5.36) | 0.032 |
| **Kidney** | | | | | | | | |
| Attributable | (2005, 2019) | 5.92 | (4.87, 6.98) | <0.0001 | (2015, 2019) | 0.27 | (-3.06, 3.71) | 0.86 |
| Non-Attributable | (2005, 2019) | 4.16 | (2.53, 5.82) | <0.0001 | (2017, 2019) | -3.35 | (-14.12, 8.78) | 0.54 |
| Overall | (2005, 2019) | 4.57 | (3.31, 5.85) | <0.0001 | (2016, 2019) | -0.62 | (-6.20, 5.29) | 0.81 |
| **Liver** | | | | | | | | |
| Attributable | (2005, 2019) | 2.16 | (-1.52, 5.97) | 0.25 | (2014, 2019) | -3.64 | (-12.29, 5.85) | 0.4 |
| Non-Attributable | (2005, 2019) | 0.66 | (-1.04, 2.39) | 0.42 | (2005, 2019) | 0.66 | (-1.04, 2.39) | 0.42 |
| Overall | (2005, 2019) | 1.02 | (-0.72, 2.80) | 0.23 | (2005, 2019) | 1.02 | (-0.72, 2.80) | 0.23 |
| **Multiple Myeloma** | | | | | | | | |
| Attributable | (2005, 2019) | 3.78 | (-0.02, 7.73) | 0.051 | (2013, 2019) | 0.49 | (-3.53, 4.67) | 0.79 |
| Non-Attributable | (2005, 2019) | 4.06 | (2.56, 5.58) | <0.0001 | (2005, 2019) | 4.06 | (2.56, 5.58) | <0.0001 |
| Overall | (2005, 2019) | 4.16 | (2.66, 5.69) | <0.0001 | (2005, 2019) | 4.16 | (2.66, 5.69) | <0.0001 |
| **Pancreas** | | | | | | | | |
| Attributable | (2005, 2019) | 3.72 | (1.94, 5.52) | <0.0001 | (2013, 2019) | 1.23 | (-2.22, 4.80) | 0.45 |
| Non-Attributable | (2005, 2019) | 1.76 | (0.96, 2.56) | 0.00035 | (2005, 2019) | 1.76 | (0.96, 2.56) | 0.00035 |
| Overall | (2005, 2019) | 1.98 | (1.18, 2.79) | 0.00013 | (2005, 2019) | 1.98 | (1.18, 2.79) | 0.00013 |

| Rate | AAPC | | | | Most Recent APC | | | |
|---|---|---|---|---|---|---|---|---|
| | Time Period | AAPC[1] | CI 95% | P Value | Time Period | APC[2] | CI 95% | P Value |
| **Thyroid** | | | | | | | | |
| Attributable | (2005, 2019) | 7.09 | (5.18, 9.04) | <0.0001 | (2011, 2019) | 3.77 | (2.62, 4.94) | 0.00011 |
| Non-Attributable | (2005, 2019) | 5.99 | (4.74, 7.25) | <0.0001 | (2011, 2019) | 3.72 | (2.14, 5.32) | 0.00034 |
| Overall | (2005, 2019) | 5.63 | (3.68, 7.61) | <0.0001 | (2011, 2019) | 3.43 | (2.24, 4.64) | 0.00024 |

[1]Average Annual Percentage Change

[2]Annual Percentage Change

**Panel C**

| | AAPC | | | | Most Recent APC | | | |
|---|---|---|---|---|---|---|---|---|
| Rate | Time Period | AAPC[1] | CI 95% | P Value | Time Period | APC[2] | CI 95% | P Value |
| **Breast** | | | | | | | | |
| Attributable | (2005, 2019) | 1.01 | (0.64, 1.38) | <0.0001 | (2014, 2019) | 0.25 | (-0.68, 1.19) | 0.57 |
| Non-Attributable | (2005, 2019) | 0.33 | (0.10, 0.55) | 0.0075 | (2005, 2019) | 0.33 | (0.10, 0.55) | 0.0075 |
| Overall | (2005, 2019) | 0.39 | (0.17, 0.61) | 0.0023 | (2005, 2019) | 0.39 | (0.17, 0.61) | 0.0023 |
| **Colorectum** | | | | | | | | |
| Attributable | (2005, 2019) | 0.74 | (0.21, 1.27) | 0.0064 | (2009, 2019) | -0.16 | (-0.60, 0.28) | 0.43 |
| Non-Attributable | (2005, 2019) | -0.03 | (-0.63, 0.58) | 0.93 | (2017, 2019) | 1.88 | (-2.02, 5.92) | 0.3 |
| Overall | (2005, 2019) | 0.12 | (-0.51, 0.76) | 0.7 | (2017, 2019) | 1.98 | (-2.10, 6.23) | 0.29 |
| **Endometrium** | | | | | | | | |
| Attributable | (2005, 2019) | 2.18 | (1.74, 2.62) | <0.0001 | (2011, 2019) | 0.83 | (0.27, 1.39) | 0.0082 |
| Non-Attributable | (2005, 2019) | 0.80 | (0.34, 1.26) | 0.00062 | (2013, 2019) | -0.35 | (-1.26, 0.57) | 0.42 |
| Overall | (2005, 2019) | 1.29 | (0.87, 1.72) | <0.0001 | (2012, 2019) | 0.16 | (-0.51, 0.83) | 0.62 |
| **Gallbladder** | | | | | | | | |
| Attributable | (2005, 2019) | 4.03 | (3.11, 4.95) | <0.0001 | (2010, 2019) | 3.03 | (2.08, 3.98) | <0.0001 |
| Non-Attributable | (2005, 2019) | 2.75 | (2.25, 3.26) | <0.0001 | (2005, 2019) | 2.75 | (2.25, 3.26) | <0.0001 |
| Overall | (2005, 2019) | 3.00 | (2.50, 3.51) | <0.0001 | (2005, 2019) | 3.00 | (2.50, 3.51) | <0.0001 |
| **Kidney** | | | | | | | | |
| Attributable | (2005, 2019) | 3.47 | (2.42, 4.54) | <0.0001 | (2013, 2019) | 0.35 | (-1.70, 2.44) | 0.71 |
| Non-Attributable | (2005, 2019) | 2.40 | (1.53, 3.27) | <0.0001 | (2014, 2019) | -0.88 | (-3.00, 1.30) | 0.39 |
| Overall | (2005, 2019) | 2.66 | (1.75, 3.58) | <0.0001 | (2014, 2019) | -0.76 | (-2.99, 1.53) | 0.48 |
| **Liver** | | | | | | | | |
| Attributable | (2005, 2019) | 4.63 | (3.26, 6.01) | <0.0001 | (2013, 2019) | 1.34 | (-1.30, 4.05) | 0.29 |
| Non-Attributable | (2005, 2019) | 3.38 | (2.15, 4.63) | <0.0001 | (2013, 2019) | 0.64 | (-1.76, 3.10) | 0.57 |
| Overall | (2005, 2019) | 3.66 | (2.41, 4.93) | <0.0001 | (2013, 2019) | 0.81 | (-1.63, 3.31) | 0.48 |

| Rate | AAPC | | | | Most Recent APC | | | |
|---|---|---|---|---|---|---|---|---|
| | Time Period | AAPC[1] | CI 95% | P Value | Time Period | APC[2] | CI 95% | P Value |
| **Multiple Myeloma** | | | | | | | | |
| Attributable | (2005, 2019) | 3.42 | (2.62, 4.22) | <0.0001 | (2005, 2019) | 3.42 | (2.62, 4.22) | <0.0001 |
| Non-Attributable | (2005, 2019) | 2.67 | (1.88, 3.47) | <0.0001 | (2005, 2019) | 2.67 | (1.88, 3.47) | <0.0001 |
| Overall | (2005, 2019) | 2.74 | (1.95, 3.54) | <0.0001 | (2005, 2019) | 2.74 | (1.95, 3.54) | <0.0001 |
| **Ovary** | | | | | | | | |
| Attributable | (2005, 2019) | -0.22 | (-0.82, 0.38) | 0.48 | (2011, 2019) | -1.13 | (-1.89, -0.35) | 0.009 |
| Non-Attributable | (2005, 2019) | -1.35 | (-1.66, -1.04) | <0.0001 | (2005, 2019) | -1.35 | (-1.66, -1.04) | <0.0001 |
| Overall | (2005, 2019) | -1.29 | (-1.60, -0.97) | <0.0001 | (2005, 2019) | -1.29 | (-1.60, -0.97) | <0.0001 |
| **Pancreas** | | | | | | | | |
| Attributable | (2005, 2019) | 1.41 | (0.99, 1.83) | <0.0001 | (2013, 2019) | 0.35 | (-0.48, 1.19) | 0.37 |
| Non-Attributable | (2005, 2019) | 0.66 | (0.33, 0.99) | <0.0001 | (2013, 2019) | 0.01 | (-0.65, 0.67) | 0.98 |
| Overall | (2005, 2019) | 0.75 | (0.42, 1.08) | <0.0001 | (2013, 2019) | 0.05 | (-0.61, 0.72) | 0.86 |
| **Thyroid** | | | | | | | | |
| Attributable | (2005, 2019) | 5.42 | (4.39, 6.46) | <0.0001 | (2014, 2019) | 2.53 | (-0.02, 5.15) | 0.052 |
| Non-Attributable | (2005, 2019) | 4.45 | (3.33, 5.58) | <0.0001 | (2014, 2019) | 2.10 | (-0.67, 4.95) | 0.12 |
| Overall | (2005, 2019) | 4.55 | (3.45, 5.66) | <0.0001 | (2014, 2019) | 2.15 | (-0.59, 4.96) | 0.11 |

[1]Average Annual Percentage Change

[2]Annual Percentage Change

**Panel D**

| Rate | AAPC | | | | Most Recent APC | | | |
|---|---|---|---|---|---|---|---|---|
| | Time Period | AAPC[1] | CI 95% | P Value | Time Period | APC[2] | CI 95% | P Value |
| **Colorectum** | | | | | | | | |
| Attributable | (2005, 2019) | 0.78 | (0.38, 1.19) | 0.00016 | (2015, 2019) | 1.12 | (0.27, 1.97) | 0.016 |
| Non-Attributable | (2005, 2019) | -0.78 | (-1.64, 0.08) | 0.074 | (2014, 2019) | -0.86 | (-1.82, 0.11) | 0.074 |
| Overall | (2005, 2019) | -0.51 | (-1.32, 0.31) | 0.22 | (2014, 2019) | -0.58 | (-1.49, 0.34) | 0.18 |
| **Gallbladder** | | | | | | | | |
| Attributable | (2005, 2019) | 5.27 | (4.29, 6.26) | <0.0001 | (2005, 2019) | 5.27 | (4.29, 6.26) | <0.0001 |
| Non-Attributable | (2005, 2019) | 1.90 | (1.08, 2.74) | 0.00024 | (2005, 2019) | 1.90 | (1.08, 2.74) | 0.00024 |
| Overall | (2005, 2019) | 2.31 | (1.47, 3.15) | <0.0001 | (2005, 2019) | 2.31 | (1.47, 3.15) | <0.0001 |
| **Kidney** | | | | | | | | |
| Attributable | (2005, 2019) | 3.91 | (2.94, 4.89) | <0.0001 | (2014, 2019) | 1.00 | (-1.41, 3.46) | 0.38 |
| Non-Attributable | (2005, 2019) | 2.15 | (1.08, 3.22) | <0.0001 | (2014, 2019) | -0.35 | (-2.98, 2.36) | 0.78 |
| Overall | (2005, 2019) | 2.51 | (1.47, 3.57) | <0.0001 | (2014, 2019) | -0.04 | (-2.63, 2.62) | 0.97 |
| **Liver** | | | | | | | | |
| Attributable | (2005, 2019) | 6.04 | (5.52, 6.56) | <0.0001 | (2013, 2019) | 3.32 | (2.30, 4.34) | <0.0001 |
| Non-Attributable | (2005, 2019) | 3.67 | (2.90, 4.44) | <0.0001 | (2014, 2019) | 0.99 | (-0.91, 2.94) | 0.27 |
| Overall | (2005, 2019) | 4.29 | (3.74, 4.85) | <0.0001 | (2013, 2019) | 2.04 | (0.95, 3.14) | 0.0018 |
| **Multiple Myeloma** | | | | | | | | |
| Attributable | (2005, 2019) | 4.02 | (3.31, 4.73) | <0.0001 | (2005, 2019) | 4.02 | (3.31, 4.73) | <0.0001 |
| Non-Attributable | (2005, 2019) | 2.73 | (2.04, 3.42) | <0.0001 | (2005, 2019) | 2.73 | (2.04, 3.42) | <0.0001 |
| Overall | (2005, 2019) | 2.86 | (2.17, 3.55) | <0.0001 | (2005, 2019) | 2.86 | (2.17, 3.55) | <0.0001 |
| **Pancreas** | | | | | | | | |
| Attributable | (2005, 2019) | 3.02 | (2.51, 3.53) | <0.0001 | (2013, 2019) | 2.18 | (1.18, 3.20) | 0.00065 |
| Non-Attributable | (2005, 2019) | 0.78 | (0.51, 1.04) | <0.0001 | (2005, 2019) | 0.78 | (0.51, 1.04) | <0.0001 |
| Overall | (2005, 2019) | 1.08 | (0.82, 1.34) | <0.0001 | (2005, 2019) | 1.08 | (0.82, 1.34) | <0.0001 |

| Rate | AAPC | | | | Most Recent APC | | | |
|---|---|---|---|---|---|---|---|---|
| | Time Period | AAPC[1] | CI 95% | P Value | Time Period | APC[2] | CI 95% | P Value |
| **Thyroid** | | | | | | | | |
| Attributable | (2005, 2019) | 6.45 | (4.66, 8.26) | <0.0001 | (2013, 2019) | 3.39 | (-0.07, 6.98) | 0.054 |
| Non-Attributable | (2005, 2019) | 4.55 | (2.84, 6.29) | <0.0001 | (2014, 2019) | 1.37 | (-2.83, 5.76) | 0.49 |
| Overall | (2005, 2019) | 4.73 | (3.01, 6.48) | <0.0001 | (2014, 2019) | 1.53 | (-2.70, 5.93) | 0.45 |

[1]Average Annual Percentage Change

[2]Annual Percentage Change

**Supplementary Table 7**: PubMed search terms to identify relative risks for established risk factors and cancer risk associations that were updated from Brown et al, 2018[6]

| Cancer Site | PubMed Search Term |
|---|---|
| Oral | (oral OR mouth) AND (cancer OR tumour) |
| Pancreas | (pancreas OR pancreatic) AND (cancer OR tumour) |
| Colorectum* | (colorectal OR colorectum) AND (cancer OR tumour) |
| Liver | (liver OR hepatic OR hepatocellular) AND (cancer OR tumour) |
| Breast | breast AND (cancer OR tumour) |
| Endometrium | (endometrium OR endometrial) AND (cancer OR tumour) |
| Kidney | (kidney OR renal OR renal cell) AND (cancer OR carcinoma OR tumour) |
| Ovarian | (ovary OR ovarian) AND (cancer OR carcinoma OR tumour) |
| **Risk Factor** | **PubMed Search Term** |
| Smoking | tobacco OR cigarette OR smoking OR environmental tobacco smoke OR secondhand smoke |
| Alcohol | alcohol OR alcoholic OR ethanol |
| BMI | weight OR BMI OR body mass index OR obesity OR obese OR overweight OR adiposity OR body size |
| Physical Activity | physical OR activity OR exercise OR physically active OR sedentary |
| Red Meat* | Meat OR bacon OR ham OR sausages OR jerky OR salami OR cured OR salted |
| Processed Meat | Meat OR bacon OR ham OR sausages OR jerky OR salami OR cured OR salted |
| Fiber | fibre OR fiber |

*Adjusted search terms from Brown et. al methodology

### Supplementary Figures

**Supplementary Figure 1:** Joinpoint average annual percentage change (AAPC) in cancer incidence rates in England between 2001-2019 by age group for 22 cancers in women (**Panel A**) and 21 cancers in men (**Panel B**). Figure for women doesn't show Kaposi sarcoma as levels were too low to generate an AAPC.

#### Panel A

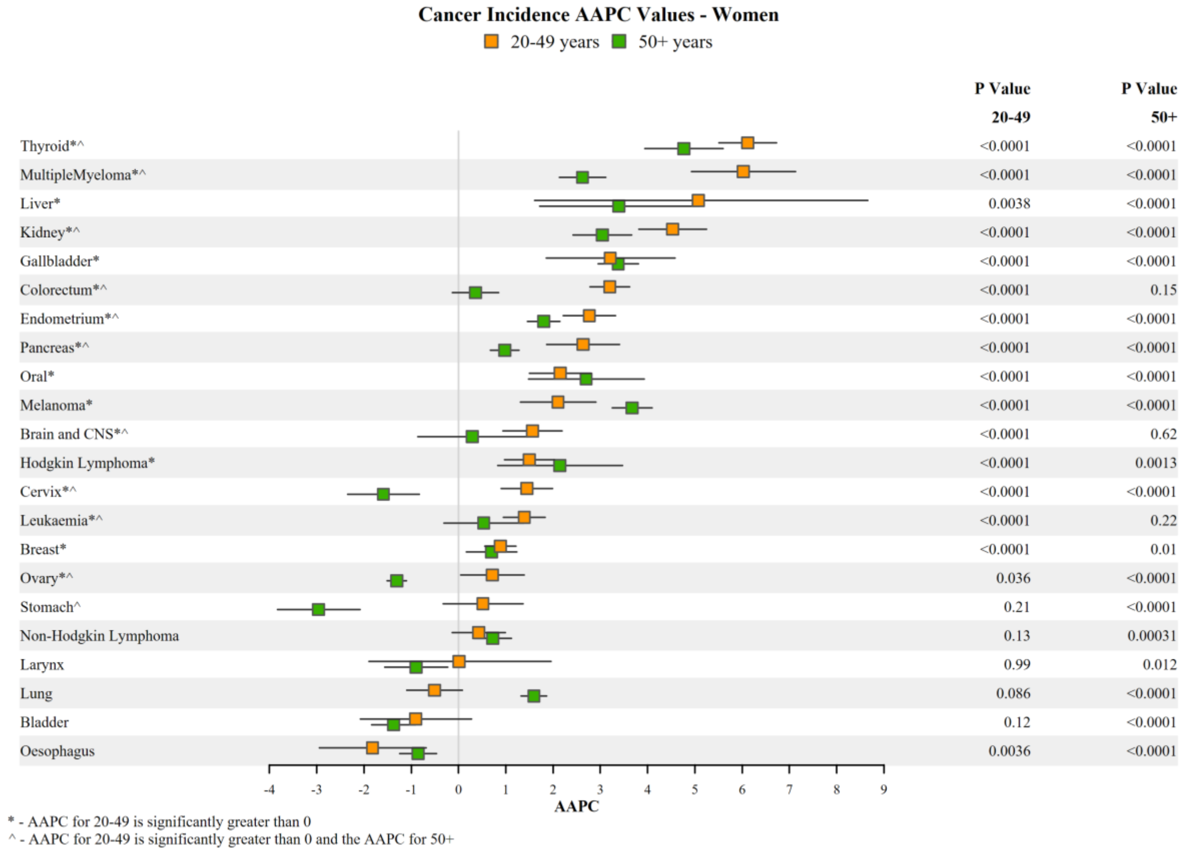

### Panel B

**Cancer Incidence AAPC Values - Men**

20-49 years  50+ years

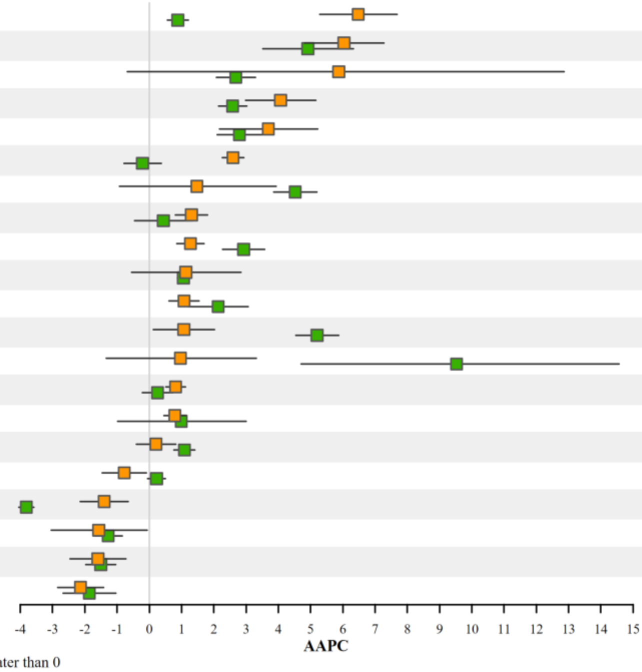

| | P Value 20-49 | P Value 50+ |
|---|---|---|
| Prostate*^ | <0.0001 | <0.0001 |
| Thyroid* | <0.0001 | <0.0001 |
| Gallbladder | 0.08 | <0.0001 |
| MultipleMyeloma*^ | <0.0001 | <0.0001 |
| Kidney* | <0.0001 | <0.0001 |
| Colorectum*^ | <0.0001 | 0.48 |
| Liver | 0.23 | <0.0001 |
| Leukaemia*^ | <0.0001 | 0.34 |
| Oral* | <0.0001 | <0.0001 |
| Pancreas | 0.19 | <0.0001 |
| Hodgkin Lymphoma* | 0.00014 | <0.0001 |
| Melanoma* | 0.026 | <0.0001 |
| Kaposi Sarcoma | 0.41 | 0.00052 |
| Brain and CNS*^ | <0.0001 | 0.29 |
| Testis* | 0.00012 | 0.33 |
| Non-Hodgkin Lymphoma | 0.51 | <0.0001 |
| Oesophagus | 0.028 | 0.11 |
| Stomach^ | 0.001 | <0.0001 |
| Lung | 0.041 | <0.0001 |
| Larynx | 0.0013 | <0.0001 |
| Bladder | <0.0001 | <0.0001 |

* - AAPC for 20-49 is significantly greater than 0
^ - AAPC for 20-49 is significantly greater than 0 and the AAPC for 50+

**Supplementary Figure 2**: Flowchart showing the selection of cancer sites and risk factors, and sources of data for the calculation of population attributable fractions (PAF)

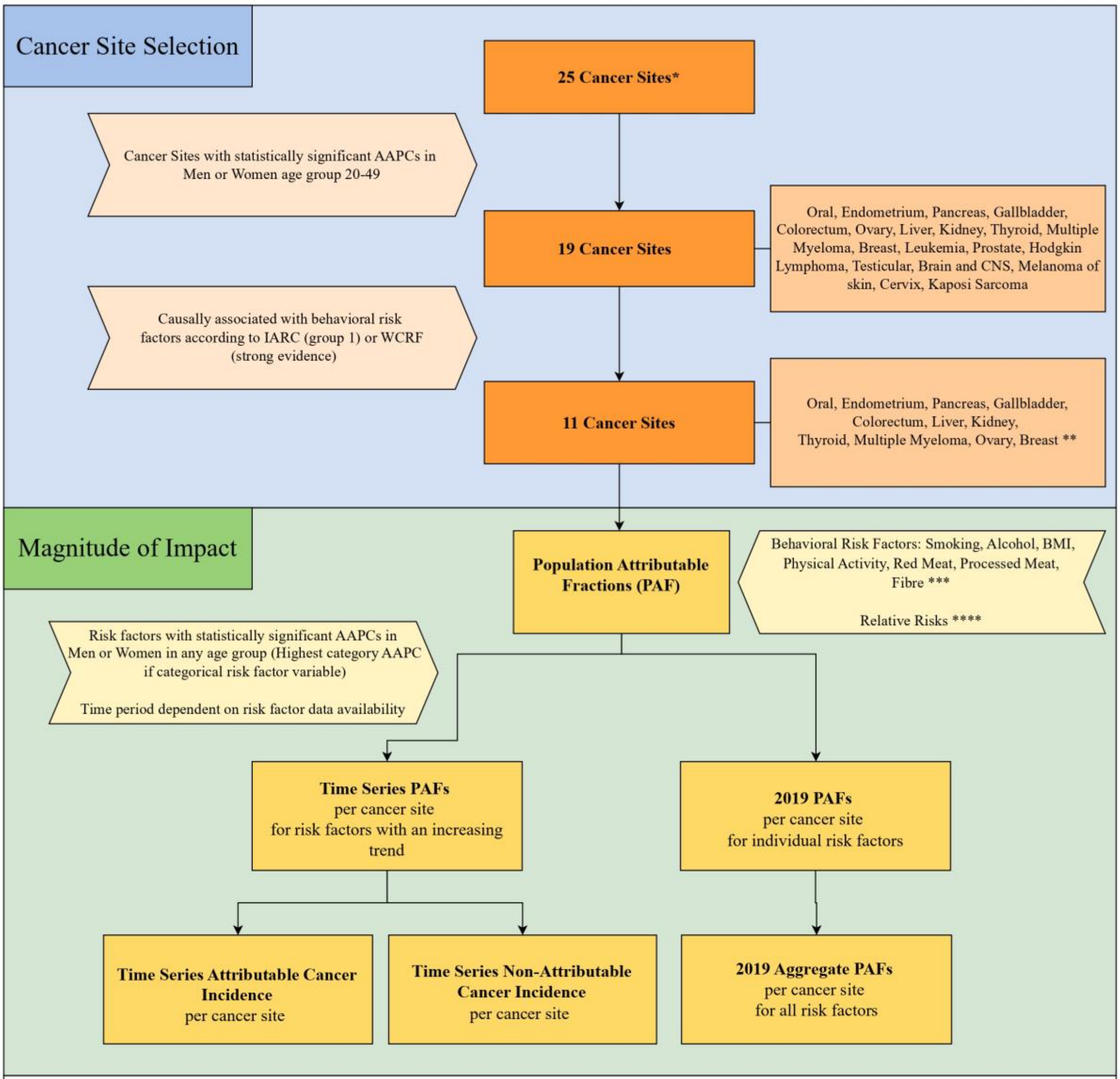

* National Disease Registration Service (NDNS) for England 2001-2019, stratified by ages (20-49, 50+) and sex (Men,Women)

** Leukaemia, prostate, hodgkin lymphoma, brain/CNS, melanoma, testicular cancer excluded because they do not have established behavioural risk factors. Cervical cancer and Kaposi sarcoma were excluded due to the strong association with infections that drive their aetiology.

*** Supplementary Table 1

**** Supplementary Table 3

**Supplementary Figure 3:** Trends in established behavioural risk factors in England (1995-2019) by Index of Multiple Deprivation (IMD) status with estimates of the Joinpoint average annual percentage change (AAPC).

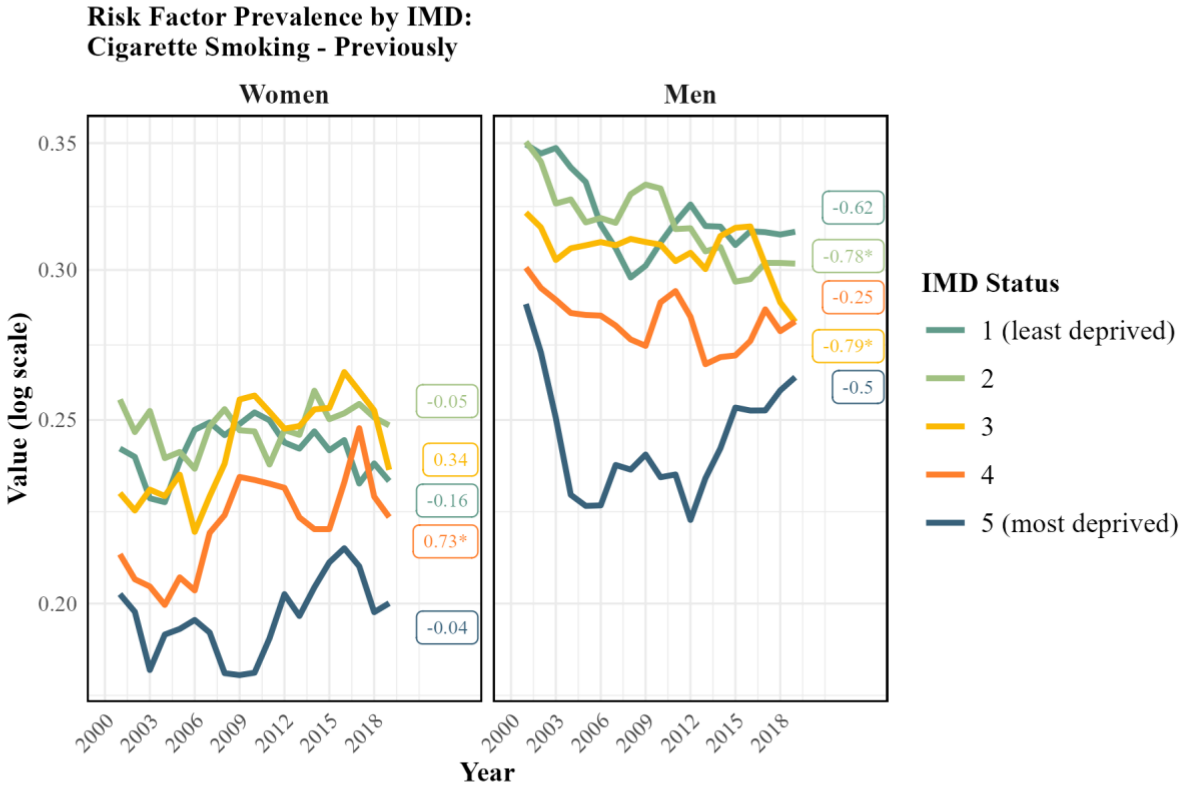

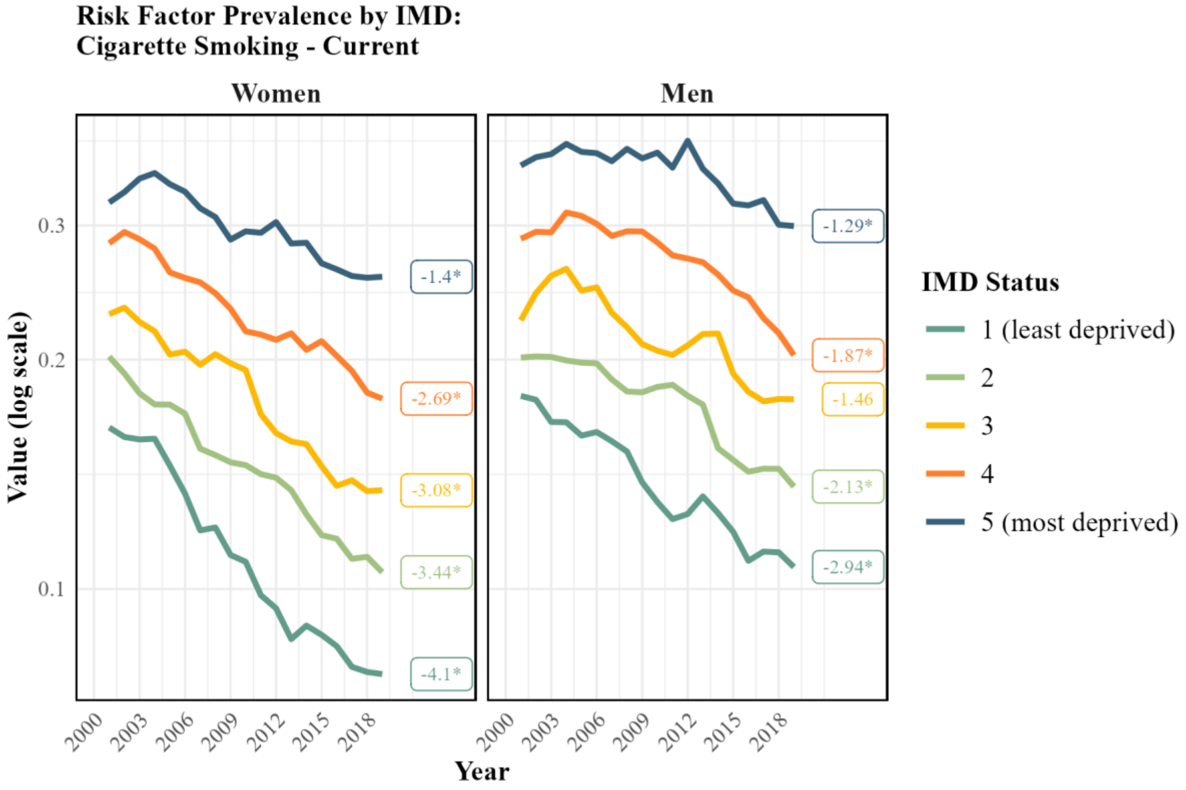

### Risk Factor Prevalence by IMD:
### Alcohol Consumption - Light Drinker

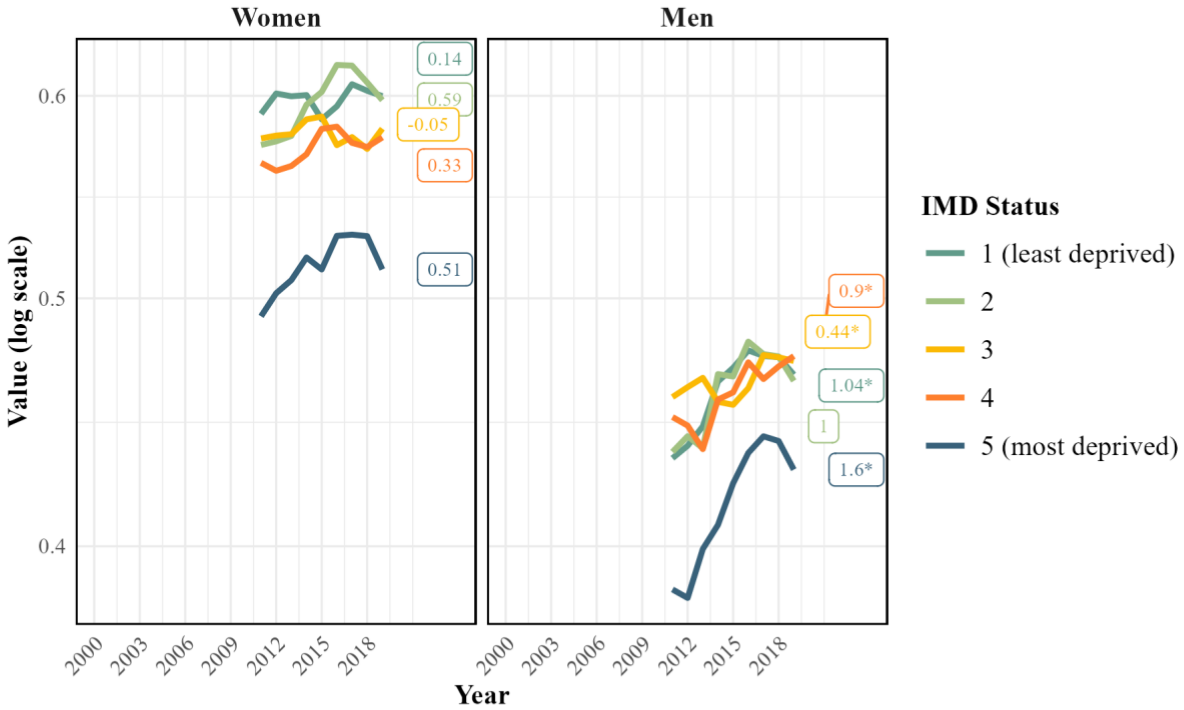

### Risk Factor Prevalence by IMD:
### Alcohol Consumption - Moderate Drinker

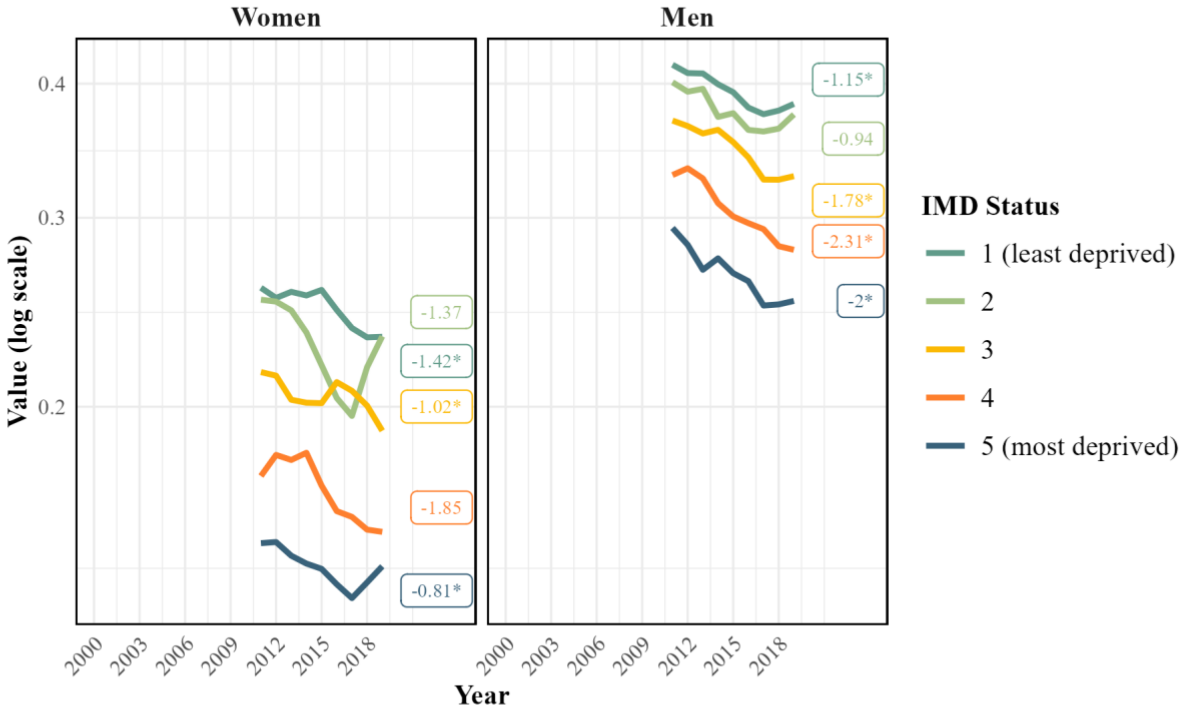

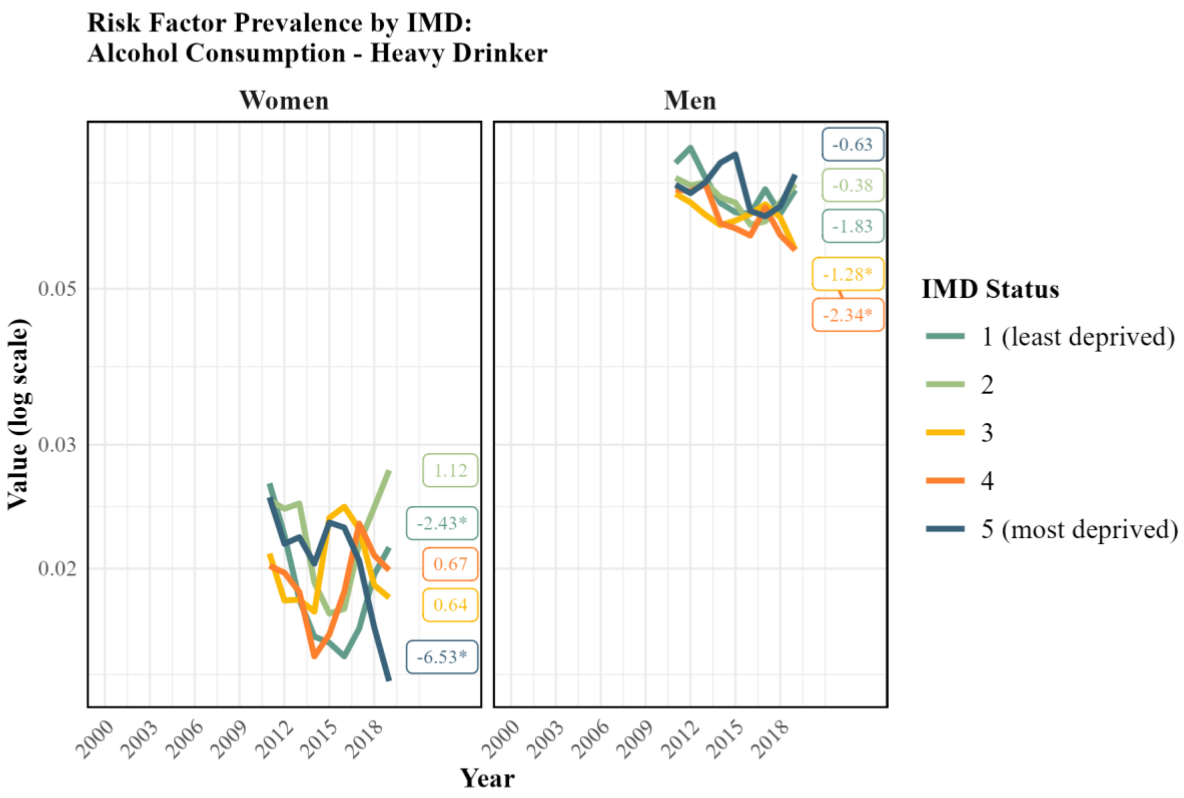
Risk Factor Prevalence by IMD:
Alcohol Consumption - Heavy Drinker
Women
Men
Value (log scale)
0.05
0.03
0.02
1.12
-2.43*
0.67
0.64
-6.53*
-0.63
-0.38
-1.83
-1.28*
-2.34*
2000
2003
2006
2009
2012
2015
2018
Year
IMD Status
1 (least deprived)
2
3
4
5 (most deprived)

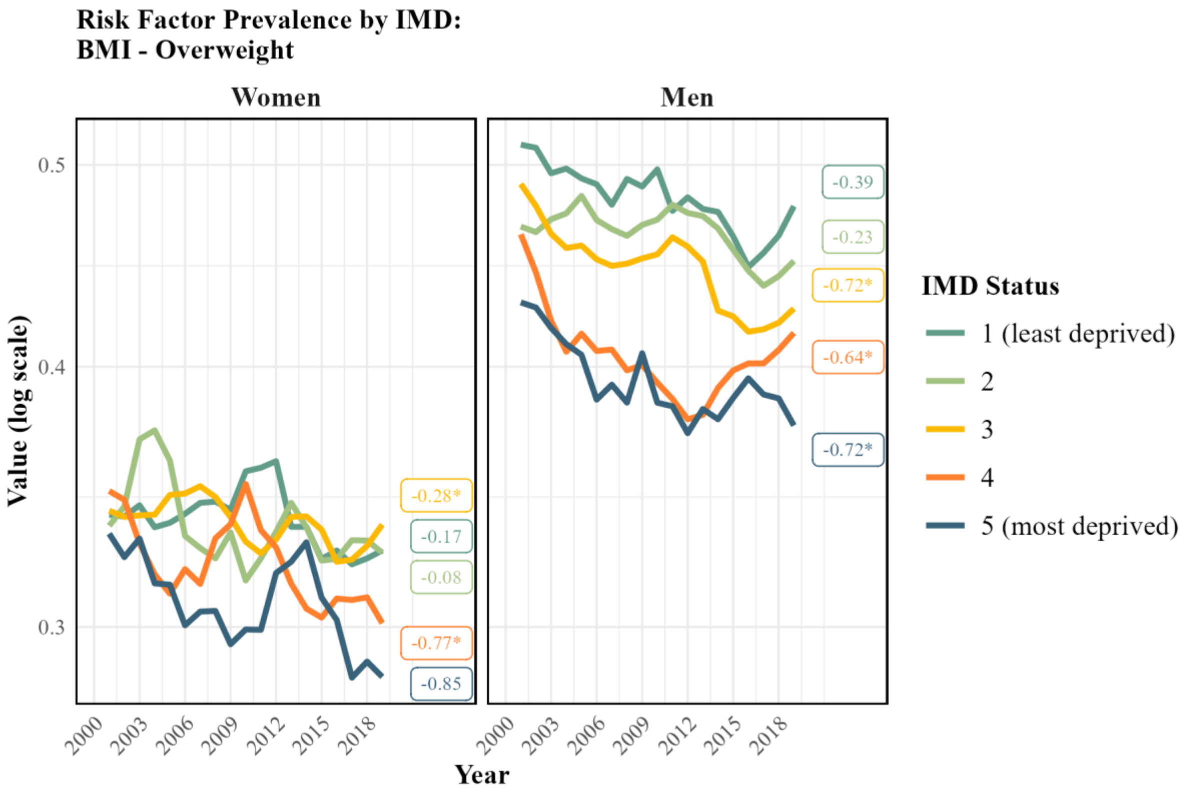
Risk Factor Prevalence by IMD:
BMI - Overweight
Women
Men
Value (log scale)
0.5
0.4
0.3
-0.28*
-0.17
-0.08
-0.77*
-0.85
-0.39
-0.23
-0.72*
-0.64*
-0.72*
2000
2003
2006
2009
2012
2015
2018
Year
IMD Status
1 (least deprived)
2
3
4
5 (most deprived)

#### Risk Factor Prevalence by IMD:
#### BMI - Obese

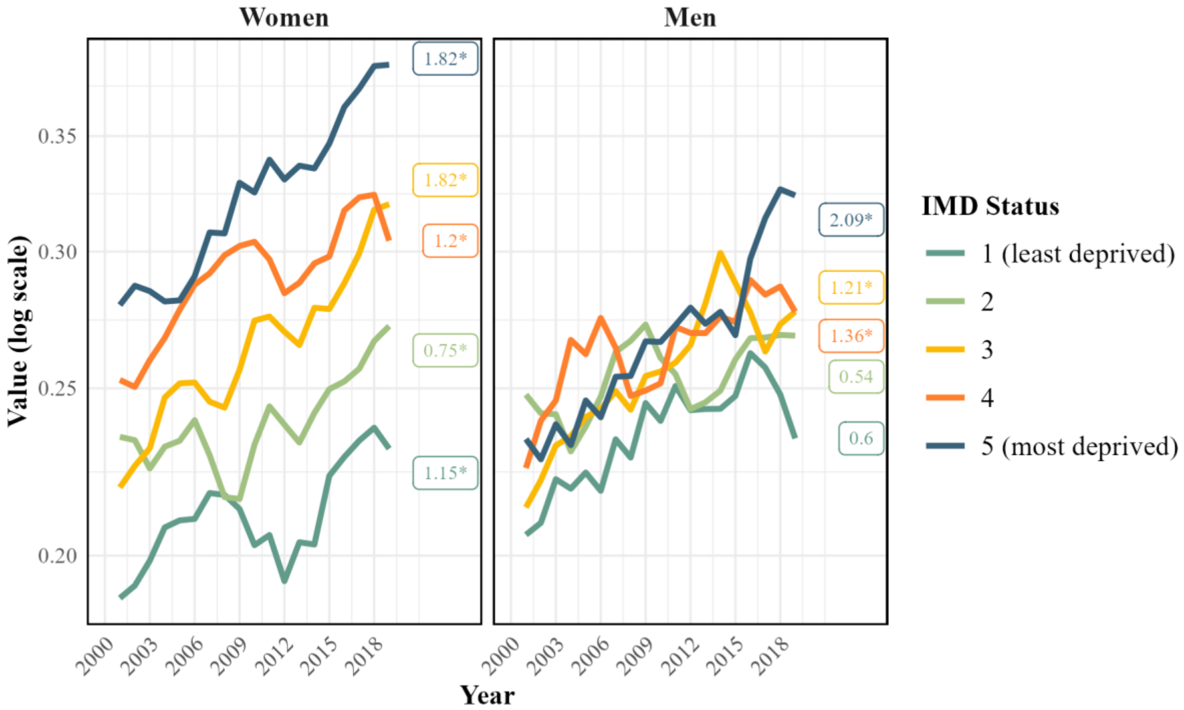

#### Risk Factor Prevalence by IMD:
#### Physical Inactivity - Below Old Recommendations

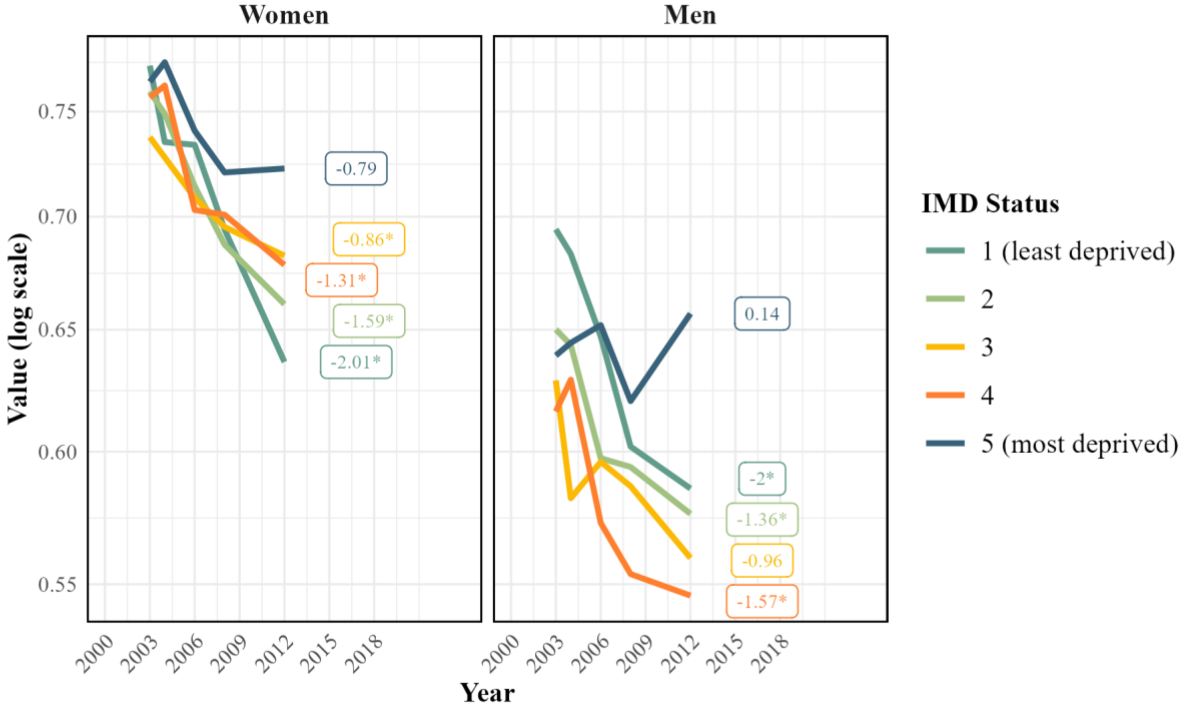

**Supplementary Figure 4:** Risk factor specific population attributable fractions (PAF) for established behavioural risk factors in 11 cancer sites by age group and sex in England, 2019

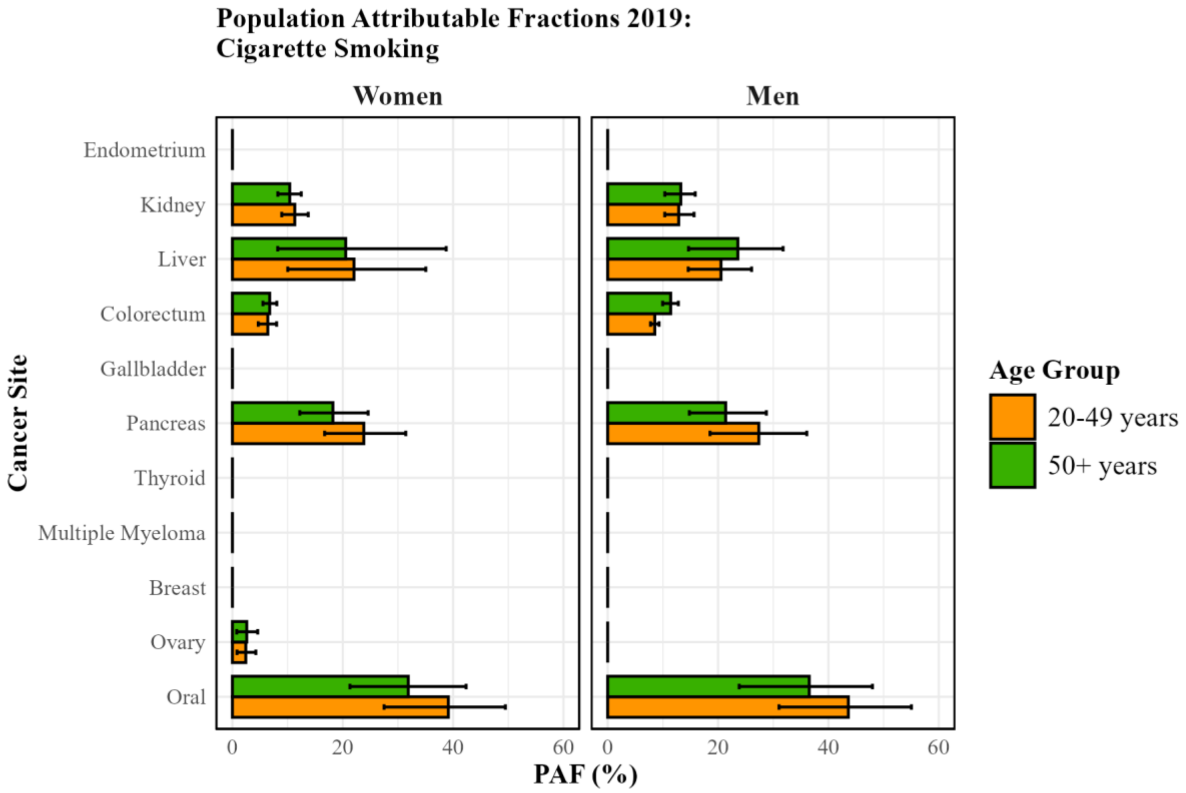

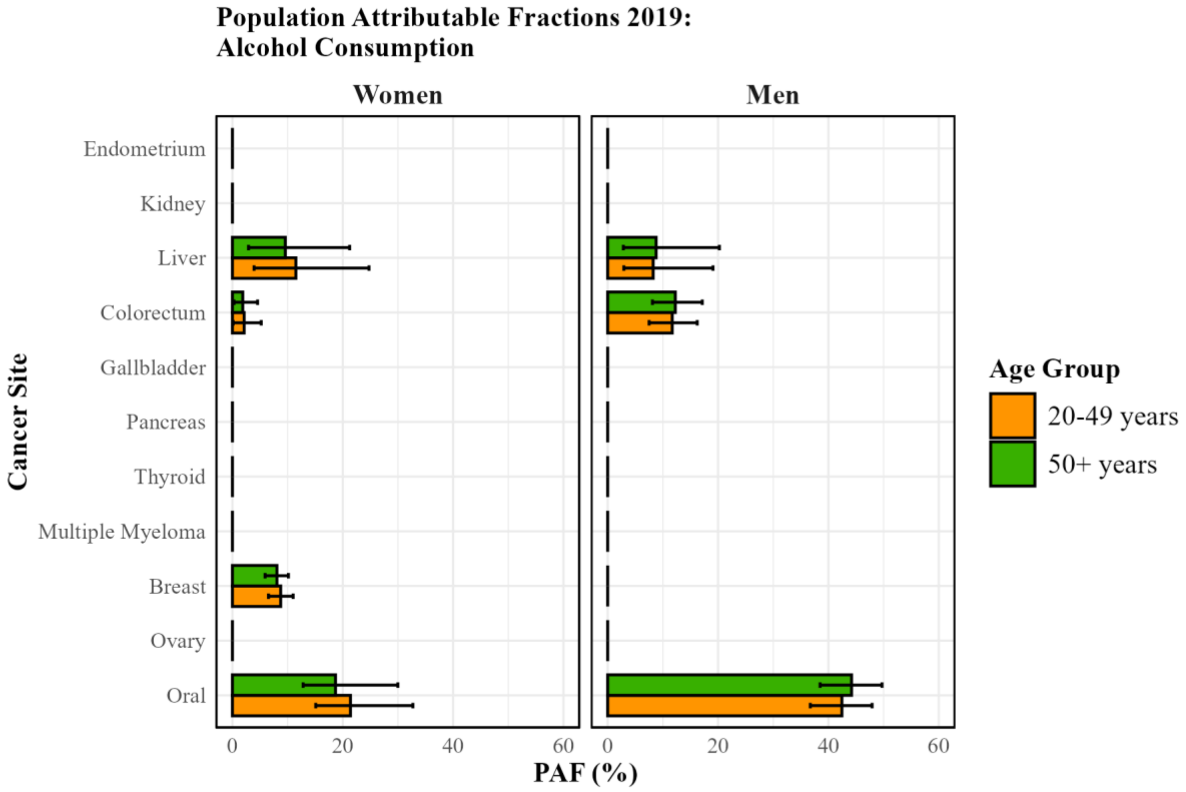

**Population Attributable Fractions 2019:
Body Mass Intake (BMI)**

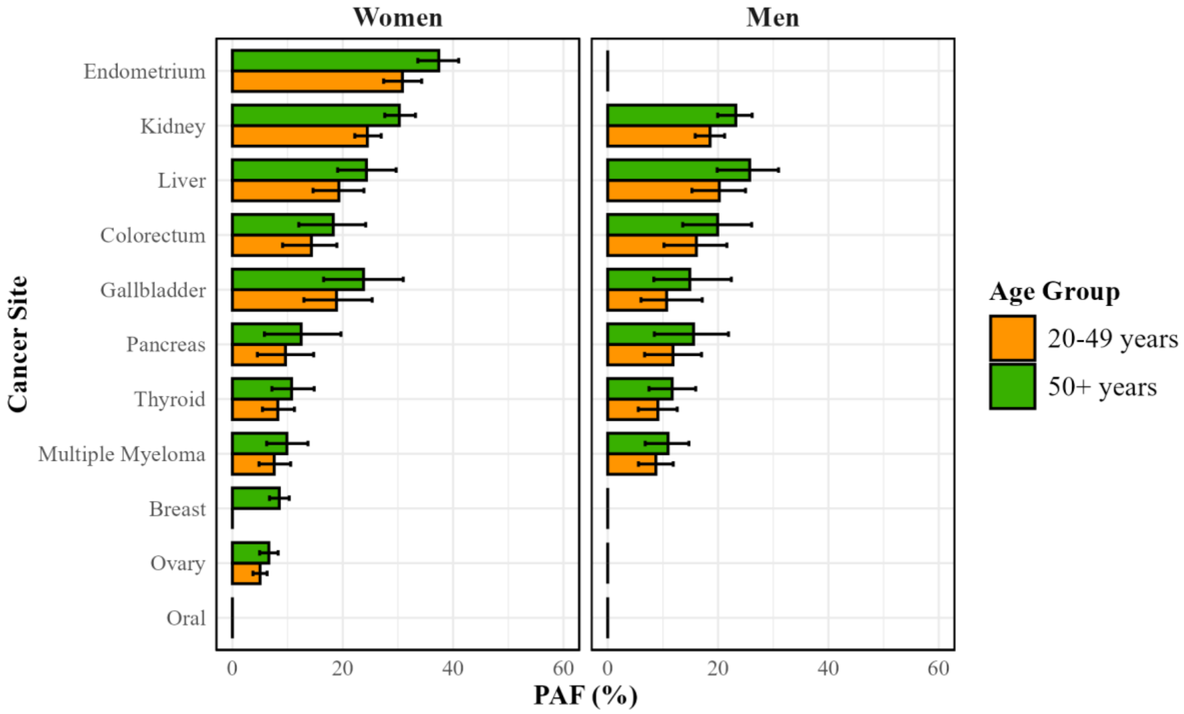

**Population Attributable Fractions 2019:
Physical Inactivity**

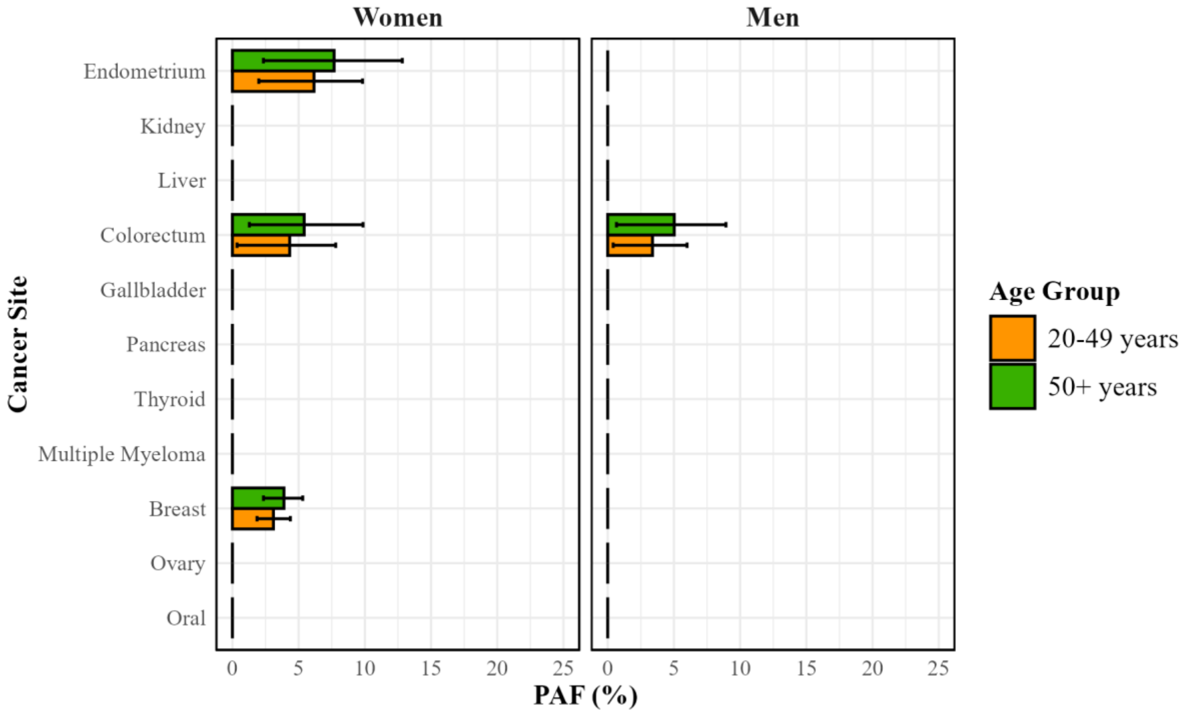

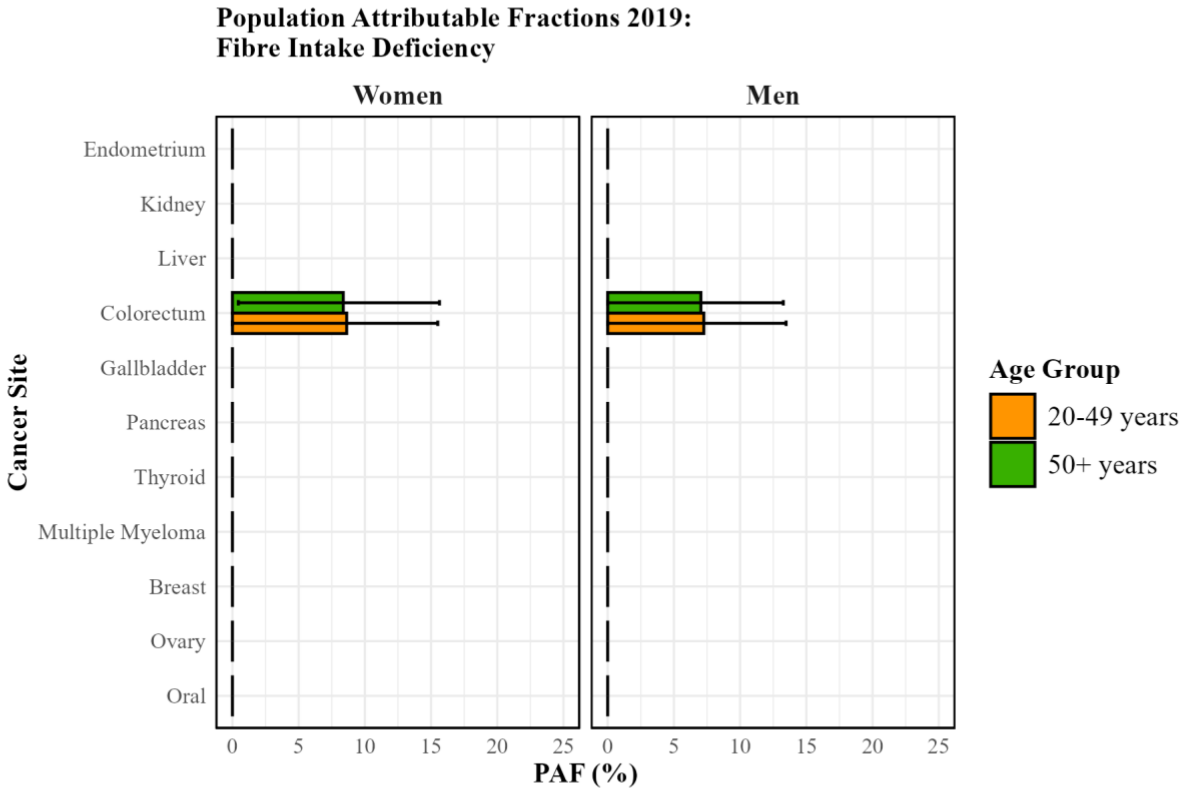
Population Attributable Fractions 2019:
Fibre Intake Deficiency
Women
Men
Cancer Site
Endometrium
Kidney
Liver
Colorectum
Gallbladder
Pancreas
Thyroid
Multiple Myeloma
Breast
Ovary
Oral
0
5
10
15
20
25
PAF (%)
Age Group
20-49 years
50+ years

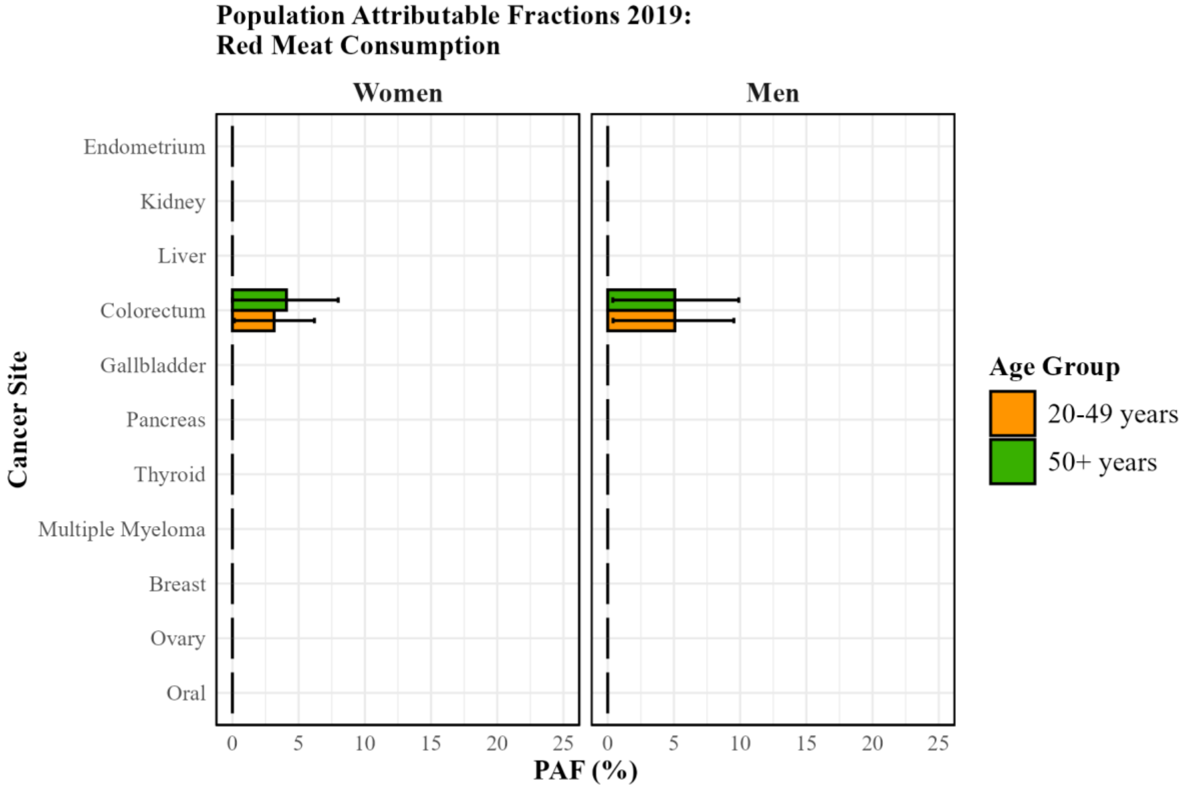
Population Attributable Fractions 2019:
Red Meat Consumption
Women
Men
Cancer Site
Endometrium
Kidney
Liver
Colorectum
Gallbladder
Pancreas
Thyroid
Multiple Myeloma
Breast
Ovary
Oral
0
5
10
15
20
25
PAF (%)
Age Group
20-49 years
50+ years

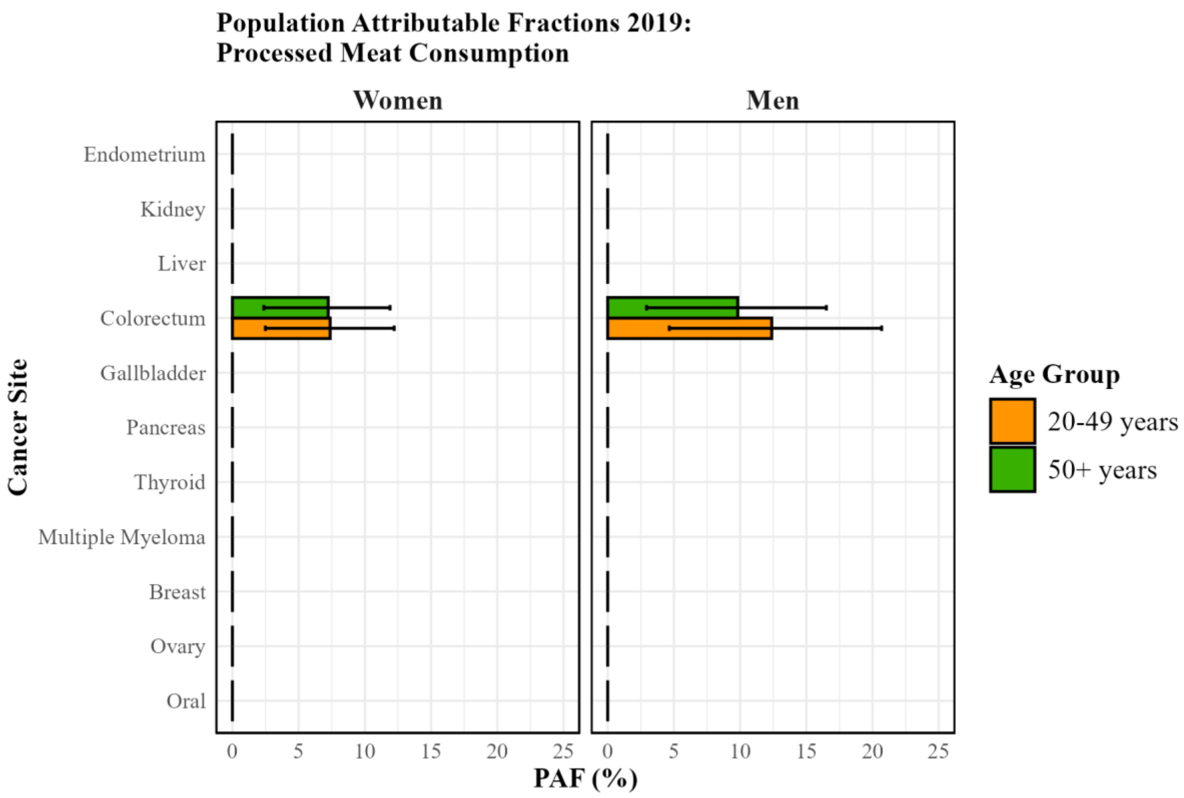
Population Attributable Fractions 2019:
Processed Meat Consumption
Women
Men
Cancer Site
Endometrium
Kidney
Liver
Colorectum
Gallbladder
Pancreas
Thyroid
Multiple Myeloma
Breast
Ovary
Oral
0 5 10 15 20 25
PAF (%)
Age Group
20-49 years
50+ years

**Supplementary Figure 5:** Cancer incidence rates for older adults (50+ years) by sex for the 11 selected cancers partitioned into overall, BMI-attributable, and BMI-non-attributable cancer incidence rates, England 2001-2019. See **Figure 4** for similar figures for younger adults, and **Supplementary Table 6** for estimates of the most recent annual percentage change (APC) and the average APC (AAPC).

**Panel A**

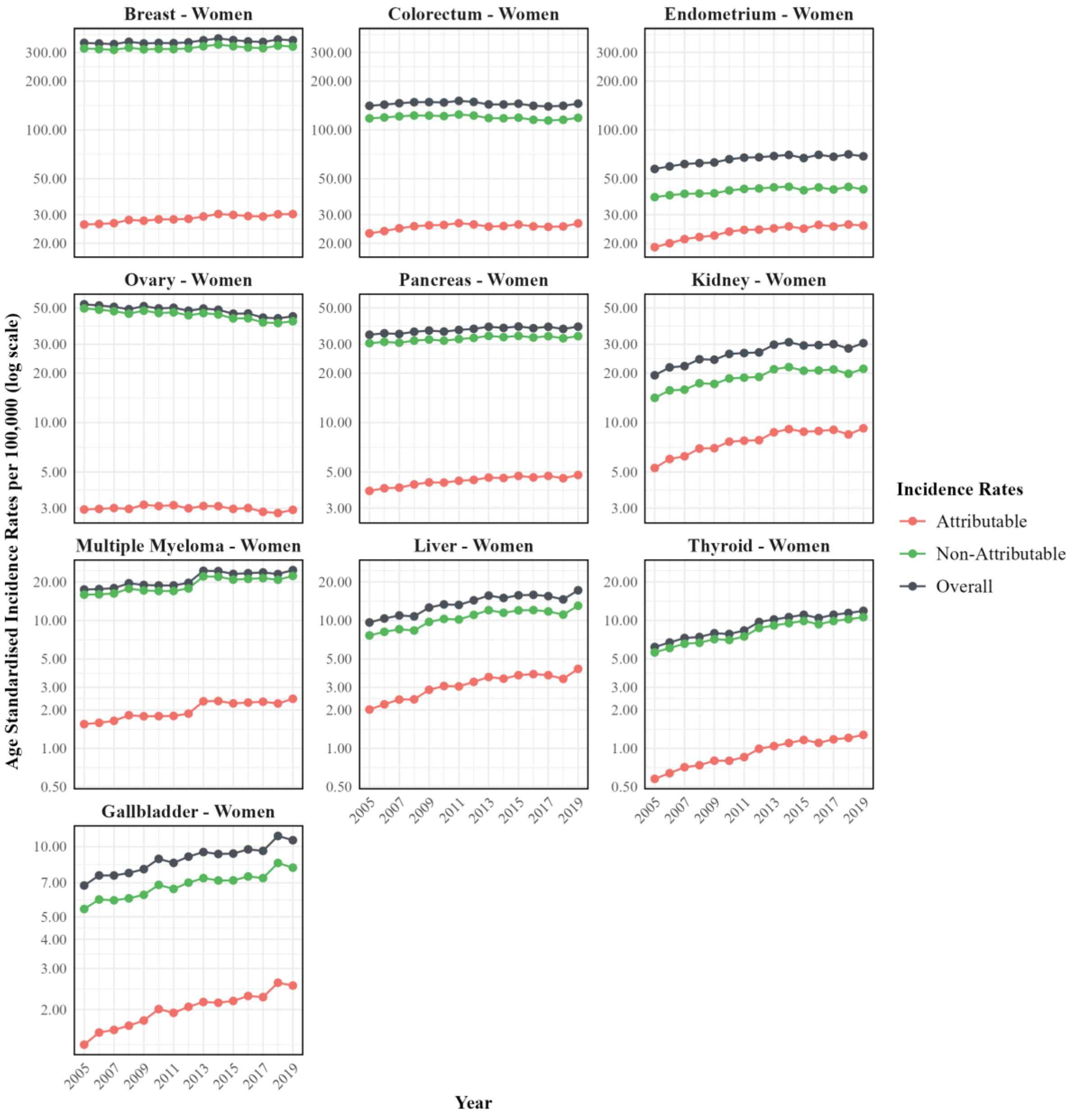

**Panel B**

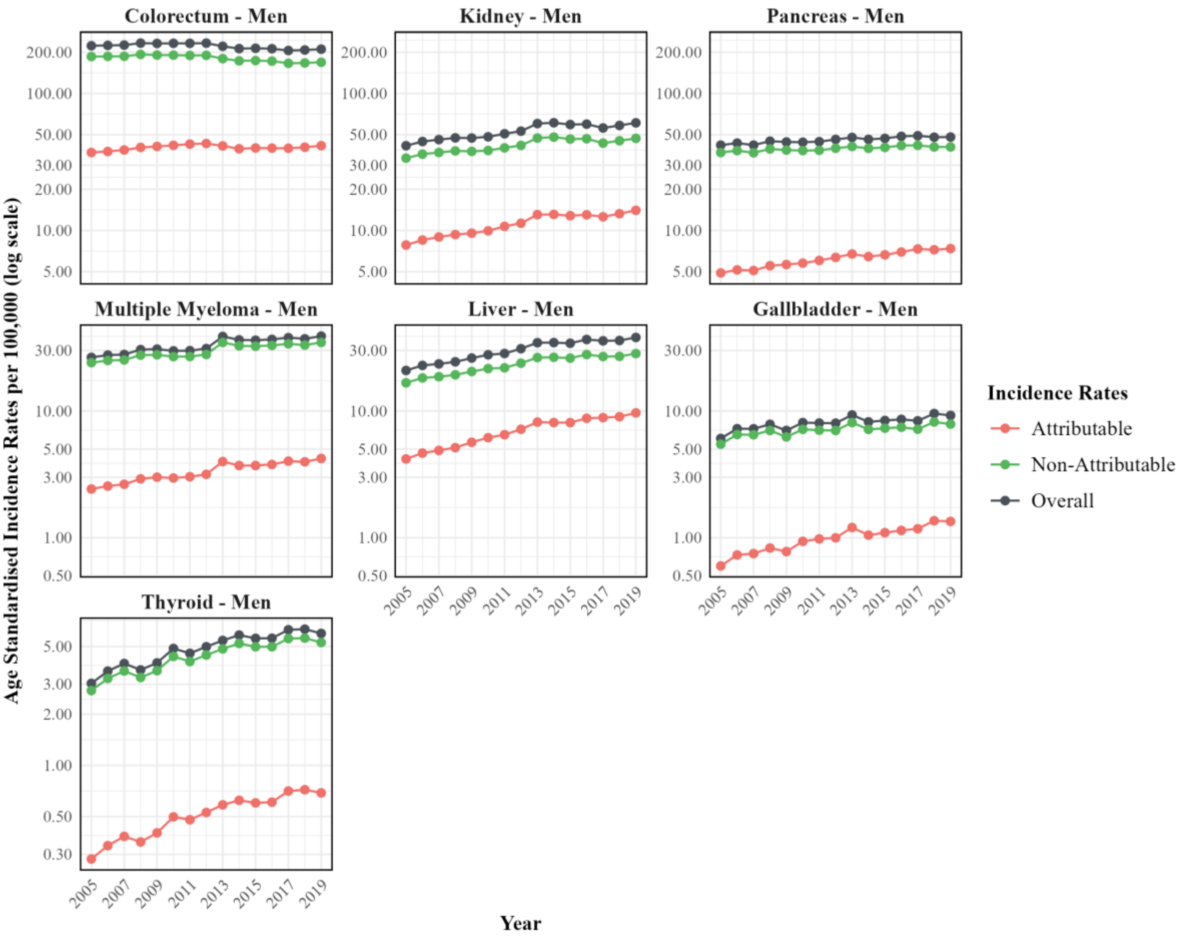

Colorectum - Men
Kidney - Men
Pancreas - Men
Multiple Myeloma - Men
Liver - Men
Gallbladder - Men
Thyroid - Men
Age Standardised Incidence Rates per 100,000 (log scale)
Year
Incidence Rates
Attributable
Non-Attributable
Overall

**Supplementary Figure 6**: Influence of risk factor prevalence and relative risk on the population attributable fraction (PAF)

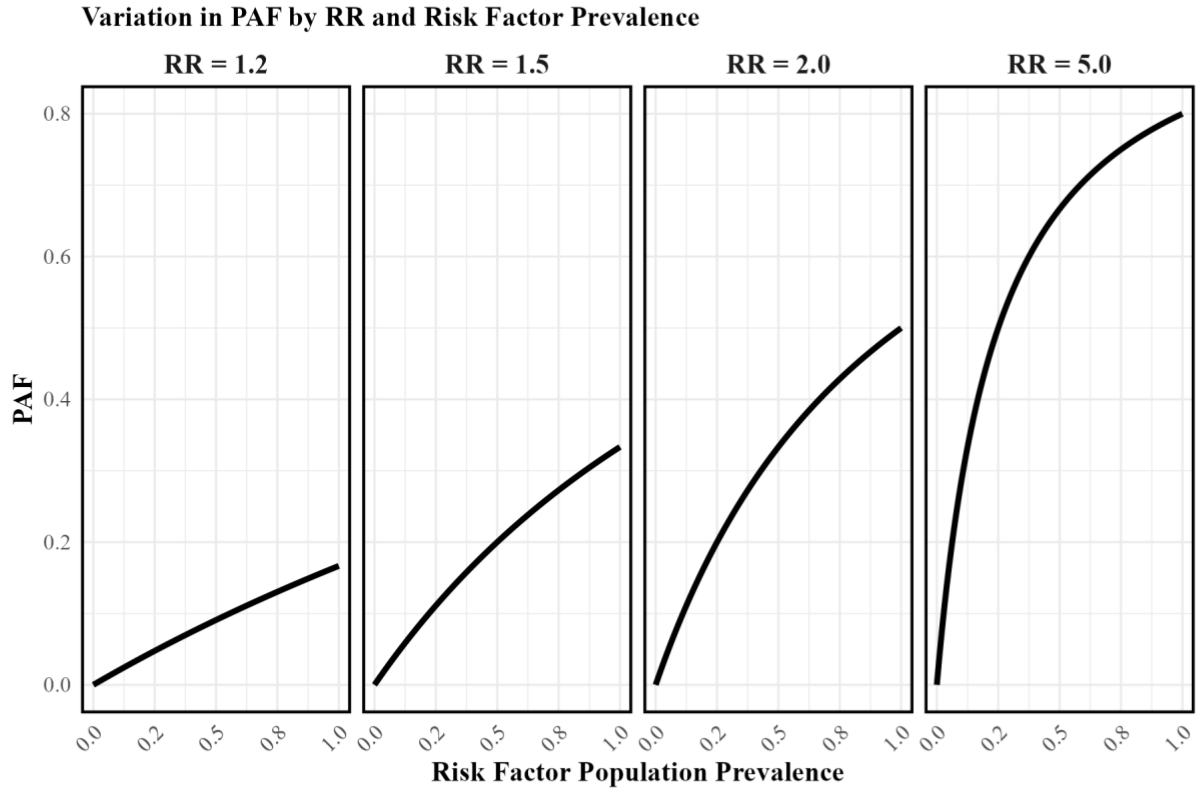

*Risk factors with weak associations (e.g., RR = 1.2) are unlikely to account for a substantial proportion of cancers, even when highly prevalent (e.g., PAF ≈ 20% for 80% prevalence). If such weak risk factors increase over time, their contribution remains minimal, even with large prevalence changes. Risk factors with moderate associations (e.g., RR = 2.0) can explain a meaningful fraction of cases if they are common (e.g., PAF = 20% for 50% prevalence).*

**Supplementary Figure 7**: Proportion of total age-standardised cancer incidence rate by age group for eleven selected cancer sites for women and men in England, 2019.

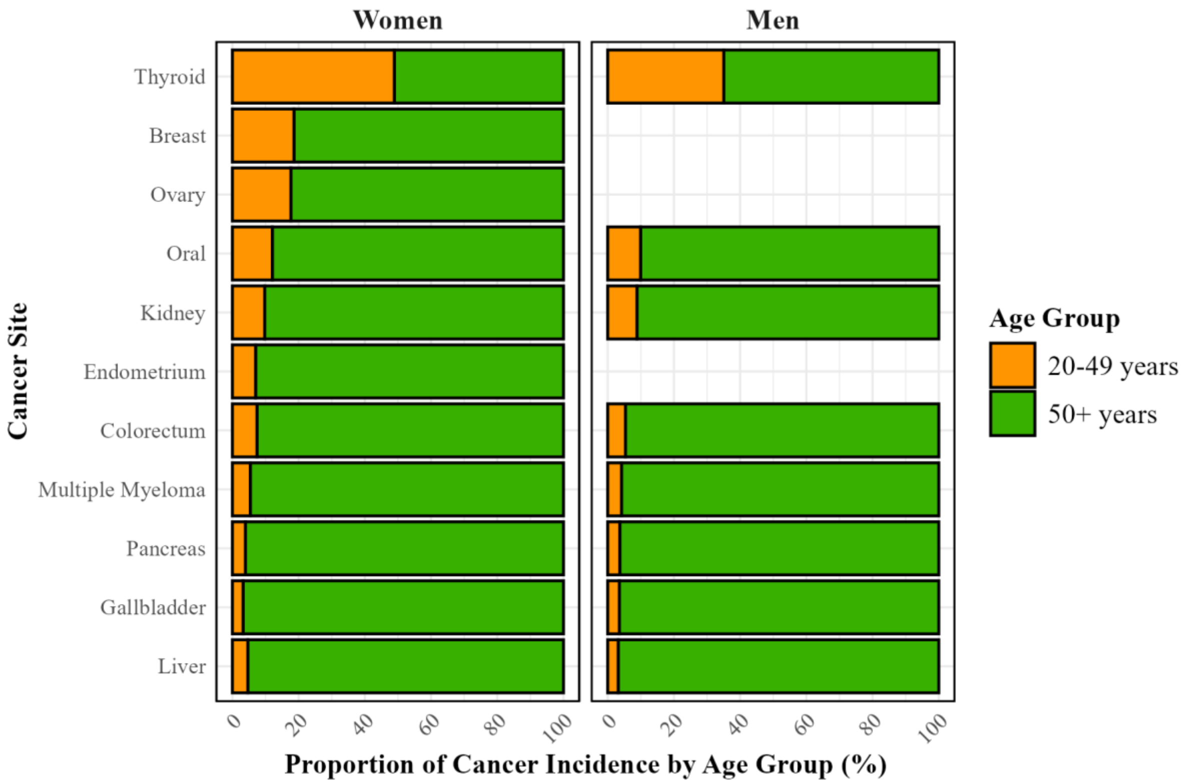

**Supplementary Figure 8**: Trends in early-onset colorectal cancer incidence rates and risk factors in younger adults and children (obesity/overweight only) in England. Cancer incidence and risk factor trends are shown with a 10 year shift to reflect a 10 year lag time between exposure to cancer incidence. Separate panels are shown for women (**Panel A**) and men (**Panel B**)

**Panel A**

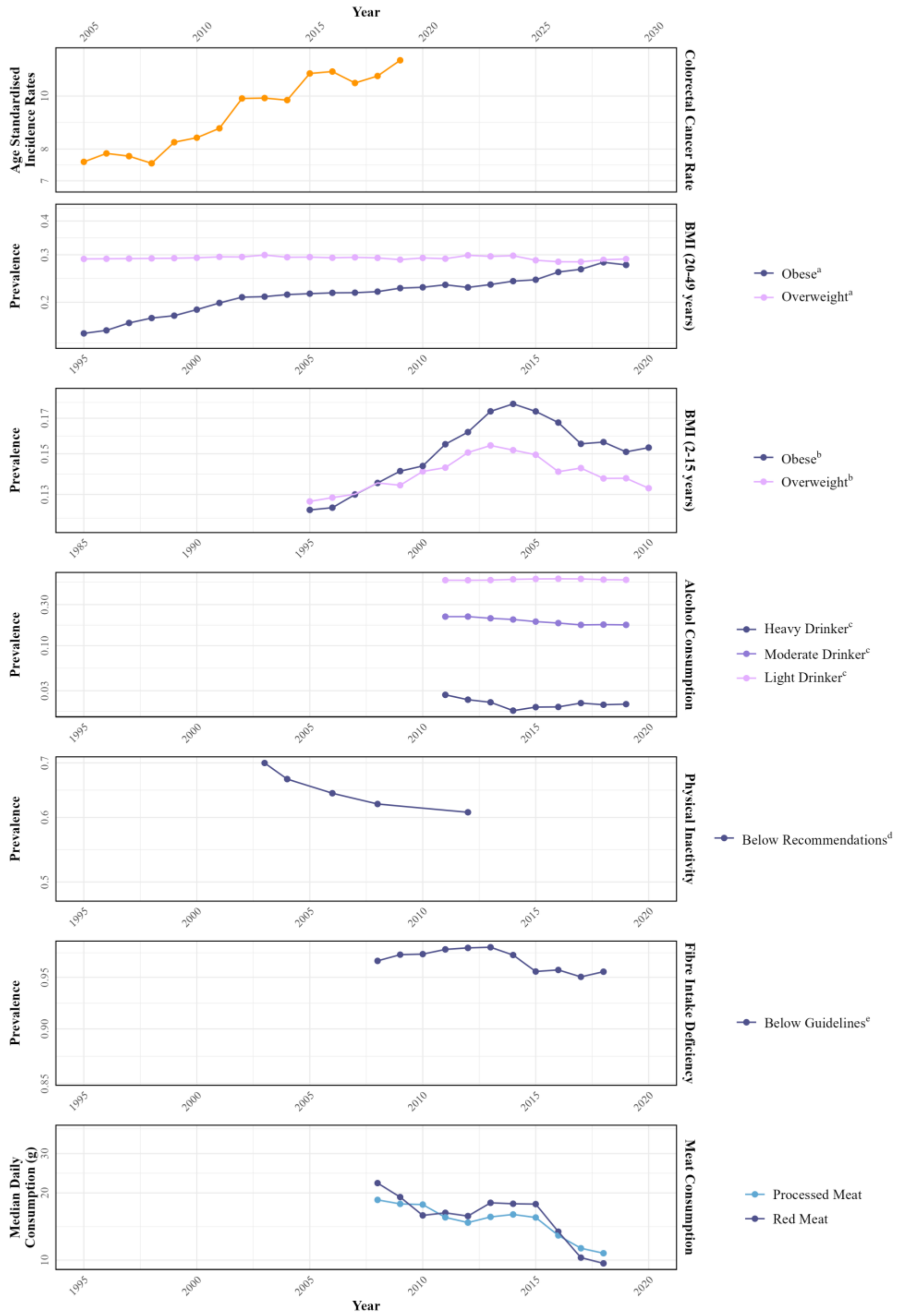

[a] Overweight (>30 $kg/m^2$), obese (25-30 $kg/m^2$)
[b] Overweight , obese
[c] Heavy (>50g/day), moderate (12-50 g/day), light (<12 g/day)
[d] Recommendations are 150 min/week of moderate or 75 min/week or vigorous activity
[e] Guidelines are 30g of fibre per day

### Panel B

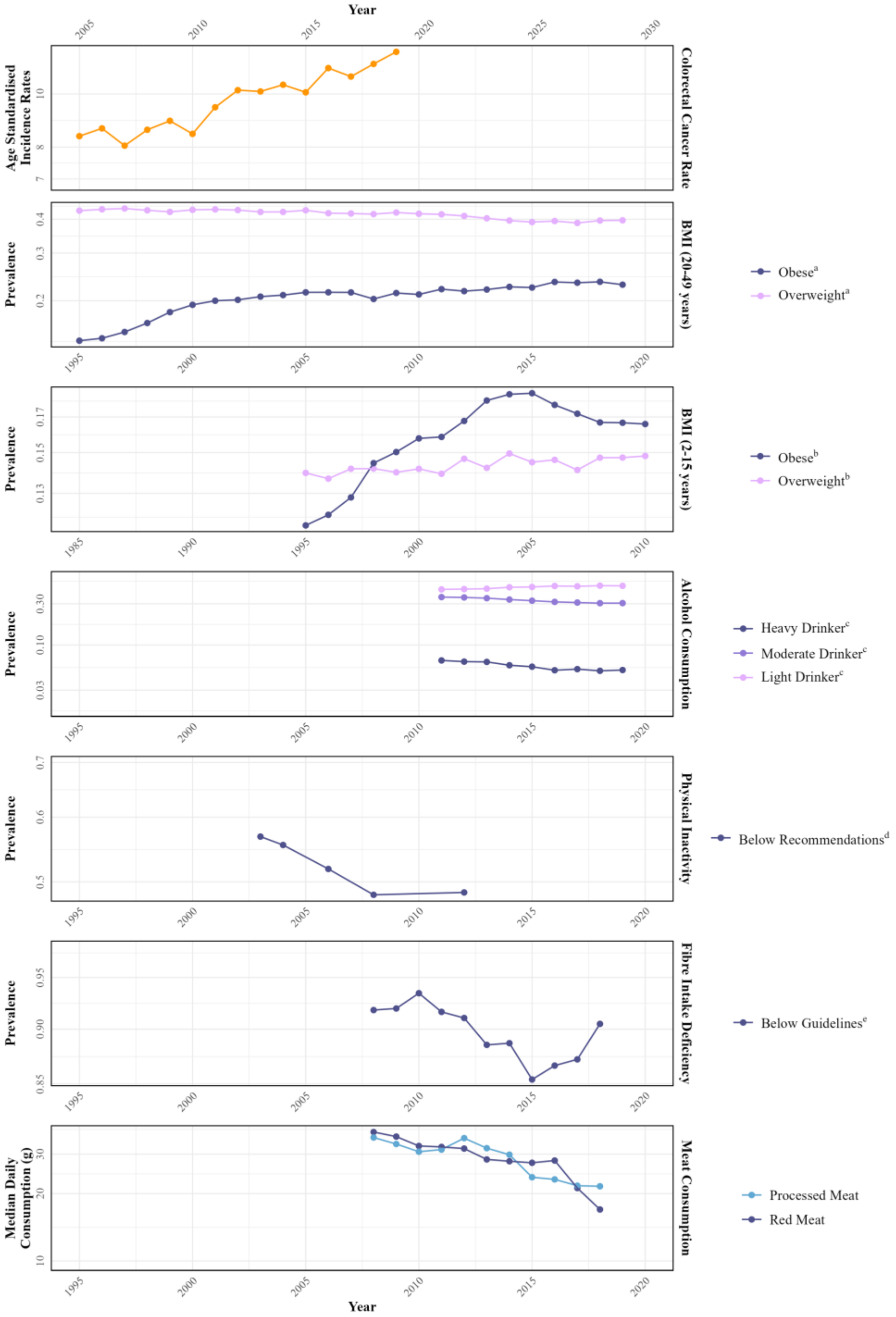

[a] Overweight ($>30$ kg/m$^2$), obese (25-30 kg/m$^2$)
[b] Overweight , obese
[c] Heavy ($>50$g/day), moderate (12-50 g/day), light ($<12$ g/day)
[d] Recommendations are 150 min/week of moderate or 75 min/week or vigorous activity
[e] Guidelines are 30g of fibre per day

**References**

1 Eurostat's task force. Eurostat Methodologies and Working papers. Revision of the European Standard Population. Report of Eurostat's task force. 2013; : 128.

2 Cancer Over Time. International Agency for Research on Cancer. https://gco.iarc.fr/overtime/en/database#cancer-dictionary (accessed Feb 19, 2025).

3 Kim H-J, Fay MP, Feuer EJ, Midthune DN. Permutation Tests for Joinpoint Regression with Applications to cancer rates. *Stat Med* 2000; **19**: 351.

4 Index of Multiple Deprivation (IMD). Consumer Data Research Centre. 2025. https://data.cdrc.ac.uk/dataset/index-multiple-deprivation-imd (accessed Feb 19, 2025).

5 Joinpoint Regression Program. National Cancer Institute. https://surveillance.cancer.gov/joinpoint/ (accessed Feb 19, 2025).

6 Brown KF, Rumgay H, Dunlop C, *et al.* The fraction of cancer attributable to modifiable risk factors in England, Wales, Scotland, Northern Ireland, and the United Kingdom in 2015. *Br J Cancer* 2018; **118**: 1130.

7 Islami F, Marlow EC, Thomson B, *et al.* Proportion and number of cancer cases and deaths attributable to potentially modifiable risk factors in the United States, 2019. *CA Cancer J Clin* 2024; published online July 11. DOI:10.3322/CAAC.21858.

8 Poirier AE, Ruan Y, Volesky KD, *et al.* The current and future burden of cancer attributable to modifiable risk factors in Canada: Summary of results. *Prev Med (Baltim)* 2019; **122**: 140–7.

9 List of classifications by cancer sites with sufficient or limited evidence in humans, IARC Monographs Volumes 1-137. *International Agency for Research on Cancer* 2024; published online Sept 29.

10 Alcoholic drinks and the risk of cancer. *World Cancer Research Fund/American Institude of Cancer Research*.

11 Lauby-Secretan B. Body Fatness and Cancer — Viewpoint of the IARC Working Group. *N Engl J Med* 2016; **375**. https://www.nejm.org/doi/full/10.1056/NEJMsr1606602 (accessed Feb 21, 2025).

12 Exercise and cancer. World Cancer Research Fund. https://www.wcrf.org/preventing-cancer/topics/exercise-and-cancer/ (accessed Feb 21, 2025).

13 Meat and cancer. World Cancer Research Fund. 2018. https://www.wcrf.org/preventing-cancer/topics/meat-and-cancer/ (accessed Feb 21, 2025).

14 Wholegrains, vegetables and fruit and the risk of cancer. *World Cancer Research Fund/American Institude of Cancer Research* https://www.wcrf.org/wp-content/uploads/2024/10/Wholegrains-veg-and-fruit.pdf (accessed Feb 20, 2025).

15 Gandini S, Botteri E, Iodice S, *et al.* Tobacco smoking and cancer: A meta-analysis. *Int J Cancer* 2008; **122**: 155–64.

16 Lee YCA, Cohet C, Yang YC, Stayner L, Hashibe M, Straif K. Meta-analysis of epidemiologic studies on cigarette smoking and liver cancer. *Int J Epidemiol* 2009; **38**: 1497–511.

17 Botteri E, Borroni E, Sloan EK, *et al.* Smoking and Colorectal Cancer Risk, Overall and by Molecular Subtypes: A Meta-Analysis. *Am J Gastroenterol* 2020; **115**: 1940–9.
18 Liu X, Peveri G, Bosetti C, *et al.* Dose-response relationships between cigarette smoking and kidney cancer: A systematic review and meta-analysis. *Crit Rev Oncol Hematol* 2019; **142**: 86–93.
19 Bosetti C, Lucenteforte E, Silverman DT, *et al.* Cigarette smoking and pancreatic cancer: an analysis from the International Pancreatic Cancer Case-Control Consortium (Panc4). *Annals of Oncology* 2011; **23**: 1880.
20 Calle EE, Gapstur SM, Patel A V., *et al.* Ovarian cancer and smoking: individual participant meta-analysis including 28 114 women with ovarian cancer from 51 epidemiological studies. *Lancet Oncol* 2012; **13**: 936.
21 Bagnardi V, Rota M, Botteri E, *et al.* Alcohol consumption and site-specific cancer risk: a comprehensive dose–response meta-analysis. *Br J Cancer* 2014; **112**: 580.
22 Jenabi E, Poorolajal J. The effect of body mass index on endometrial cancer: a meta-analysis. *Public Health* 2015; **129**: 872–80.
23 Xue K, Li FF, Chen YW, Zhou YH, He J. Body mass index and the risk of cancer in women compared with men: a meta-analysis of prospective cohort studies. *Eur J Cancer Prev* 2017; **26**: 94–105.
24 Shi X, Deng G, Wen H, *et al.* Role of body mass index and weight change in the risk of cancer: A systematic review and meta-analysis of 66 cohort studies. *J Glob Health* 2024; **14**: 04067.
25 Chen Y, Wang X, Wang J, Yan Z, Luo J. Excess body weight and the risk of primary liver cancer: An updated meta-analysis of prospective studies. *Eur J Cancer* 2012; **48**: 2137–45.
26 Wang F, Xu Y. Body mass index and risk of renal cell cancer: A dose-response meta-analysis of published cohort studies. *Int J Cancer* 2014; **135**: 1673–86.
27 Wallin A, Larsson SC. Body mass index and risk of multiple myeloma: a meta-analysis of prospective studies. *Eur J Cancer* 2011; **47**: 1606–15.
28 Munsell MF, Sprague BL, Berry DA, Chisholm G, Trentham-Dietz A. Body Mass Index and Breast Cancer Risk According to Postmenopausal Estrogen-Progestin Use and Hormone Receptor Status. *Epidemiol Rev* 2014; **36**: 114.
29 Schmid D, Ricci C, Behrens G, Leitzmann MF. Adiposity and risk of thyroid cancer: a systematic review and meta-analysis. *Obesity Reviews* 2015; **16**: 1042–54.
30 Aune D, Rosenblatt DAN, Chan DSM, *et al.* Anthropometric factors and ovarian cancer risk: A systematic review and nonlinear dose-response meta-analysis of prospective studies. *Int J Cancer* 2015; **136**: 1888–98.
31 Garcia L, Pearce M, Abbas A, *et al.* Non-occupational physical activity and risk of cardiovascular disease, cancer and mortality outcomes: a dose-response meta-analysis of large prospective studies. DOI:10.1136/bjsports-2022-105669.
32 Vieira AR, Abar L, Chan DSM, *et al.* Foods and beverages and colorectal cancer risk: a systematic review and meta-analysis of cohort studies, an update of the evidence of the WCRF-AICR Continuous Update Project. *Annals of Oncology* 2017; **28**: 1788–802.
